# Tumor-infiltrating lymphocytes in breast cancer through artificial intelligence: biomarker analysis from the results of the TIGER challenge

**DOI:** 10.1101/2025.02.28.25323078

**Authors:** Mart van Rijthoven, Witali Aswolinskiy, Leslie Tessier, Maschenka Balkenhol, Joep M. A. Bogaerts, Damien Drubay, Laura Comerma Blesa, Dieter Peeters, Elisabeth Specht Stovgaard, Anne-Vibeke Lænkholm, Harry Haynes, Ligia Craciun, Denis Larsimont, Mohamed T. Amgad, Lee AD Cooper, Cyril de Kock, Valerie Dechering, Johannes Lotz, Nick Weiss, Mieke van Bockstal, Christine Galant, Esther Lips, Hugo M. Horlings, Jelle Wesseling, Lennart Mulder, Sandra van den Belt, Karsten Weber, Paul Jank, Carsten Denkert, Enrico Munari, Giuseppe Bogina, Chris Russ, Alex Lemm, Sherene Loi, Julia Dixon Douglas, Stephan Michiels, Heikki Joensuu, Ming Fan, Daehong Lee, Jaehyung Ye, Kangwon Byun, Jeongyeol Kim, Shuoyu Xu, Zheng Ji, Feng Xie, Jinbo Kuang, Xulin Chen, Liliang Chen, Anna Maria Tsakiroglou, Richard Byers, Martin Fergie, Vishwesh Ramanathan, Anne L. Martel, Adam Shephard, Shan E Ahmed Raza, Mostafa Jahanifar, Nasir M Rajpoot, Sungduk Cho, Dong-Hee Kim, Hyungjoon Jang, Chanmin Park, Kyungdoc Kim, Rogier Donders, Scott Maurits, Miriam Groeneveld, Anne Mickan, James Meakin, Bram van Ginneken, Roberto Salgado, Jeroen van der Laak, Francesco Ciompi

**Affiliations:** Radboud University Medical Center, Nijmegen, The Netherlands; Division of Cancer Research, Peter MacCallum Cancer Centre, Melbourne, Australia; Department of Pathology, ZAS Hospitals, Antwerp, Belgium; Center for Medical Image Science and Visualization, Linköping University, Linköping, Sweden; Canisius Wilhelmina Ziekenhuis, Nijmegen, The Netherlands; Gustave Roussy, Université Paris-Saclay, Villejuif, France; Hospital del Mar, Barcelona, Spain; Universitair Ziekenhuis Antwerpen, Antwerpen, Belgium; Department of Pathology, Herlev and Gentofte Hospital, Herlev, Denmark; Zealand University Hospital, Køge, Denmark; Great Western Hospitals NHS Foundation Trust, Swindon, UK; Pathology, Institut Jules Bordet, Université Libre de Bruxelles (ULB), Brussels, Belgium; Department of Pathology, Northwestern University Feinberg School of Medicine, Chicago, IL, USA; Fraunhofer Institute for Digital Medicine MEVIS, Lübeck, Germany; Department of Pathology, Cliniques Universitaires Saint-Luc, Brussels, Belgium; The Netherlands Cancer Institute (NKI), Amsterdam, The Netherlands; GBG German Breast Group, Neu-Isenburg, Germany; Institute of Pathology, Philipps University Marburg and Marburg University Hospital (UKGM), UCT Frankfurt-Marburg, Marburg, Germany; Department of Pathology and Diagnostics, University and Hospital Trust of Verona, Verona, Italy; IRCCS Sacro Cuore Don Calabria Hospital, Negrar di Valpolicella, Verona, Italy; Amazon Web Services EMEA SARL (AWS Europe), Luxembourg; The Sir Peter MacCallum Department of Medical Oncology, The University of Melbourne, Parkville, Australia; Department of Oncology, Helsinki University Hospital and University of Helsinki, Helsinki, Finland; Aivis Inc, Seoul, Republic of Korea; Bio-totem Pte Ltd, Suzhou, P.R. China; Cells Vision (Guangzhou) Medical Technology Inc., China; Spotlight Pathology Ltd, Manchester, United Kingdom; Department of Medical Biophysics, University of Toronto, Toronto, ON, CA; Physical Sciences, Sunnybrook Research Institute, Toronto, ON, CA; Tissue Image Analytics Centre, Department of Computer Science, University of Warwick, Coventry, UK; VUNO Inc., Seoul, South Korea; Department of Pathology, Korea University Guro Hospital, Korea University College of Medicine, Seoul, Korea; Pôle MORF, Institut de Recherche Expérimentale et Clinique (IREC), Université catholique de Louvain, Brussels, Belgium; Department of Clinical Medicine, University of Copenhagen, Denmark

## Abstract

The prognostic significance of tumor-infiltrating lymphocytes (TILs) in breast cancer has been recognized for over a decade. Although histology-based scoring recommendations exist to standardize visual TILs assessment, interobserver agreement and reproducibility are hampered by heterogeneous infiltration patterns, highlighting the importance of computational approaches. Despite advances to automate TILs quantification, adoption of computational models has been hindered by lack of consensus on scoring methods and lack of large-scale benchmarks. To address these limitations, we launched the international TIGER challenge, a public competition to build open-source computational TILs (cTILs) models in digital pathology. Here, we present the largest comprehensive multi-centric validation of multiple cTILs methods on surgical resections and biopsies using 3,708 Triple Negative Breast Cancer (TNBC) and human epidermal growth factor receptor 2 positive (HER2+) breast cancers from clinical practice and phase 3 clinical trials. We report benchmarks on image analysis performance of each method and show the strong agreement of cTILs with panels of pathologists. We show the positive association of cTILS with response after neoadjuvant therapy in HER2-positive, superior to visually scored TILs. We also show that cTILs add independent information to clinical variables in surgically resected TNBC but not in HER2-positive disease and breast biopsies.

## Introduction

Since the beginning of the 20th century, clinicians observed that breast cancer (BC) patients with higher levels of lymphocytic infiltration in tumors, now defined as tumor-infiltrating lymphocytes (TILs), often had a better prognosis compared to patients with lower levels^1^. Over the years, high TILs density has also been shown to be associated with better responses to immunotherapy and chemotherapy. However, the lack of a formal and universally shared method to quantify the TILs, hampered for many years the translation of those clinical observations into a biomarker, until recently.

In 2015, the International Immuno-oncology Biomarker Working Group (also known as the TILs WG) proposed recommendations^2^ on evaluating and measuring stromal TILs (sTILs) in hematoxylin and eosin (H&E) stained histopathology slides of BC, *de facto* reviving interest in TILs and their role as a biomarker in immuno-oncology^3^. Using these recommendations, research initially focused on measuring inter-observer variability in visual TILs quantification. A study conducted by the TILs WG^4^ found very good concordance rates across several clinically relevant cut-points, but also reported inter-observer disagreement, for example in the presence of heterogeneous infiltration of immune cells^5^. A more recent study reported substantial interobserver disagreement when TILs are scored on core-needle biopsies^6^. A large study on a pooled individual patient analysis from phase 3 clinical trials^7^ showed the prognostic value of sTILs as a biomarker for cancer recurrence and patient survival in early-stage triple-negative breast cancer (TNBC). Studies^8,9^ have also shown the correlation of sTILs with pathological complete response (pCR) following neoadjuvant chemotherapy (NACT) in early-stage TNBC and human epidermal growth factor receptor 2 positive (HER2+) breast cancers, as well as correlation with cancer recurrence and survival, regardless of the hormone receptor (HR) status. Recent initiatives are investigating the impact of TILs as a biomarker to forgo chemotherapy in TNBC cases^10^.

Aiming at reducing inter-observer variability and complementing pathologists where TIL-evaluation can be difficult, researchers have focused on artificial intelligence (AI) models^11^ to automate computational TILs (cTILs) quantification in digital pathology whole-slide images (WSI). Most presented models rely on deep learning to classify small image regions (i.e., patches)^12–14^, analyze tissue morphology (i.e., detect cells and segment tissue)^8,15–18^, also using open-source tools^19,20^, or end-to-end learning from raw data^21^. The bulk of work on cTILs has focused on breast cancer^5,9,16,18,19,21–25^, but similar solutions have also been proposed for other cancer types such as melanoma^20^, gastric^12^, head and neck^17^, lung^26,27^, oropharyngeal^15^ and testicular cancer^28^. In colorectal cancer, the Immunoscore^29^ has been proposed to quantify immune response based on the immunohistochemical (IHC) analysis of CD8- and CD3-positive T lymphocytes at the central part and invasive front of the tumor. Partly inspired by the Immunoscore approach, AI-based quantification of peritumoral lymphocytes in H&E-stained slides has been found to hold prognostic value in TNBC^30^. However, so far no computational method has been considered to have sufficient level of clinical validity to be usable in daily practice settings. This is mostly due to a) the lack of consensus or recommendations on how to design computational scoring methods, and b) the lack of large-scale studies to comprehensively benchmark cTILs across multiple clinical end-points, including correlation with visual assessment in both biopsies and surgical resections, and clinical validation on large scale clinical trials.

In response to these needs, we created a public competition on Tumor InfiltratinG lymphocytes in breast cancER (TIGER) in the form of a two-phase Grand Challenge (https://tiger.grand-challenge.org/) (Supplementary section *TIGER challenge design*), focusing on the most aggressive BC, namely TNBC and HER2+ breast cancer. With TIGER, we involved the scientific community in a quest to develop AI models to analyze the morphology of WSIs and to use the output of this analysis to engineer an automatic cTILs quantification algorithm in whole-slide images. We publicly released a multi-centric dataset of images and manual annotations to allow participants to train AI models (Figure 1a), and a public evaluation procedure on the Grand Challenge^31^ (GC) platform. Via GC, we benchmarked AI models both from a computer vision perspective (i.e., the automated identification of tumor and stroma tissue regions in images) and from a clinical perspective (i.e., the analysis of the prognostic value of cTILs). We fostered a privacy-preserving benchmarking procedure by implementing, for the first time on the Grand Challenge platform, validation on sequestered (i.e., not directly accessible by participants) data using entire whole-slide images. We also promoted an open and reproducible science approach by releasing training data, the code of baseline AI model and evaluation metrics, and required all participants to open source their final cTILs solution.

**Figure 1.**
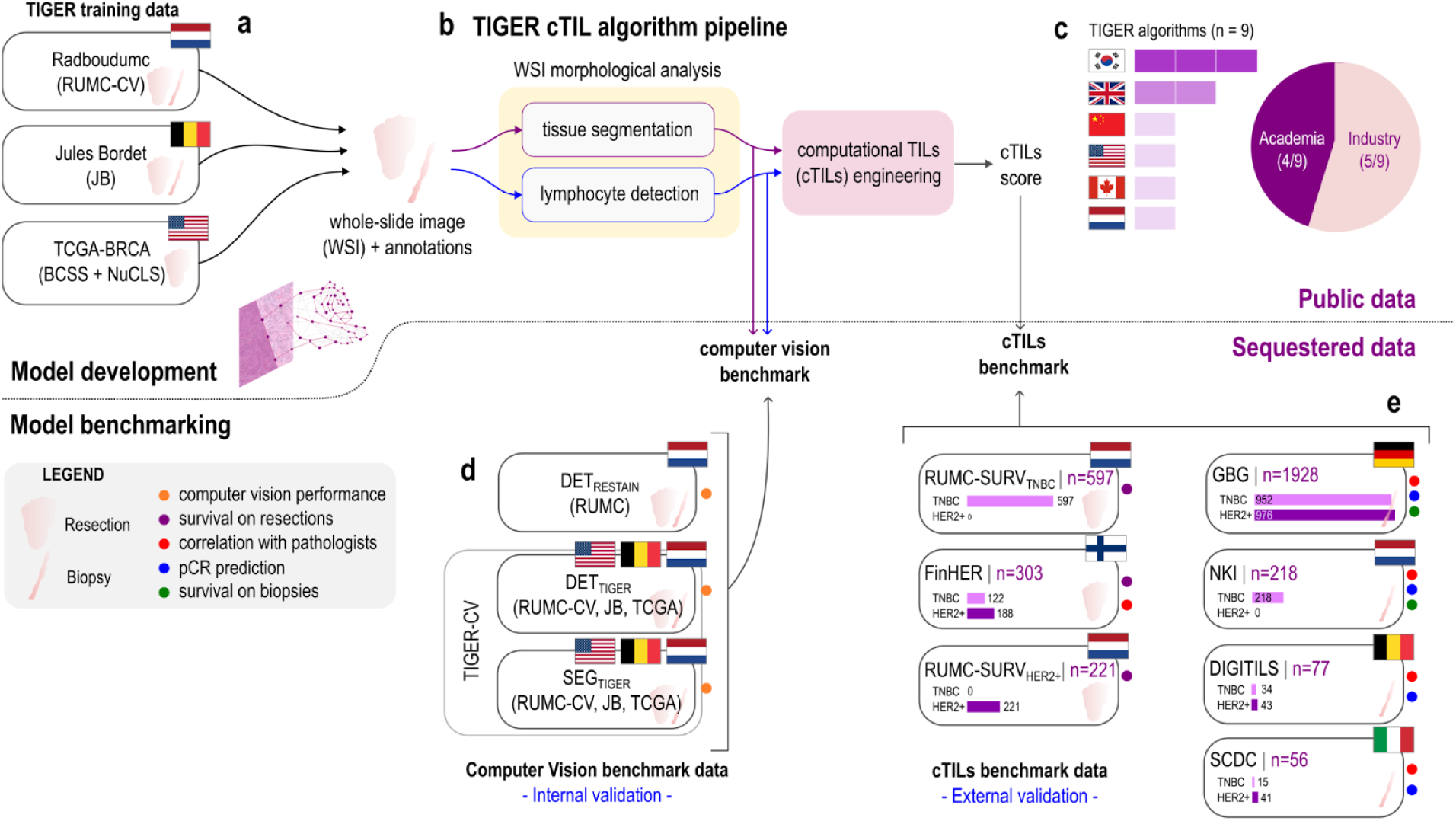
Overview of cTILs algorithm development and benchmarking. **a,** Publicly available multi-centric training data, containing WSIs with corresponding manual annotations released as part of the TIGER challenge to train the morphological analysis component of cTILS models. **b**, Schematic overview of the components of cTILS models developed in TIGER. Manually annotated training WSIs were used to build computer models for tissue segmentation and lymphocyte detection. The output of these models was used to engineer a cTILs score. Both models for morphological analysis and the cTILs score were benchmarked in this study. **c**, From the final phase of TIGER, we obtained a total of nine AI models from six different countries, both from academia (n=4) and industry (n=5). **d**, Sequestered data used to benchmark the computer vision performance (i.e., lymphocyte detection and tissue segmentation) of the morphological analysis module. We used the multi-centric TIGER-CV dataset (the test set of the final phase in the computer vision task of TIGER, consisting of DETTIGER for cell detection and SEGTIGER for tissue segmentation) and DETRESTAIN for cell detection, with an immunohistochemistry-based reference standard. Given the common source with TIGER training data, this is considered internal validation data. **e,** Sequestered data used to benchmark several aspects of cTIL): i) prognostic value on surgical resections, ii) prognostic value on biopsies, iii) correlation with pathologists’ visual sTILs, iv) prediction of pCR in neoadjuvant chemotherapy treatment. We included both HER2+ and TNBC in all sets. Since there is no overlap with TIGER training data, we consider this as external validation data. See Table *2* for an overview on data acquisition processes. Abbreviations: cTILs: computational tumor-infiltrating lymphocytes; HER2+: human epidermal growth factor receptor 2 positive; TNBC: triple-negative breast cancer; TCGA: The Cancer Genome Atlas; DET: detection; SEG: segmentation; WSI: whole-slide image; pCR: pathological complete response. Legend: orange is computer vision performance, purple is survival on resections, red is correlation with pathologist, blue is pCR prediction, green is survival on biopsies.

The output of the TIGER challenge is the foundation of the study presented in this paper. While retracing the main phases of the challenge, we present here a comprehensive post-challenge analysis to benchmark various aspects of cTILS models obtained from the final phase of TIGER. To the best of our knowledge, we established the largest multicenter international dataset for cTILs validation, consisting of n=3,708 early-stage breast cancers (n=1,938 TNBC, n=1,770 HER2+, see Methods section *Materials*) from clinical practice and phase 3 clinical trials, including both surgical resections and preoperative (core-needle) biopsies. For each cTILs algorithm, we first report results of the *computer vision benchmark* (Figure 1d) that measures tissue segmentation and lymphocyte detection performance. For this, we used an internal validation dataset from the TIGER challenge and a post-TIGER challenge dataset for which the reference standard was based on restaining cases with lymphocyte-sensitive immunohistochemistry stains (Supplementary section *Re-stained data*). Second, we report results of the *cTILs benchmark* using a large external validation set (Figure 1e, Table 2) that is completely independent from the TIGER training data. The cTILs benchmark measures a) the correlation of cTILs with visual scoring of pathologists in both surgical resections and biopsies; b) the association between cTILs and pCR in a NACT setting, using core-needle biopsies as input; and c) the prognostic value of cTILs (assessed on both biopsies and surgical resections) in terms of disease-free interval (DFI) and overall survival (OS).

## Results

We validated all cTILS models that made valid submissions to the final phase for survival evaluation of the TIGER challenge, as well as the top-3 AI models for the final phase for the computer vision evaluation, resulting in nine open-source AI models (Figure 1c): `aivis`^1^, `biototem`, `cellsvision`, `didsr`, `radboud`^2^, `spotlight`, `sri`^3^, `tiager`, `vuno` (see Supplementary sections *TIGER challenge evaluation* and *Survival track* for more information about the TIGER challenge setup and the survival phase. Supplementary section *TIGER Models*, includes the descriptions of the AI models).

### Computer vision benchmarks

We evaluated the performance of cTILs models to analyze the morphology of whole-slide images, consisting in a) the segmentation (i.e., pixel classification) of tumor and stroma regions and b) detection (i.e., prediction of location) of lymphocytes.

For *segmentation*, we used the SEG_TIGER_ subset of TIGER-CV from the final phase of the computer vision track of TIGER, consisting of 286 regions of interest (ROIs), sampled from 195 H&E WSIs, with exhaustive manual annotations (i.e., all pixels annotated) of three tissue classes (tumor, stroma and other tissue). For each algorithm, we computed the Dice score, which measures the overlap between each segmented region and the corresponding manual annotations, and considered it for the tumor and the stroma classes over the whole set of ROIs. We found that all AI models segmented tumor equally well as stroma (Figure 2a), with a slightly better performance on stroma segmentation (average Dice=0.75) vs tumor segmentation (average Dice=0.71).

**Figure 2.**
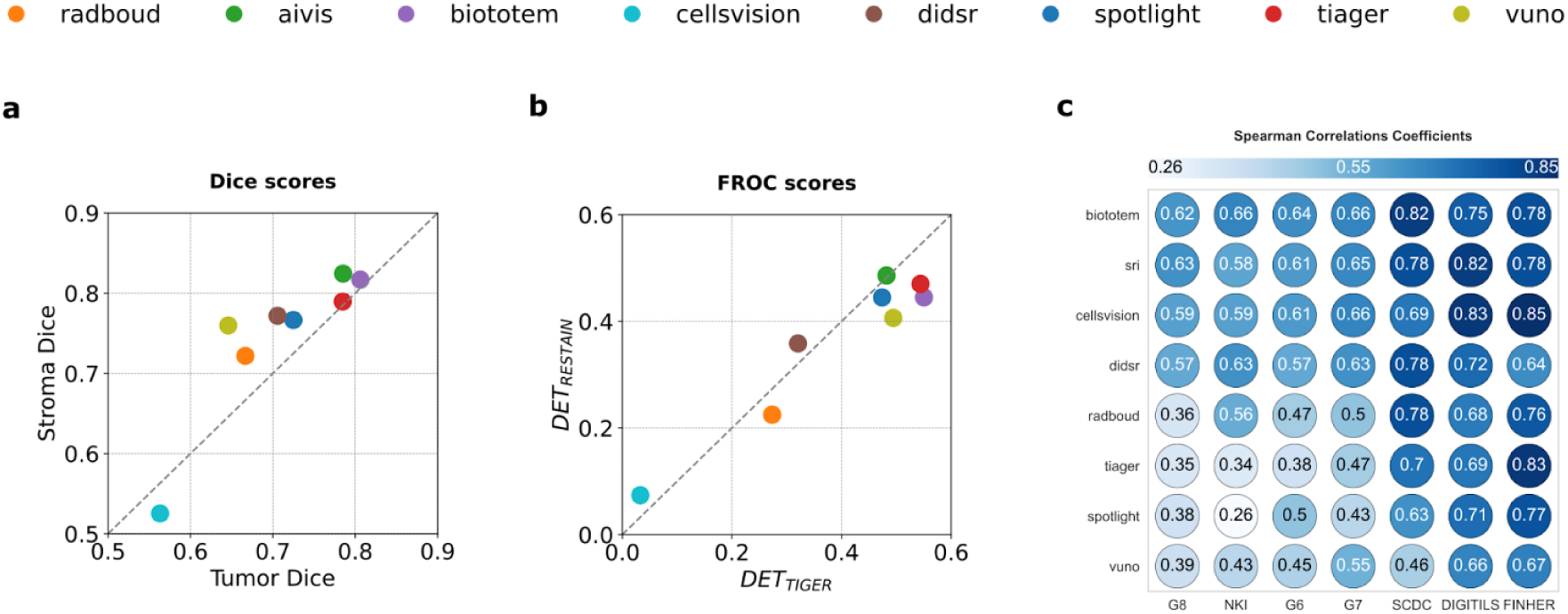
Results of TIGER models for computer vision performance and correlation with pathologists. **a**, Scatter plot of Dice score for tumor versus stroma for each algorithm. **b**, Scatter plot of FROC scores for each algorithm when tested on DETTIGER versus DETRESTAIN. **c**, Spearman rank correlation coefficients for cTILs from each final TIGER algorithm with vTILs assessed by pathologists for each cohort; note that the `aivis` algorithm did not produce a cTILs score and is excluded from this analysis. Each circle’s color and numerical value indicate the correlation coefficient. Rows and columns are sorted based on the average correlation per algorithm (rows) and per dataset (columns). Abbreviations. FROC: Free-Response Receiver Operating Characteristic; DET: detection; cTILs: computational TILs; vTILs: visual TILs, computed as a combination of visual TILs assessments by a group of pathologists.

For *detection*, we used the DET_TIGER_ and DET_RESTAIN_ datasets. DET_TIGER_ was the set of manual lymphocyte annotations in TIGER-CV (Figure 1d), consisting of 1,879 ROIs taken from 168 H&E WSIs, with lymphocyte locations annotated on the slide by five pathologists, one pathologist per slide. DET_RESTAIN_ was built post-challenge using paired images of the same slides stained with H&E first and then re-stained with IHC (CD3 for T lymphocytes and CD79a for B lymphocytes). Manual annotations of lymphocyte locations were made on the IHC slide and transferred to the H&E image using an algorithm of image registration^32^ (i.e., image alignment). For each algorithm, we computed Free-Response Receiver Characteristics (FROC)^33^ curves and derived a FROC score by averaging sensitivities computed at 10, 20, 50, 100, 200 and 300 false positives per *mm*^2^. Results are depicted in Figure 2b. FROC scores show that the AI models detect lymphocytes equally well on DET_TIGER_ and DET_RESTAIN_, although with a general trend to lower scores in DET_RESTAIN_ (range [0.07-0.49] in DET_RESTAIN_ vs. [0.03-0.55] in DET_TIGER_). Using DET_RESTAIN_ served both as a way to extend the benchmark including an objective reference standard, and as an indirect way to compare, although qualitatively, manual lymphocyte annotations used in TIGER with IHC-based annotations. Overall, we found that performance rank of AI models is similar in segmentation and detection tasks resulting in `biototem`, `tiager` and `aivis` as the three top-performing AI models. For detailed descriptions of the datasets, annotations, evaluation metrics, and supplementary analyses, refer to Supplementary sections *Building Annotated Data for TIGER*, *Re-stained Data*, *TIGER Challenge Evaluation*, and *Online Methods: Computer Vision Performance*, as well as Supplementary Table 4 for the final computer vision performance.

### Computational TILs benchmarks

We analyzed cTILs scores using several benchmarks, namely a) the correlation of cTILs with visual TIL scores from pathologists, b) the association of cTILs with response to neoadjuvant chemotherapy, c) the prognostic value of cTILs considering disease-free interval (DFI) and overall survival (OS) as end points.

#### cTILs show on average strong correlation with visual TILs

We analyzed the correlation between cTILs and visual scoring of stromal TILs (sTILs) assessed by pathologists following the TIL WG recommendations^2^. We used data from 2,582 BC cases, including both biopsies and resections for which visual TIL scores were available. For biopsies, we used 2,279 WSIs from four cohorts (Figure 1e): DIGITILS (n=77), three cohorts from the GBG (n=1,928 in total) consisting of GeparSixto (G6, n=553), GeparSepto (G7, n=617), GeparOcto (G8, n=758), NKI (n=218) and SCDC (n=56). For resections, we used a subset of 303 cases from FinHER (Method section *Data*). In all cohorts, visual TILs were scored by one or more pathologists (range [1-40]). Following Van Bokstal et al.^6^, we computed a “median” pathologist score (vTILs) as the slide-level median sTIL (Method section *Combination of sTILs*) and used it as a reference to compute the Spearman rank correlation coefficient (ρ) with cTILs. Figure 2c depicts an overview of the correlation values ranked per algorithm and per dataset (Supplementary Section: *Scatter Correlations* shows the trends of each pair of vTILs and cTILs). We found that all cTILs showed on average strong correlation with vTILs (mean ρ value = 0.61, range [0.26-0.85]), with top-3 highest correlations for `biototem`, `sri` and `cellsvision`, and on average highest correlation on the FinHER dataset, which contains surgical resections. In some cases, weak correlation was found in G8 and NKI for `radboud`, `tiger`, `spotlight` and `vuno`. Interestingly, these methods mostly used a concave hull-based approach to define the region of the tumor bulk in the slide (Supplementary section *TIGER AI Models*). In contrast, `sri` and `cellsvision` adopted alternative solutions to use tumor segmentation to define the region to score cTILs. This suggests the importance of careful design choices on the definition of the tumor bulk region, where the concave hull approach might be too sensitive to noisy tumor segmentation outputs, and might be mitigated by improving tumor segmentation accuracy, as in the case of `biototem`.

### cTILs show association with response to neoadjuvant chemotherapy in HER2+ breast cancer

We benchmarked the association of cTILs with neoadjuvant chemotherapy (success of NACT), formulated as prediction of pCR treated as a binary target, and measured performance via Receiver Operating Characteristic (ROC) analysis and the Area Under the (ROC) Curve. We used cohorts of pre-operative biopsy images of patients treated with NACT from both clinical practice (NKI, DIGITILS, SCDC) and GBG clinical trials G6, G7 and G8 (Online Methods section: *Materials*). In TNBC (Figure 3a), all vTILs showed AUC values > 0.5 (range [0.59-0.81]), whereas some cTILs reported AUC < 0.5 (range [0.47-0.87]). No cTILs achieved statistical significance in G6, and in SCDC neither cTILs nor vTILs showed significant association with NACT. In HER2+ (Figure 3b), cTILs showed AUC values (range [0.52-0.78]) comparable or higher than vTILs (range [0.55-0.65]) in most cohorts. Notably, the `sri` and `didsr` methods always achieved AUC higher than vTILs in HER2+ cases and in two TNBC cohorts.

**Figure 3:**
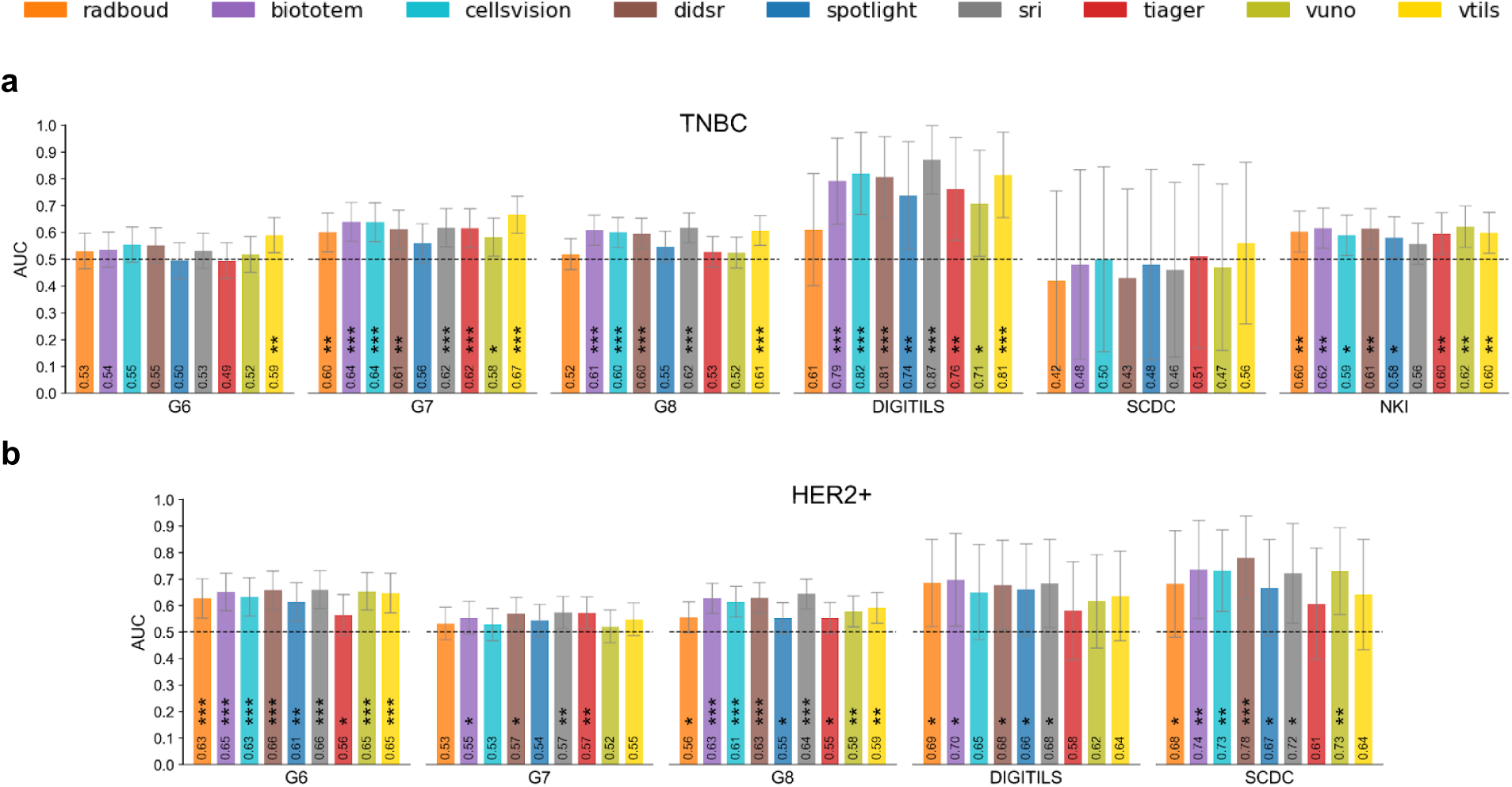
AUC for pCR prediction on biopsies. **a,** Bar plots of AUC values and their confidence interval for predicting pCR after NACT using cTILs and vTILs across different biopsy datasets of TNBC cases. **b**, Bar plots for HER2+ cases. Note that the NKI cohort solely contains TNBC cases. Abbreviations: AUC: Area Under the Curve,NACT: neoadjuvant chemotherapy, pCR: pathological complete response, cTILs: computational TILs, vTILs: visual TILs; TNBC: triple-negative breast cancer, HER2+: human epidermal growth factor receptor 2 positive. * p<0.05, ** p<0.01, *** p<0.001.

To gain additional insights on the overall association with success of NACT for each algorithm, we ranked them based on their AUC value for the TNBC and HER2+ cohorts. Results are depicted in Table 1. We observed that a) the top-ranking AI models are those that achieve the overall higher correlation with pathologists (`biototem`, `sri`, `cellsvision`, `didsr`) (see Figure 2c), b) most of these models are in the top-3 positions in both TNBC and HER2+, c) vTILs are ranked first for TNBC but showed an overall association with NACT success lower or comparable with cTILs due to lower AUC values in HER2+.

**Table 1.**
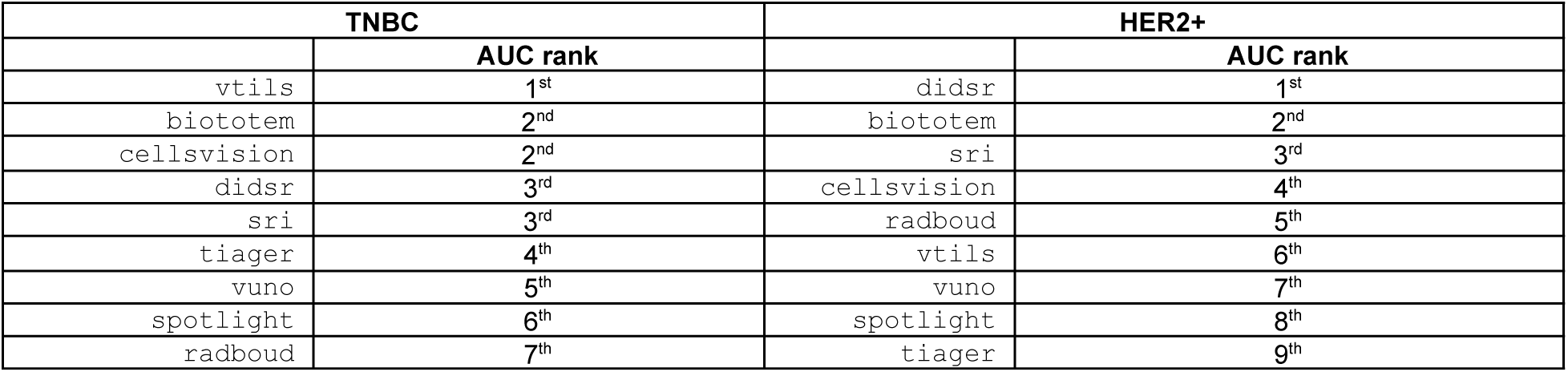
Ranking of cTILs and vTILs based on their AUC values. Each method and visual scores of the “median” pathologist are ranked based on their AUC value for pCR prediction in the TNBC (left) and the HER2+ (right) cohorts.

#### On the prognostic value of cTILs

We analyzed the prognostic value of cTILs on both pretreatment biopsies and surgical resections. From the set of 3,407 cases (see Figure 1e) we used 3,227 cases from 7 cohorts (RUMC-TNBC, RUMC-HER2+, FinHER, NKI, G6, G7, G8), for which survival data was available. Clinical endpoints were Disease-Free Interval (DFI) and Overall Survival (OS).

#### Resections

For surgical resections, we defined two datasets: 1) RES_DFI_, containing n=907 cases from RUMC-SURV_TNBC_ and FinHER, for which DFI information was available; 2) RES_OS_, containing n=1,128 cases from RUMC-SURV_TNBC_, RUMC-SURV_HER2+_ and FinHER, for which OS information was available.

All cTILs identified two subgroups of BC patients with different prognosis via dichotomization based on per-algorithm median cTILs value, with more favorable prognosis correlating with higher cTILs. This is visible in the Kaplan-Meier (KM) curves of cTILs (Figure 4a), only showing the results for cTILs achieving the lowest p-value via log-rank test on RES_DFI_ and RES_OS_, where significant difference was found in TNBC but not in HER2+ subtypes. The KM curves for all models are depicted in Supplementary section *Resections - Kaplan-Meier curves*. In univariable Cox regression analysis for TNBC, all cTILs showed significant correlation (p<0.001) with patient outcome in both RES_DFI_ and RES_OS_. In contrast, for HER2+, none of the models showed significant associations with either DFI or OS (Figure 4b).

**Figure 4:**
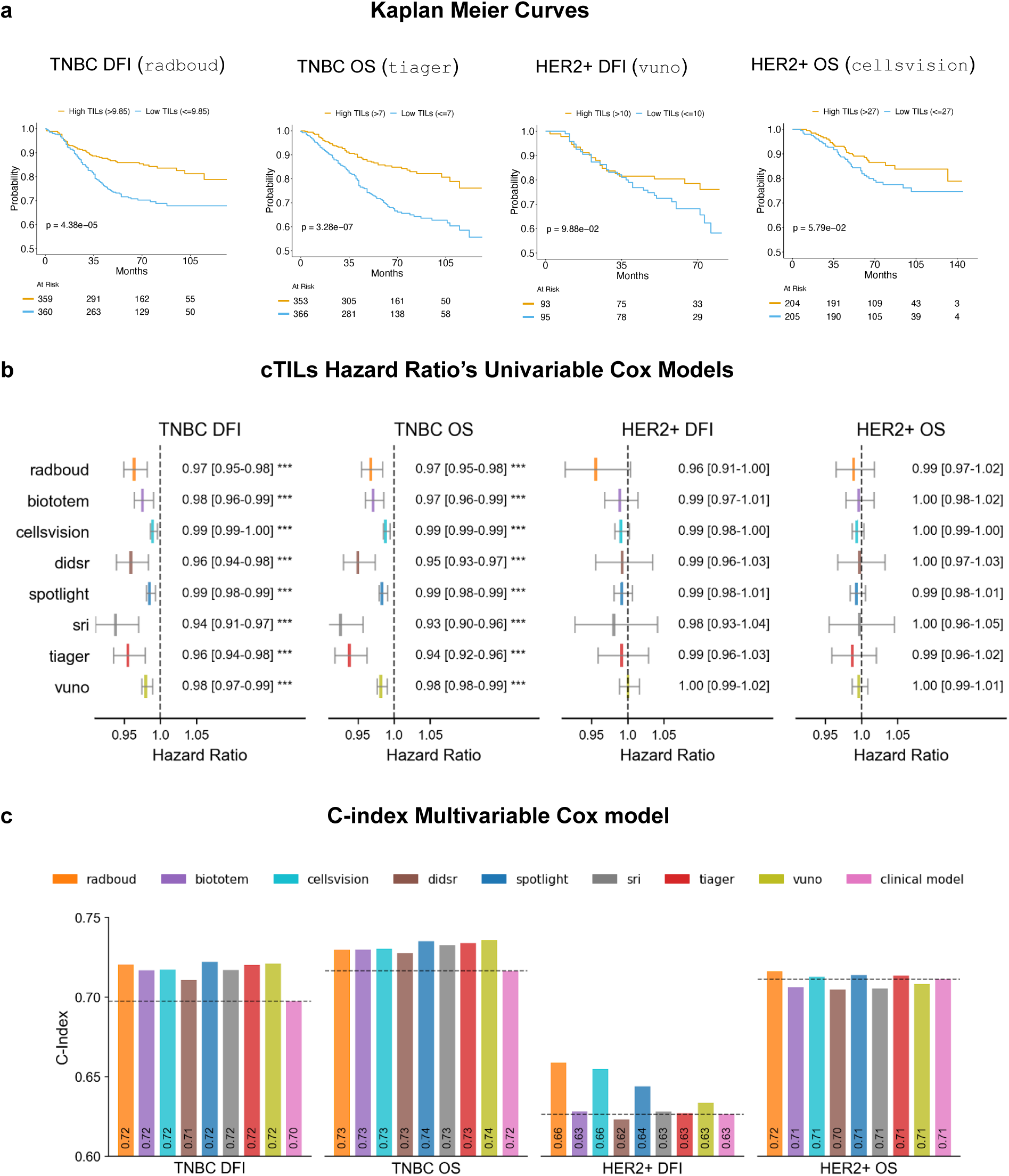
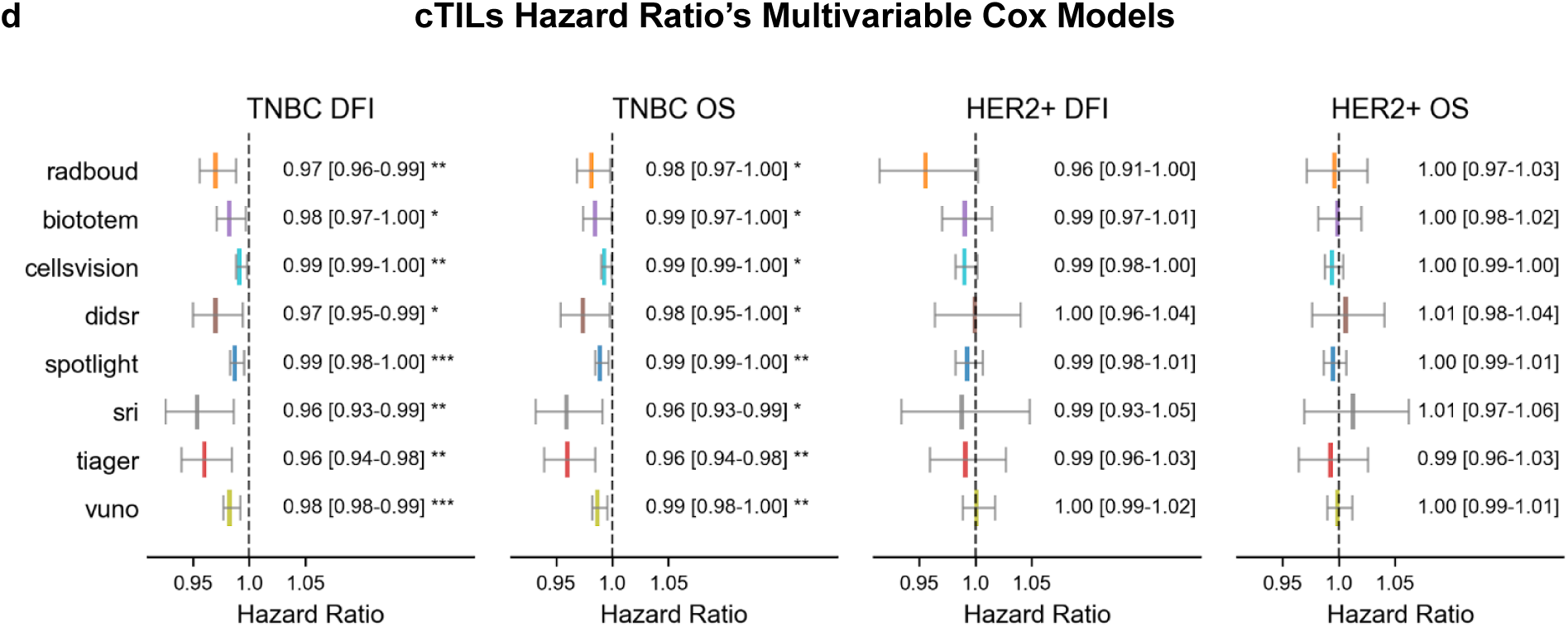
Prognostic value of cTILs as continuous values in resection samples. High and low sTILs are determined by the median cutoff value. **A**, Kaplan-Meier curves for cTIL models and survival outcomes in specific breast cancer subtypes: Radboud with DFI in TNBC; Tiager with OS in TNBC; VUNO with DFI in HER2+; and CellsVision with OS in HER2+. Each algorithm achieved the lowest p-value in its respective analysis. **B** Forest plots of multivariable Cox regression models indicating hazard ratio and confidence interval in the format HR [CI] using DFI and OS on TNBC as endpoints**. c**, Bar plots of C-indexes computed using the prediction of a multivariable Cox regression model using a clinical model and a cTIL score as input. Values are compared with the C-index of the clinical model as a predictor (see dashed line). The graphs show that AI models achieve a higher C-index score than the clinical baseline in TNBC cases, while only some AI models outperform the clinical baseline in HER2+ cases. **d**, Forest plots of multivariable Cox regression models indicating hazard ratio and confidence interval in the format HR [CI]. Plots show association for some AI models with DFI and OS on TNBC. Abbreviations: cTILs: Computational Tumor-Infiltrating Lymphocytes, TNBC: Triple-Negative Breast Cancer, DFI: Disease-Free Interval, HER2+: Human Epidermal Growth Factor Receptor 2 Positive, OS: Overall Survival. * p<0.05, ** p<0.01, *** p<0.001.

In multivariable Cox regression analyses, we first built a “clinical” model using commonly used clinicopathological variables as covariates: age, histopathological subtype, histopathological grade, molecular subtype, stage, surgical procedure and administration of adjuvant therapy. Second, we built a multivariable “enriched” model by using the output of the clinical model used as a predictor and the cTILs as a second variable (Method section *Statistical analysis*). For TNBC, all multivariable-enriched models achieved a C-index higher than the clinical model in both RES_DFI_ (C-index enriched model range [0.71-0.72], C-index clinical model=0.70) and RES_OS_ (C-index enriched model range [0.73-0.74], C-index clinical model=0.72) (Figure 4c). This shows the added value of cTILs in addition to commonly used clinical variables, also seen in the adjusted hazard ratios for the multivariable Cox analysis of continuous cTILs scores (Figure 4d). For HER2+, only some enriched models achieved C-index higher than the clinical model in RES_DFI_ (C-index clinical model=0.63, C-index enriched model range [0.62-0.66]) and RES_OS_ (C-index clinical model=0.71, C-index enriched model range [0.71-0.72]) but none of them significantly added value on top of clinical variables, as shown by the adjusted hazard ratios in Figure 4d.

#### Biopsies

For preoperative biopsies, we included 2,099 patients from the G6, G7, G8 and NKI cohorts (Method section *Materials*; Figure 1e) for which DFI, OS and vTILs information was available, and analyzed each cohort independently. We performed survival analyses considering both cTILs and vTILs as continuous values. See Supplementary Figure *Biopsies - Kaplan Meier Curves,* for an overview of survival curves.

Overall, we found that cTILs computed on biopsies did not show a strong consistent prognostic value across cohorts and endpoints. In univariable analysis of the TNBC subset within the G7 dataset, several cTIL models demonstrated significant associations with OS (`vuno`, `sri`, `didsr`, `cellsvision`, and `biototem`). Furthermore, vTILs were significantly associated with OS in both G6 and G7. For DFI, only `vuno` and vTILS were significantly associated in G7. Figure 5a shows an overview of all Hazard Ratios for the TNBC subset of G6 and G7 datasets. In the HER2+ subset, significant associations with OS were observed in the G7 dataset for the cTILS models `radboud`, `biototem`, and `sri`. Regarding DFI, significant associations were identified in the G6 dataset for `radboud`, `cellsvision`, `didsr`, `spotlight`, and `sri`. Additionally, vTILs showed significant associations with DFI exclusively in the G6 dataset. Figure 5b shows an overview of all Hazard Ratios for the HER2+ subset of G6 and G7 datasets. No significance was observed in the G8 and NKI datasets, nor for the TNBC and HER2+ subgroups in terms of OS or DFI for any cTILs algorithm or vTILs.

**Figure 5:**
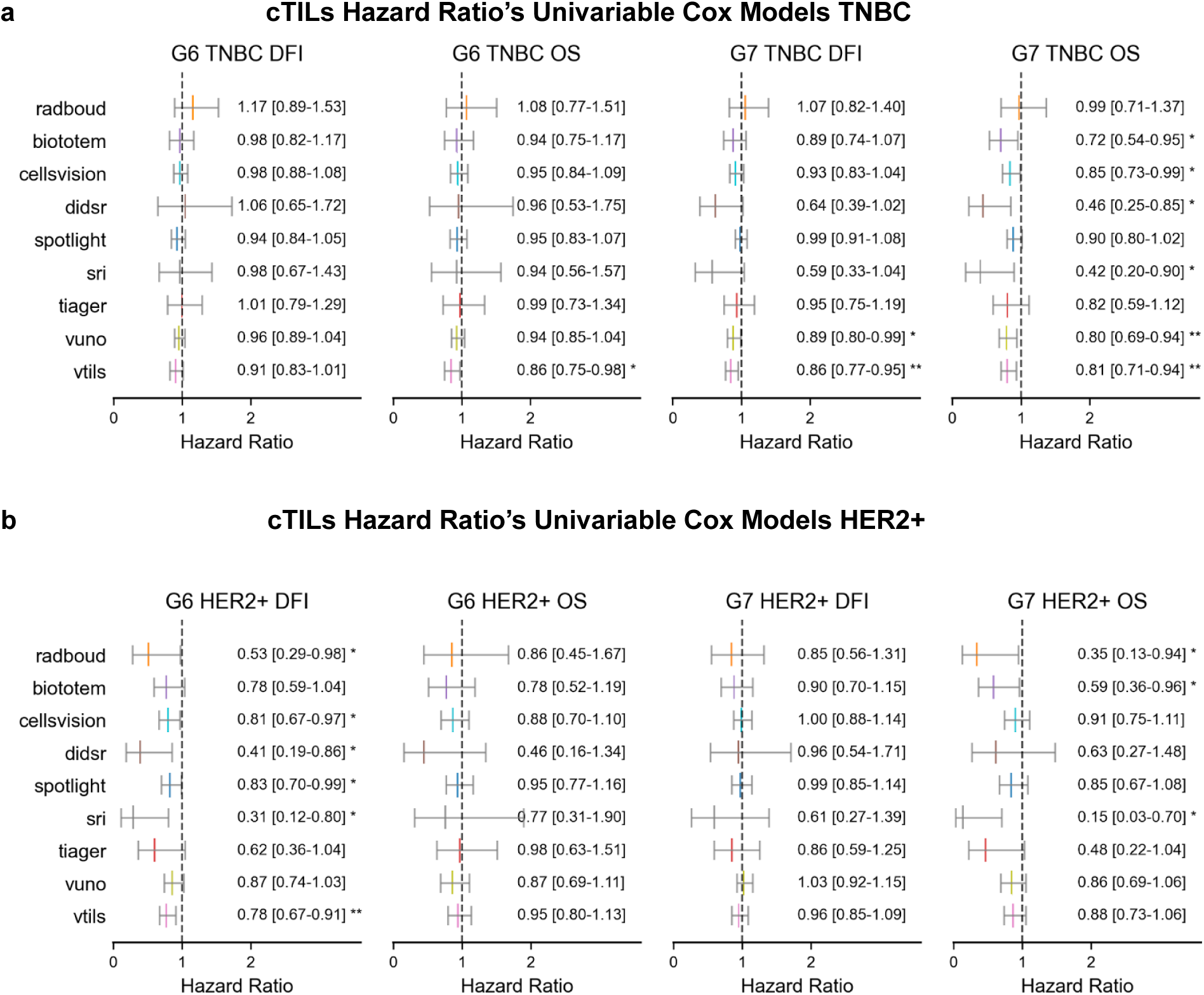
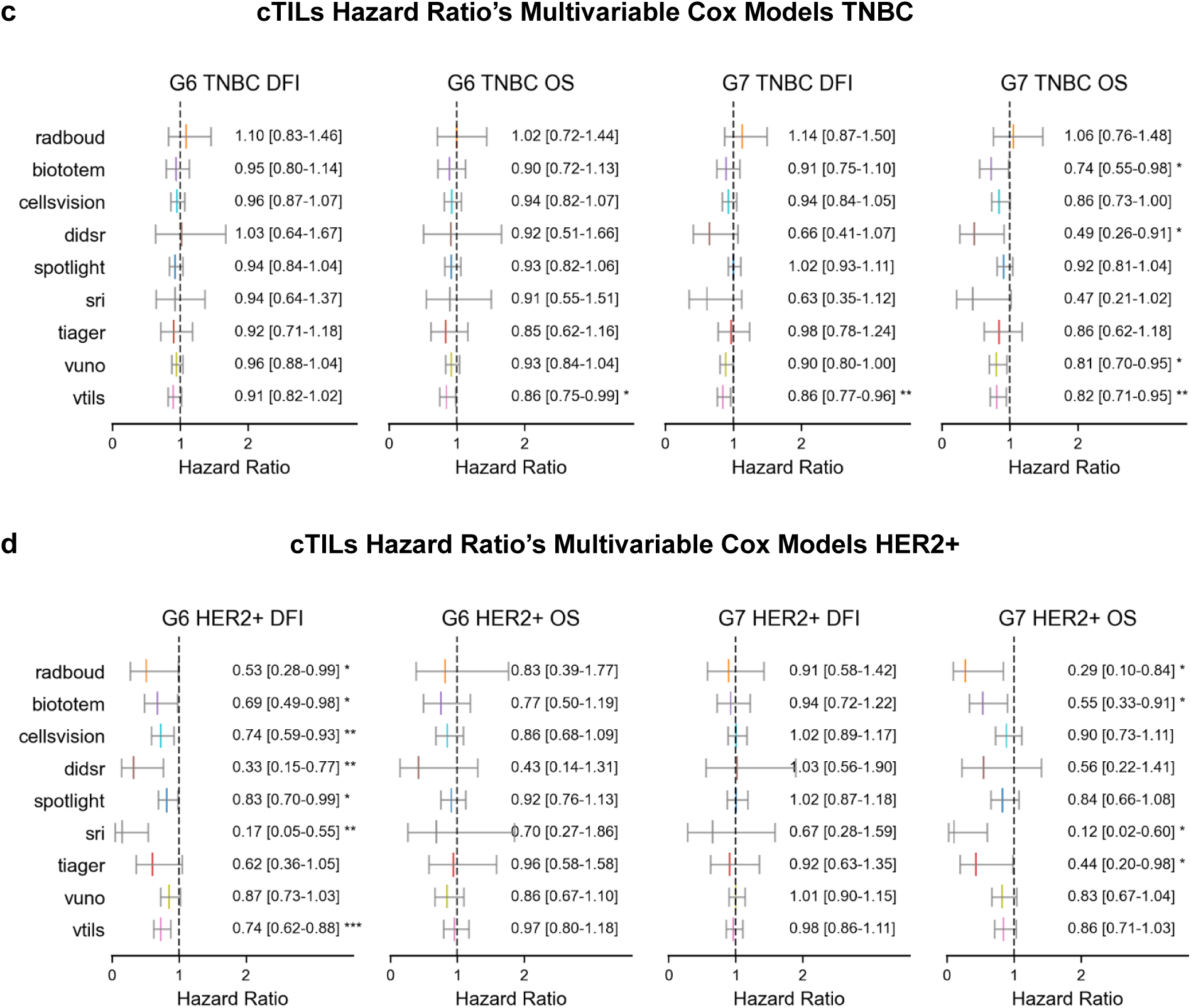
Prognostic value of cTILs in biopsy samples. **a**, Forest plot for Univariable Cox regression models for all cTILs and vTILs scores for TNBC biopsies. **b**, Forest plot for Univariable Cox regression models for all cTILs and vTILs scores for HER2+ biopsies. **c**, Forest plot for Multivariable Cox regression models for all cTILs and vTILs scores for TNBC biopsies. **d**, Forest plot for Multivariable Cox regression models for all cTILs and vTILs scores for HER2+ biopsies. * p<0.05, ** p<0.01, *** p<0.001.

Multivariable Cox analyses confirmed the significance of cTILs and vTILs observed in univariable analyses across cohorts and endpoints, except for `cellsvision` and `sri`, which were non-significant in G7 TNBC OS, `vuno` was not significant in G7 TNBC DFI, while `tiager` was significant in G7 HER2 OS, and `biototem` was significant in G6 HER2 DFI. Figure 5c,d show overviews of all cTILs Hazard Ratios for the TNBC and HER2+ subsets of the G6 and G7 datasets. See Supplementary Section: *Biopsies - Cox Proportional Hazards Analysis* for the uni- and multivariable analysis on the G8 and NKI datasets.

## Discussion

The TIGER challenge was designed with a “reproducibility-first” mindset, generating high-quality public sources for development and validation of AI models for cTILs quantification. These include a) multi-centric digital pathology data to train AI models (see *Data Availability* section), b) a public platform for benchmarking computer vision and prognostic performance of cTILs, c) open-source baseline models, including training and testing pipelines, d) and final AI models for cTILs quantification, available as stand-alone applications on the *Grand Challenge* platform and as open-source solutions with a permissive license (see *Code Availability* section).

The outputs of the TIGER challenge are the foundation of this study. Using the privacy-preserving Grand Challenge platform to benchmark AI models, we performed the first external validation of TIGER cTIL models on multi-centric data from clinical practice and phase-3 trials. We showed the strong Spearman rank correlation coefficient of most cTILs and vTILs (mean ρ value = 0.61, range [0.26-0.85]), the positive association of both cTILs and vTILs with response to neoadjuvant chemotherapy in HER2+ breast cancer (AUC cTILs range [0.52–0.78], vTILs range [0.55–0.65]), and the prognostic association of cTILs with both DFI and OS in TNBC surgical resections (multivariable C-Index range DFI [0.71,0.72], OS [0.73-0.74]). We also showed the poor association between cTILs and survival (DFI and OS) on HER2+ surgical resections and on preoperative biopsies. The cohorts used in TIGER as training and test data contained higher numbers of TNBC compared to HER2+ cases, as well as more surgical resection slides than biopsies. Although this information was not disclosed to the TIGER participants, these factors may have implicitly steered models’ optimization towards performance on TNBC resections during the experimental phase of the challenge. Access to larger training data sets containing more preoperative cases in the future could help balance model optimization and improve performance on biopsies.

While an accurate analysis of tissue morphology remains the base for robust cTILs scores, we found a discrepancy between methods achieving high computer vision performance (`tiager`, `biototem`, `spotlight`), cTILs showing strong correlation with pathologists (`biototem`, `sri`, `cellsvision`), and association with pCR and survival. We found that methods that automated the recommendations of the TILs WG^2^ reported Spearman rank correlation coefficient with vTILs ranging from weak (ρ=0.26) to very strong (ρ=0.85), especially for biopsies. This may indicate the importance of algorithmically defining the region where TILs are scored, since methods strongly relying on the definition of the “tumor bulk” (`radboud`, `tiger`, `spotlight`) or on peritumoral regions (`vuno`) showed lower correlation with visual TILs and lower association of cTILs with pCR computed on biopsies. This shows that the optimal way to engineer morphological spatial information into a computational biomarker may be different from the way it is defined for human readers and is still an active area of research.

In this context, our study provides insights for both researchers and clinicians. For researchers, we provide indications on how to develop cTILs methods, addressing the lack of consensus methods on cTILs design, fostering further development and validation of improved AI models for cTILs in the future. For clinicians, we introduced a way to obtain indirect access to models via Grand Challenge to test automated cTILs assessment on anonymous data for research use. Our results also suggest that cTILs could be a useful tool to assist pathologists in TILs quantification. Compared to visual TILs assessment, cTILs are more objective and reproducible, can provide visual feedback on tissue segmentation and lymphocyte detection and support decision making of pathologists with interpretable visual output. This may be particularly relevant in settings where no, or few, specialized pathologists are available.

The path towards clinical adoption of these solutions will require further validation on both retrospective and prospective data. In this context, the privacy-preserving mechanism for indirect access to data (i.e., without sharing data with AI developers) and the public benchmark built within TIGER have the potential to become an easily-accessible benchmark for validation of present and future cTILs methods, with potential value to support certification for regulatory purposes. Compared to TIGER, in this study we have broadened the statistical analysis across multiple cohorts and clinical endpoints (pCR, DFI, OS). Accordingly, we have expanded^4^ the TIGER web platform to include the additional statistical tests used in this study. Finally, international reader studies and research on the usability and impact of cTIL models are needed, to better understand their potential as a computer-aided supportive tool or as a basis for computational biomarkers.

This study has limitations. First, our validation was based on a mechanism of indirect access to data, which did not allow us to investigate the causes of differences in computer vision performance and predicted cTILs. Examples are segmentation outputs on specific regions of WSIs that play an important role in the tumor microenvironment, such as tertiary lymphoid structures (TLS), lymphoid stroma, perivascular invasion, and other confounders known to play a role in visual TIL scoring, which could not be fully explored. These aspects deserve a more detailed analysis and should be addressed in dedicated future studies.

Second, we analyzed raw cTILs values as produced by AI models, without performing any form of calibration. In the roadmap towards assessing the clinical applicability of computational biomarkers and their potential complementarity with visual scores, research on calibration of cTILs scores in line with what recently proposed by Arab et al.^34^ will be needed.

Third, the TIGER challenge instructed participants to produce models producing cTILs scores within a range between 0 and 100. However, each cTILs algorithm produced scores with a different distribution. As a consequence, it was not possible to dichotomize groups of patients using cut-off values such as 30%^7^ or 10%-60%^9^, used in previous studies. Therefore, in Kaplan-Meier analyses, we considered the median cTILs value per cohort as a cut-off point to dichotomize subgroups of patients. Next to calibration, future work will require the estimation of per-algorithm optimal cut-off points.

Fourth, this study did not identify a clear “winner” among the evaluated methods across all considered end points. While this leaves no standard digital assay for immediate broad adoption, our study provides indications on the validity of some cTILs methods on each of the considered end points based on the data at hand. One of the future directions of AI-based TIL measurement may lie in exploring combinations of approaches to develop an effective solution.

Finally, we limited the analysis to the association of cTILs with response to neoadjuvant chemotherapy. Given the increasing use of immune checkpoint inhibitors in combination with chemotherapy in early-stage TNBC, future development and validation of AI-based TILs should extend to treatment regimens including immune-checkpoint inhibitors. Moreover, given the previously demonstrated excellent long-term clinical outcomes of patients with anatomical low-stage TNBC and high TILs, even in the absence of chemotherapy, the potential for AI-based TIL measures to select patients for de-intensification is a compelling application that requires dedicated prospective evaluation.

## Conclusion

We have presented the analysis of several cTIL models on the largest multi-centric cohort of digital pathology whole-slide images of breast cancer patients. We have reported benchmarks on image analysis performance of each method, showed the agreement of cTILs with panels of pathologists, the positive association of cTILs with response after neoadjuvant therapy, and revealed a trend of achieving higher AUC values than pathologists in HER2+ cases. We showed that cTILs are associated with survival both as a stand-alone biomarker and by adding independent information to clinical variables in multivariable analyses in surgically resected TNBC, but not in HER2-positive disease. Finally, we showed that cTILs on biopsies did not show a strong consistent prognostic value across cohorts and endpoints. All methods tested in this work originated in an open community-driven project based on the TIGER challenge. To the best of our knowledge, this pioneers a model for public development and validation of cTIL models with artificial intelligence, which could serve as a blueprint for investigating other computational biomarkers, beyond the TILs and breast cancer.

## Online Methods

### Materials

In this section, we introduce all cohorts involved in *cTILs benchmarking* in this study and report the type of annotations available for each cohort. We refer to the Supplementary material for TIGER training data and computer vision benchmark data, including the re-staining study. All slides were stained with H&E, scanned in the origin center with whole-slide image scanners locally available and processed to standard TIFF at 20X magnification (0.5 μm/px) for this study.

### Definition of clinical outcome

#### Pathological complete response

In all cohorts, pCR was based on the local histopathological analysis of the resection specimen after neoadjuvant chemotherapy and indicated the absence of invasive cancer in the breast and axillary nodes, which is our primary target. GBG and SCDC defined pCR as ypT0/ypN0 (i.e., absence of in-situ cancer) but using different scoring systems (see Data for details); NKI and DIGITILS defined pCR as ypT0/is ypN0 (irrespective of ductal carcinoma in-situ as this information was not available).

#### Survival

We used the following definitions of survival data: disease-free interval (DFI), defined as the time in months between diagnosis (for clinical cases) or randomization (for clinical trials) and the date of clinically and/or pathologically detected (loco)regional or distant recurrence of BC. If no recurrence occurred, patients were censored at the date of last follow-up, death, or secondary malignancy. Overall survival (OS) is defined as the time in months between date of diagnosis of BC and date of death (independent of cause) or the date of last follow up (right-censored).

### Data

#### RUMC-SURV_TNBC_

This cohort consisted of n=597 TNBC cases (stage 1-3), derived from a previous study^35^. In brief, TNBC cases diagnosed between 2006 and 2014 in five hospitals from Eastern Netherlands (Radboudumc, Nijmegen; Canisius-Wilhelmina Hospital, Nijmegen; Jeroen Bosch Hospital, ‘s-Hertogenbosch; Bernhoven Hospital, Uden and Hospital Pantein, Boxmeer) were included. At the time of inclusion, patients had not received neoadjuvant chemotherapy and did not have prior history of breast cancer. All tissue blocks were collected centrally and cut and stained in batches in the pathology laboratory of the Radboudumc. For this reason, although patient data is multi-centric, we refer to this cohort as the RUMC-SURV_TNBC_ cohort. All slides were scanned with a Pannoramic 1000 DX scanner (3DHistech) at 40X magnification (0.24 um/px). For each patient, the following clinicopathological information was available: age, histological subtype, grade, molecular subtype, stage, surgery, adjuvant therapy. For this cohort, DFI and OS end-points were available, visual assessment of stromal TILs (sTILs) was not available.

#### RUMC-SURV_HER2+_

This cohort consisted of n=221 HER2+ cases (stage 1-3) collected between 2006 and 2013 at two Dutch hospitals (Radboudumc, Nijmegen; Canisius-Wilhelmina Hospital, Nijmegen). As for RUMC-SURV_TNBC_, all tissue blocks were collected, cut, stained and scanned at Radboudumc; therefore, we refer to this cohort as the RUMC-SURV_HER2+_. Patients did not get any neoadjuvant treatment, did not have multiple breast tumors at the time of diagnosis nor bilateral breast cancer, did not have multiple diagnosis of breast or other invasive tumors within 5 years, and all of them received treatment. HER2+ status was based on immunohistochemistry (3+ score) and/or FISH (amplification). Scanner settings and clinicopathological information were the same as RUMC-SURV_TNBC_. For this cohort, only OS follow-up data was available, sTILs assessment was not available.

#### FinHER

This cohort consisted of n=303 cases from the FinHer trial^36^. Slides were stained with H&E and scanned at the Jules Bordet institute (Belgium) using a NanoZoomer 2.0-RS C10730 series (Hamamatsu) at a resolution of 0.23 µm/px. From the original FinHer cohort we selected TNBC (n=120) and HER2+ (n=183) cases, after excluding cases with incomplete data (e.g., no match between image data and clinical data) and cases with stage 4 disease at initial presentation. Both HR status and HER2 expression were determined by immunohistochemistry according to each institution’s guidelines. When HER2 expression was considered positive in immunohistochemistry (either 2 or 3 on a scale from 0 to 3), gene amplification status was determined centrally using CISH. Cancers with six or more gene copies were considered HER2 positive. The same clinicopathological information as RUMC-SURV_TNBC_ and RUMC-SURV_HER2_ were available. DFI and OS data were available as endpoints, as well as visual sTIL assessment scored by two breast pathologists (RS, NS).

#### GBG

We included a set of n=1928 patients with primary breast cancer who were treated with neoadjuvant combination chemotherapy from three randomized trials done by the German Breast Cancer Group (GBG), namely GeparSixto^37^ (G6), GeparSepto^38^ (G7), GeparOcto^39^ (G8). All patients underwent core-needle biopsy prior to breast cancer diagnosis and subsequent neoadjuvant chemotherapy treatment. We included n=553 cases from G6 (297 TNBC; 256 HER2+), n=617 from G7 (265 TNBC; 352 HER2+) and n=758 from G8 (390 TNBC, 368 HER2+). Slides were stained and scanned at the Institute of Pathology, Philipps-University Marburg, University Hospital UKGM Marburg using either a Leica Aperio AT2, a Sysmex/3DHitech’s Pannoramic 250 FLASH III and Pannoramic SCAN II or a NanoZoomer 2.0-HAT from Hamamatsu. The following clinicopathological variables were available for these cohorts assessed at baseline: pathological grading, molecular subtype, calculated on the basis of HR (ER, PR) and HER2 status, Ki67 value, tumor size (T stage) and nodal status (N stage). For these cohorts, DFI and OS data were available as endpoints; pCR was defined as ypT0/ypN0, sTILs were available and scored as described in^9^.

#### DIGITILS

We included a set of n=77 cases from the Cliniques Universitaires Saint-Luc (Brussels, Belgium), n=34 TNBC and n=43 HER2+. All patients underwent core-needle biopsy prior to breast cancer diagnosis and subsequent neoadjuvant chemotherapy treatment. The TNBC part of this cohort belongs to the IVITA study^6^, involving a reader study with n=40 pathologists per slide that assessed sTILs. The HER2+ part belongs to a recent study^40^, involving a reader study with n=3 pathologists per slide. All slides were scanned at Saint Luc Hospital using a Hamamatsu Nanozoomer 2.0RS scanner at 40X magnification. For each patient, available information contained tumor subtype, tumor grade, and response to neoadjuvant chemotherapy scored based on the Residual Cancer Burden (RCB) scoring system (RCB=0 corresponds to pCR defined as ypT0/is ypN0).

#### NKI

We included a set of n=218 TNBC routine diagnostics cases from the Dutch Cancer Institute (Nederland Kanker Instituut, NKI; Amsterdam, Netherlands). All patients underwent core-needle biopsy prior to breast cancer diagnosis and subsequent neoadjuvant chemotherapy treatment. All slides were stained at the NKI and scanned with an Aperio AT2 (Leica Biosystems) at 40X magnification. The same clinicopathological variables used in GBG were available; DFI and OS data were available as end points; pCR was defined as ypT0/is ypN0, meaning absence of invasive cancer in breast and axillary nodes irrespective of ductal carcinoma in-situ as this information was not available; sTILs were scored by two pathologists following the recommendations of the TIL WG, as described in^41^.

#### SCDC

We included n=56 cases (15 TNBC, 41 HER2+) from the IRCCS Sacro Cuore Don Calabria Hospital (SCDC, Verona, Italy). All slides are diagnostic biopsies stained with H&E, extracted via core-needle procedure (before chemotherapy), and scanned with a Ventana DP 200 slide scanner at 20X magnification. The same clinical variables and clinical outcomes included in the GBG and NKI cohorts were available, with the exception of T stage. However, due to the limited number of events (n=11 recurrence, n=6 death) and the high rate of censored cases (n=41 (73%), for no recurrence and no death), we only considered this cohort for the correlation with pathologists and the prediction of pCR. pCR was derived from the PINDER scoring system^42^ (PINDER=1i for breast and PINDER=1+2 for lymph nodes). sTILs were scored by one pathologist (EM) following the recommendations of the TIL WG.

## Statistical analysis

### Computer vision performance

We benchmarked tissue segmentation and lymphocyte detection with the same approach used in the computer vision track of TIGER.

#### Segmentation

For tissue segmentation, we focused on two classes that play a central role in the definition of the TILs score and computed an overall Dice score as the average of the Dice score for tumor and the Dice score for tumor-associated stroma. We computed these scores over all slides by taking the target class (either tumor or tumor-associated stroma, respectively) as foreground and rest (including tumor-associated stroma or tumor, respectively).

#### Detection

For lymphocyte detection, we performed a Free Response Operating Characteristic (FROC) analysis, computing sensitivity (true positive rate, TPR) versus average false positives (FP) per mm² over all slides. Given the i-th predicted cell locations *c_p,i_* = (*x_p,i_*,*y_p,i_*) of lymphocytes and plasma cells, each with likelihood *l_i_*, and the j-th manual annotation *c_m,j_* = (*x_m,j_*,*y_m,j_*), we considered a hit of manual annotation *c_m,j_* if *l_i_* > τ and if *d*(*c_p,i_*, *c_m,j_*) ≤*D*, where τ is a threshold and *d*(•) is the Euclidean distance. In our case, τ assumed all likelihood values predicted by each algorithm on the entire test set, and D=4um, based on previous work. For each threshold τ, we obtained values for the true positives (TPs), false negatives (FNs), and false positives (FPs) and used those to build an FROC curve. From the curve, we derived a “FROC score” by averaging sensitivity computed at five pre-selected values of FP/mm²: (10, 20, 50, 100, 200, 300).

### Combination of sTILs

In several cohorts (NKI, DIGITILS, GBG, FinHER, SCDC), one or more pathologists visually assessed the stromal TILs according to recommendations of the TIL working group. The analysis of the inter-observer variability is out of scope for this study because it was addressed in previous studies, including the ones in which the used cohorts were originally introduced^6,36–40^. In this study, following^6^, we combined the multiple sTILs scores at slide level into a single score by computing the *median* sTILs score. Note that a) this will result in the mean sTILs value when only two pathologists were involved; b) we combined the sTILs score without any transformation, because following the TILs WG recommendations, each score indicates the percentage of stroma covered by the TILs in the tumor bulk, therefore producing a value between 0 and 100. We refer to this combined score as the pathologist visual TILs (vTILs). When multiple slides per patient were available, one representative slide was selected being the one with the highest mean sTILs score.

### Correlation with vTILs

We measured the correlation between each cTIL score and the vTILs. We report correlation metrics in terms of Spearman rank correlation coefficient and via scatter plots for each pair of TIL scores (see Figure 2c). Possible discretized pathologist scores and cTILs distributions not necessarily aligned with the ones of visual scoring make the Spearman rank correlation coefficient appropriate for capturing monotonic relationships between cTIL and vTIL scores.

### Treatment response: prediction of pCR

Based on previous studies, we assume that high TILs predict pCR while low TILs predict no pCR. For cohorts of pre-treatment biopsies of patients that received neoadjuvant chemotherapy (NKI, GBG, DIGITILS, SCDC), we performed Receiver Operating Characteristic (ROC) curve analysis for all cTILs and vTILs using pCR as a target. From each ROC curve, we computed the area under the curve (AUC) as the performance metric as well as its confidence interval. We also computed a one-sided Wald p-value from a normal distribution approximation. No adjustments were made for testing multiple hypotheses.

### Prediction of survival

Based on previous studies, we assumed that high TILs predict longer survival and low TILs predict a higher risk of an event. We tested this hypothesis using cTILs from all algorithms and vTILs on cohorts with survival data available (NKI, GBG, FinHER, RUMC-SURV_TNBC_, RUMC-SURV_HER2+_). We detail here the tests that we performed on cohorts containing surgical resections and biopsies. Partly derived from the survival task of TIGER, we redesigned the survival analysis performed in this study to consider additional tests and endpoints. No adjustments were made for testing multiple hypotheses.

#### Prediction based on surgical resections

For surgical resections, we tested the prognostic value of cTILs in addition to the clinical variables that we considered in the TIGER challenge, namely molecular subtype, age, morphological subtype, grade, stage, adjuvant treatment. We did it in three steps. First, we pooled together all cases from FinHER, RUMC-SURV_TNBC_ and RUMC-SURV_HER2+_, resulting in a cohort of n=1128 cases, which we called ALL-SURV. Second, we built a multivariate Cox regression by fitting the model on all clinical variables of ALL-SURV and we save this model and turn it into a *predictor*, which we refer to as the “clinical model”, to use it in downstream tasks. Third, for each algorithm, we updated the model by replacing all the previously described clinical variables with the predictor, which was common to all algorithms, and enriched it with cTILs as the second variable in the updated multivariable Cox regression model; we refer to this model as the “enriched model”. Since multivariable analysis was repeated for each AI algorithm, the predictor allows for clinical variables to be accounted for in each model in a fixed fashion, thus allowing for comparisons of the prognostic value of cTILs across models. We also used the output of the enriched model to compute the C-index using either DFI or OS as end points. For this, we used the method proposed by Harrel^43^, commonly used in the literature. Note that this is different from the C-index implementation adopted during the TIGER challenge, which was based on the Uno’s AUC approach (Supplementary section *Survival track evaluation*). Univariable analysis of cTILs is done via Kaplan-Meier curves analysis, and p-values computed via the log-rank test. Hazard ratios depicted in the forest plots were obtained from univariable Cox models. In our analysis, we considered TILs scores as a continuous value and P-values for the hazard ratios were derived from the Wald test.

#### Prediction based on pretreatment biopsies

For biopsies, we assessed the prognostic value of cTILs on OS and DFI in a univariable setting (Kaplan Meier curves and Cox regression model) and multivariable setting (Cox regression model). In Kaplan Meier curves, for each algorithm, categories high and low were dichotomized based on the data set median analyzed in the respective section, namely G6, G7, G8 or NKI. For each endpoint and for each algorithm (and vTIL) we calculated the cTIL hazard ratio (or vTIL, respectively) from a univariable and multivariable (with covariates: molecular subtype, grade, stage (T, N), TIL score) Cox regression model that was run over each biopsy cohort in which survival data was available. Corresponding cTILs were handled as a continuous variable in the Cox regression models. P-values for the hazard ratios were derived from the Wald test.

## Data Availability

Information about data used in the TIGER challenge can be found on the TIGER website (https://tiger.grand-challenge.org/Data/). TIGER training data is publicly available via AWS Open Data Registry (https://registry.opendata.aws/tiger/), which contains the full set of training slides for the WSIROI, WSIBULK and WSITILS subsets of training data, including their manual annotations. A simplified version of training data, solely containing regions of interest from the WSIROI subset and their manual annotations is available via Zenodo (https://zenodo.org/records/6014422)44. Sequestered test sets TIGER-CV and TIGER-SURV (subset of the survival data used in the TIGER challenge) are available via indirect access by submitting AI models through the Grand Challenge platform: (https://tiger.grand-challenge.org/evaluation/segmentation-and-detecton-final/submissions/create/ for TIGER-CV), (https://tiger.grand-challenge.org/evaluation/survival-final-evaluation/submissions/create/ for TIGER-SURV). Data from GBG, NKI, DIGITILS, SCDC need to be requested to the respective clinical trial groups or centers

https://zenodo.org/records/6014422

https://registry.opendata.aws/tiger/

## Extended Data Figures

**Table 2:** Table-overview-external-cohorts. Overview of cohorts used for the external validation, including details on data type, scanner, magnification, spacing (in micron/px), country and whether the data comes from daily clinical practice or from clinical trials. Magnification indicates the one used originally to scan the slides, all WSI were then processed at 0.5 um/px in this work. RUMC: Radboud University Medical Center; GBG: German Breast Group; NKI: Nederland Kanker Instituut (Dutch Cancer Institute); SCDC: Sacro Cuore Don Calabria hospital.

| Cohort | Tissue | Scanner | Magnification | micron/px | Country | Source |
| --- | --- | --- | --- | --- | --- | --- |
| RUMC-SURV | resections | Pannoramic 1000 DX | 40X | 0.242 | Netherlands | clinic |
| FinHER | resections | NanoZoomer 2.0-RS C10730 | 40X | 0.228 | Finland | trial |
| GBG<br>(G6, G7, G8) | biopsies | Leica Aperio AT2 Sysmex<br>NanoZoomer 2.0-HAT | 20X | 0.503 | Germany | trial |
|  |  | Pannoramic 250 FLASH III<br>Pannoramic SCAN II | 40X | 0.242 |  |  |
| NKI | biopsies | Aperio AT2 | 40X | 0.252 | Netherlands | trial |
| DIGITILS | biopsies | Nanozoomer 2.0RS | 40X | 0.227 | Belgium | clinic |
| SCDC | biopsies | Ventana DP 200 | 20X | 0.465 | Italy | clinic |

## Data availability

Information about data used in the TIGER challenge can be found on the TIGER website (https://tiger.grand-challenge.org/Data/). TIGER training data is publicly available via AWS Open Data Registry (https://registry.opendata.aws/tiger/), which contains the full set of training slides for the WSIROI, WSIBULK and WSITILS subsets of training data, including their manual annotations. A simplified version of training data, solely containing regions of interest from the WSIROI subset and their manual annotations is available via Zenodo (https://zenodo.org/records/6014422)^44^. Sequestered test sets TIGER-CV and TIGER-SURV (subset of the survival data used in the TIGER challenge) are available via indirect access by submitting AI models through the Grand Challenge platform: (https://tiger.grand-challenge.org/evaluation/segmentation-and-detecton-final/submissions/create/ for TIGER-CV), (https://tiger.grand-challenge.org/evaluation/survival-final-evaluation/submissions/create/ for TIGER-SURV). Data from GBG, NKI, DIGITILS, SCDC need to be requested to the respective clinical trial groups or centers.

## Code availability

Information about code used in the TIGER challenge is available on the TIGER website (https://tiger.grand-challenge.org/Code/). The code of the baseline TIGER algorithm developed by the Computational Pathology Group of Radboudumc is available at https://github.com/DIAGNijmegen/pathology-tiger-baseline. An example of making a docker container to upload an algorithm to the TIGER platform on Grand Challenge is available at https://github.com/DIAGNijmegen/pathology-tiger-algorithm-example. TIGER evaluation metrics are available at https://github.com/DIAGNijmegen/pathology-tiger-algorithm-example/tree/main/evaluations, including code to benchmark computer vision and survival performance. The WholeSlideData packages used to read and write WSIs data within TIGER are available at https://github.com/DIAGNijmegen/pathology-whole-slide-data. The source code and binaries of the ASAP viewer are available at https://github.com/computationalpathologygroup/ASAP. The Grand Challenge platform is an open-source code, available at https://github.com/comic/grand-challenge.org/. All TIGER models are publicly available under permissive licenses and are available at:

- aivis (https://github.com/AIVIS-MING/TIGER_SEG-DET)
- biototem (https://github.com/biototem/TIGER_challenge_2022)
- cellsvision (https://github.com/XulinChen/Algorithm-for-Tiger-Challenge)
- didsr (https://github.com/DIDSR/DIDSR-TiGER)
- radboud (https://github.com/DIAGNijmegen/pathology-tiger-baseline)
- spotlight (https://github.com/Spotlight-Pathology/spotlight-tiger)
- sri (https://github.com/Vishwesh4/TigerSubmission)
- tiger (https://github.com/adamshephard/TIAger)
- vuno (https://github.com/vuno/tiger_challenge).

## Author contributions

MvR, WA and LT co-designed the study, prepared and curated the data and the platform of the TIGER challenge, conducted the TIGER challenge and the post-challenge analysis, and wrote this manuscript. FC, JvdL and RS co-designed the TIGER study. FC supervised the TIGER study, designed the post-challenge study and wrote this manuscript. RS also scored visual TILs across several cohorts, including the WSITILS dataset of TIGER. MB and JB contributed to data collection and annotations, and provided scientific input during the design phase of the TIGER challenge. DD and SM provided advice on the statistical analysis during the TIGER challenge. LCB, DP, EISS, A-VL and HH provided manual annotations of data used in the TIGER training and test data, as part of the International TILs working group. LC and DL provided data from the Jules Bordet cohort. MAT and LC provided data from the BCSS and NuCLS studies, and advice on remapping existing labels to TIGER labels. CdK developed the TIGER baseline algorithm at Radboudumc under the supervision of MvR, WA and FC. VD provided support to data collection and curation at Radboudumc. JL and NW provided access to the HistokatFusion registration software and supported its use in this study. MvB and CG designed and ran the two clinical studies part of the DIGITILS cohort, curated the data and the annotations via reader studies. EL and JW designed the study on the NKI cohort, HH scored the TILs in the NKI cohort, LM contributed to slide collection and clinical data at the NKI, SvdB contributed to statistical analyses. CD initiated and co-organized the GBG original project, managed the biobank, and supplied the G6-8 samples/images. PJ produced digital slides of the GBG cohort. Karsten Weber developed the statistical analysis plan of the analysis of biopsies in this study and implemented statistical tests. EM and GB collected and curated SCDC data; EM also scored visual sTILs. JDD, HJ and SL provided access to the FinHER data. RD and SM co-developed the statistical analysis plan of the analysis of resections in this study and implemented statistical tests. JM, MG and AM developed the Grand Challenge functionalities needed to run the TIGER challenge and provided technical support on behalf of the Research Software Engineers team at Radboudumc. AL and CR enabled running the TIGER challenge on Grand Challenge by sponsoring the project, and by hosting the training dataset on AWS Open Registry. BvG supervised the developments on Grand Challenge and AWS. All authors have contributed to writing and reviewing this manuscript and approve it.

## Inclusion and ethics

The requirement for ethical approval to use data from the RUMC-TNBC and RUMC-HER2+ cohorts was waived by the institutional review board (case number 2015–1711) of the Radboud University Medical Center (Radboudumc). For the FinHer trial, an ethics committee at Helsinki University Hospital approved the study and study participants provided written informed consent (trial identifier: ISRCTN76560285). For DIGITILS, the institutional ethics committee approved this study (file number: RETRO-TNBC-15-2019/03JUL/297 for the TNBC cohort, file name: RETRO-HER2-15 for the HER2+ cohort). The approval to use slides from the GBG was given at two institutions: Charité ethic committee approval number: EA1/139/05; Philipps-University Marburg ethic committee approval number: 38/20. The use of the slides from SCDC for the study was approved by the Ethics Committee for Clinical Research of the Provinces of Verona and Rovigo under number 25046. The use of the slides from NKI for the study was approved by the institutional review board of the Netherlands Cancer Institute under number CFMPB737.

## Acknowledgments

The authors would like to thank Arian Arab, Weijie Chen, Victor Garcia, Nicholas Petrick and Brandon Gallas for their contribution to the TIGER challenge by implementing the `didsr` algorithm, Fieke Mooren for producing the re-stained slides at Radboudumc, Meyke Hermens for her initial support to data collection at Radboudumc, Nicolas Sirtaine for his contribution to the visual TILs scoring, Yat-Hee Liu for her contribution to NKI data collection, Michela Campora and Carlijn Lems for their valuable feedback on the final drafts of this manuscript. We acknowledge the staff of the NKI Core Facility Molecular Pathology & Biobanking (CFMPB) for providing the tissue samples.

## Footnotes

1 `aivis` only made valid submissions to the computer vision track, therefore it was not included in the cTILs benchmark.

2 The `radboud` algorithm was developed before TIGER and proposed by the challenge organizers as the *baseline* solution to improve upon within TIGER and did not officially compete in the challenge; it is reported here for completeness.

3 `sri` only made submissions to the survival track, therefore it was not included in the computer vision benchmark.

4 We will update the evaluation procedure on the TIGER website upon acceptance of this manuscript.

