## Supplementary Information for "Tumor-infiltrating lymphocytes in breast cancer through artificial intelligence: biomarker analysis from the results of the TIGER challenge"

#### TIGER challenge design

Between January and June 2022 (see Supplementary Figure 1), we ran the TIGER challenge, hosted on Grand Challenge[1], a platform for machine-learning solutions in biomedical imaging, which supported us in hosting sequestered test data and allowed for the application of algorithms and evaluations. To participate in the challenge, teams (either individuals or groups of people) were asked to develop AI-based algorithms to 1) segment tissue compartments of tumor and stroma and detect lymphocytes in H&E-stained breast cancer digital pathology slides, and 2) compute a computational TILs score based on the spatial analysis of (1). No constraints were made for the design of methods in (1) and (2), for example, for designing a TILs score, teams could base their algorithms on the TIL Working Group's recommendation but were free and encouraged to develop new strategies. The only requirement was that the score produced in (2) was related to TILs and within a range of values [0,100]. In practice, teams were asked to create a dockerized algorithm and submit it to Grand Challenge, to be automatically applied and evaluated on the sequestered test data, giving participants only indirect access to the data.

We structured the TIGER as a two-tracks/two-phases challenge. In the *computer vision track*, we assessed the computer vision performance of submitted algorithms by evaluating the detection of lymphocytes and plasma cells and the segmentation of invasive tumors and tumor-associated stroma (see Computer Vision track in Evaluation section for details), and combined these scores to build a “segmentation and detection” leaderboard. In this track, the evaluation was only done in densely annotated region-of-interest (ROI), therefore submitted algorithms should expect a whole-slide image accompanied by a ROI mask as input to process. In the *survival track*, we assessed the prognostic value of automated TILs scores. Only the TILs scores from the algorithms were evaluated, but algorithms were expected to use the output of segmentation and detection to find the TILs. In this track, algorithms should expect a whole-slide image accompanied by a tissue mask, and process the entire area of the tissue mask. To fulfill requirements on computational cost in this track, we excluded from the tissue mask regions containing adipose tissue, which are not relevant in the assessment of TILs while occupying a substantial portion of tissue in the slide. Given a single TILs score per slide, we computed its prognostic value targeting DFI and computing the C-index of the output of a multivariable Cox regression model using the TILs as a covariate, next to a set of clinico-pathological variables (see Survival track in Evaluation section for details).

We defined two phases in TIGER. The *experimental phase* was run in parallel to development of AI algorithms. During this phase, participants could upload and test new and updated versions of their algorithms and visualize the results on both leaderboards. In this phase, results were computed on experimental test sets, and the number of submissions per week was limited to 3 for the computer vision track and 1 for the survival track. At the end of the experimental phase, we opened the *final phase*, where participants could choose only one algorithm, which was then applied and evaluated on the final sequestered test sets of each track *only once*. The results of the leaderboards in the final phase were also the final results of the TIGER challenge (see Supplementary Tables 4 and 5).

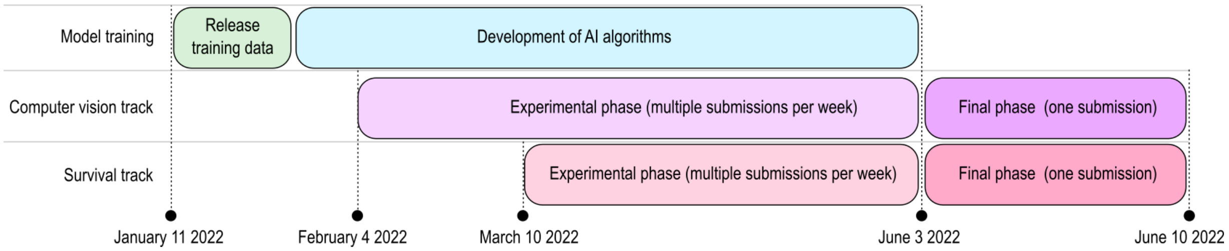


**Supplementary Figure 1:** Schematic representation of the timeline of the TIGER challenge.

### Building annotated data for TIGER

In this section, we describe the sets of whole-slide images collected and annotated to build the publicly available TIGER training dataset, as well as the sequestered test sets of the computer vision track, both for the experimental and the final phases. Supplementary Table 1 gives an overview of the data sources and their distribution across datasets.

#### RUMC-CV

This cohort consisted of n=139 whole-slide images from 139 patients (one slide per patient) from clinical cases acquired at the Radboud University Medical Center (RUMC, Nijmegen, Netherlands). Slides were selected coming from TNBC or HER2+ breast cancers, with tissue (both biopsies and resections) acquired at the time when patients were untreated systemically (e.g., no neoadjuvant therapy for patients whose we included the surgical resection specimen). Care was taken to have an approximately equal amount of biopsies and resections. As a result, we obtained 103 biopsies and 36 resections (88 HER2+, 51 TNBC); 117 invasive NST breast cancers, 9 apocrine carcinoma, 1 neuroendocrine carcinoma, 2 malignant adenomyoepithelioma, 3 micropapillary carcinoma, and 7 lobular carcinomas. All slides were scanned at RUMC with a Pannoramic 1000 DX (3DHistech) at a resolution of 0.25 μm/px; no additional clinical data or follow-up was available. This data was used in the TIGER training data and in the test data for the computer vision track, therefore we named it as RUMC-CV (to distinguish it from the RUMC-SURV cohorts, used for survival analysis).

## JB

This cohort consisted of n=99 whole-slide images from 99 patients (one slide per patient) from clinical cases acquired at the Jules Bordet (JB) institute (Belgium) from the period 2016-2020. As done for RUMC, slides were selected coming from TNBC or HER2+ breast cancers, with tissue (both biopsies and resections) acquired at the time when patients were untreated systemically, with an approximately equal amount of biopsies and resections. As a result, we obtained 52 biopsies and 47 resections, 31 HER2+ and 68 TNBC; 90 invasive NST breast cancers and 10 lobular carcinomas. All slides were scanned at JB with a NanoZoomer 2.0-RS C10730 series (Hamamatsu) at a resolution of 0.23 μm/px. No additional clinical data or follow-up was available. Similar to RUMC data, data from this cohort was used partly as TIGER training data and partly as test data for the computer vision track.

### TCGA

We included a subset of cases from the publicly available TCGA-BRCA cohort[2]. We included the n=151 TNBC cases (TCGA-TNBC) that were part of the existing BCSS[3] and NuCLS projects[4] and were used as part of the training set of TIGER (Section *TIGER training data*). Furthermore, we added 45 slides from 45 TCGA-BRCA cases (TCGA-NON-TNBC) as part of the sequestered TIGER test set for the computer vision track, 18 for the experimental phase and 27 for the final phase. Regarding molecular subtypes, we included 151 TNBC, 31 Luminal A, 13 HER2+, 1 unknown subtype; regarding morphological subtype, we included 158 infiltrating ductal carcinoma and 20 infiltrating lobular carcinoma; the rest contained infiltrating carcinoma NOS (n=1), medullary carcinoma (n=3), metaplastic carcinoma (n=5), other unspecified tumor types (n=7). Since these cases were solely used for training and benchmarking computer vision performance, we did not consider follow-up data and we included cases beyond TNBC and HER2+. All slides were scanned using an Aperio Scanscope XT scanner, partly at 0.5 um/px and partly at 0.25 um/px.

| | | **Tissue type** | | **Molecular subtype** | | | | | |
|---|---|---|---|---|---|---|---|---|---|
| **Source** | **Slides** | **Biopsies** | **Resections** | **TNBC** | **HER2+** | **Other** | **Purpose** | **Subset** | **Annotators** |
| **Radboudumc** | 26 | 12 | 14 | 11 | 15 | 0 | Training | WSIROI | Group B |
| | 57 | 47 | 10 | 21 | 36 | 0 | Training | WSIBULK | Group C |
| | 45 | 40 | 5 | 12 | 33 | 0 | Training | WSITILS | Group D |
| | 4 | 1 | 3 | 3 | 1 | 0 | Test | CV exp | Group B,C |
| | 7 | 3 | 4 | 4 | 3 | 0 | Test | CV final | Group B,C |
| **Jules Bordet** | 18 | 11 | 7 | 9 | 9 | 0 | Training | WSIROI | Group B |
| | 36 | 13 | 23 | 26 | 10 | 0 | Training | WSIBULK | Group C |
| | 37 | 23 | 14 | 29 | 8 | 0 | Training | WSITILS | Group D |
| | 4 | 2 | 2 | 2 | 2 | 0 | Test | CV exp | Group B,C |
| | 4 | 3 | 1 | 2 | 2 | 0 | Test | CV final | Group B,C |
| **TCGA-BRCA** | 151 | 0 | 151 | 151 | 0 | 0 | Training | WSIROI | Group A |
| | 18 | 0 | 18 | 0 | 6 | 12 | Test | CV exp | Group C |
| | 27 | 0 | 27 | 0 | 7 | 20 | Test | CV final | Group C |
| **TOTAL** | **434** | **155 (35%)** | **279 (65%)** | **270 (62%)** | **132 (30%)** | **32 (8%)** | | | |

**Supplementary Table 1:** Overview of data distribution of manually annotated data within the TIGER challenge. CV: computer vision track.

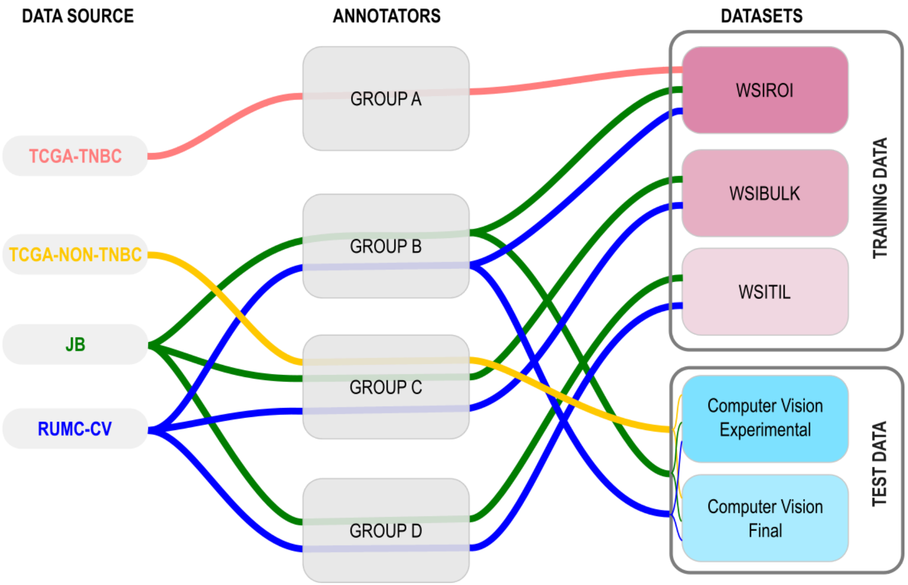


**Supplementary Figure 2:** Overview of data annotation workflow. Each colored line indicates the path from data source to final dataset, passing through the group of annotators.

### Annotations

To build the TIGER training set and the sequestered test sets for the computer vision task (both experimental and final phase), we designed an annotation process using some of the cohorts included in this study: TCGA, JB, RUMC-CV, for a total of n=434 whole-slide images of H&E-stained biopsies or resections. We defined three different types of annotations: 1) manual tissue-cell annotations, i.e., dense annotations (i.e., all pixels were labeled) of invasive tumor, tumor-associated stroma, in-situ tumor, healthy glands, necrosis non in-situ, inflamed stroma, and rest, as well as (x, y) location of lymphocytes and plasma cells within preselected regions of interest; 2) tumor bulk manual annotations, i.e., coarse manual annotations of the regions containing all tumor cells in the slide, guaranteeing that no tumor cells were left outside annotated regions; 3) TILs annotations, i.e., the visual estimation of the percentage of stromal TILs in the slide, scored based on the recommendations of the TILs WG.

#### Groups of annotators

Manual annotation tasks were performed by four different groups of experts (see Figure data-annotations).

*Group A* consisted of a team of annotators involved in the BCSS project[3] (focusing on tissue annotations) and the NuCLS project[4] (focusing on cell annotations), where a crowdsourcing platform was created to manually annotate tissue compartments and cell locations. This group made tissue-cell annotations on n=151 slides from the TCGA-BRCA public archive. Original annotations contained a larger set of classes for both tissue compartments and cell types, which were here relabeled to match the set of classes defined in TIGER (see Supplementary Tables 2 and 3 for a list of corresponding labels).

*Group B* consisted of five pathologists (LCB, DP, EISS, AL, HH) (years of practice: 2.5-20) with experience in breast pathology (1-15 years of experience), members of the International

Immuno-Oncology Working Group (since 5.5-7 years). They made tissue-cell annotations on n=63 slides from RUMC-CV and JB using the CIRRUS Pathology web viewer and the functionalities of reader studies available on Grand Challenge. In case they were not certain about their annotations, they were given the possibility to label them as “uncertain”, and a consensus meeting was later organized to come to an agreement and decide on the final labels.

*Group C* consisted of three resident pathologists (LT, MB, JB) and research assistants from Radboudumc. This group performed two tasks. First, it performed tissue-cell annotations on a set of n=45 slides from TCGA. Second, it made coarse annotations of the tumor bulk, i.e., regions containing all tumor cells, in a set of n=93 slides from Radboudumc and Jules Bordet.

*Group D* consisted of one board-certified breast pathologist (RS) with >10 years of experience on TILs assessment. This group visually estimated the percentage of stromal TILs in a set of n=82 slides with values between 0 and 100, following the recommendations of the TIL WG. This pathologist also provided comments, when appropriate, on the main characteristics of the slide and highlighted potential pitfalls for TILs scoring, in line with what reported in[5].

#### Manual annotations

Manual annotations of tissue compartments were made in pre-selected regions of interest (ROIs). For tissue annotations, we defined the following classes: *invasive tumor*, containing regions of the invasive tumor, including several morphological subtypes, such as invasive ductal carcinoma and invasive lobular carcinoma; *tumor-associated stroma*, containing regions of stroma (i.e., connective tissue) that are associated with the tumor; *in-situ tumor*, containing regions of in-situ malignant lesions, such as ductal carcinoma in situ (DCIS) or lobular carcinoma in situ (LCIS); *healthy glands*, containing regions of glands with healthy epithelial cells; *necrosis not in-situ*, containing regions of necrotic tissue that are not part of in-situ tumor; *inflamed stroma*, containing tumor-associated stroma that has a high density of lymphocytes (i.e., it is "inflamed"); *rest*, containing regions of several tissue compartments that are not specifically annotated in the other categories; examples are healthy stroma, erythrocytes, adipose tissue, skin, nipple, etc.

For the TCGA-TNBC cohort, ROIs and manual annotations were predefined in the BCSS project. In TIGER, we re-mapped labels according to the TIGER classes. For RUMC-CV, JB and TCGA-NON-TNBC, three ROIs per slide (on average) of size 500x500 micron were manually pre-defined by a resident pathologist (LT) and then manually annotated by either Group B or Group C.

In most ROIs, lymphocytes and plasma cells were annotated using point annotations and then squared bounding boxes of 8x8 micron were constructed centered on the point annotation. This fixed-size bounding box size is inspired by previous work on lymphocyte detection[6].

Annotations from groups A,B,C were finally checked for consistency and harmonized, when necessary, by two resident pathologists (MB, LT).

Data was split into a training set for training computer algorithms to segment tissue and detect cells, and a test set for the computer vision track, on detection and segmentation.

| **BCSS label** | **TIGER label** |
|---|---|
| normal_acinus_or_duct | healthy glands |
| mostly_lymphocytic_infiltrate | inflamed stroma |
| exclude | exclude (label = 0) |
| mostly_stroma | tumor-associated stroma |

| | |
|---|---|
| necrosis_or_debris | necrosis not in-situ |
| mostly_plasma_cells | inflamed stroma |
| roi | tumor-associated stroma |
| glandular_secretions | rest |
| nerve | rest |
| other_immune_infiltrate | inflamed stroma |
| angioinvasion | invasive tumor |
| lymphatics | rest |
| blood_vessel | rest |
| mostly_blood | rest |
| skin_adnexia | rest |
| mostly_tumor | invasive tumor |
| mostly_dcis | in-situ tumor |
| idc | invasive tumor |
| metaplasia_NOS | rest |
| undetermined | rest |
| mostly_fat | rest |
| mostly_mucoid_material | rest |
| background | exclude (label = 0) |

**Supplementary Table 2.** Label correspondence between BCSS and TIGER

| **NuCLS cell label** | **TIGER cell label** |
|---|---|
| Tumor | excluded |
| Mitotic figures | excluded |
| Fibroblast | excluded |
| Vasc. end. | excluded |
| Macrophage | excluded |
| Lymphocyte | Lymphocytes and plasma cells |

| | |
|---|---|
| Plasma cells | Lymphocytes and plasma cells |
| Neutrophils | excluded |
| Eosinophils | excluded |
| Myoepith. | excluded |
| Normal ep. | excluded |
| Apoptotic | excluded |
| Unlabeled | excluded |

**Supplementary Table 3.** Label correspondences between NuCLS and TIGER.

### TIGER training data

The training data consisted of n=370 slides, which we divided into three sub-datasets: WSIROIS (n=195) containing tissue-cell annotations, WSIBULK (n=93) containing tumor-bulk annotations, and WSITILS (n=82), containing TILs annotations.

#### WSIROIS

The WSIROIs dataset was specifically designed for training computer models to perform the task of segmentation and detection. It consists of n=195 whole-slide images of breast cancer, both (core-needle) biopsies, and surgical resections, with regions of interest (ROI) selected and manually annotated. This dataset contains images and annotations from multiple sources: n=151 slides from TCGA-TNBC, n=26 slides from RUMC-CV, n=18 slides from JB. Annotations were made by Group A (for TCGA-TNBC) and Group B (for RUMC-CV and JB). Supplementary Figure 3 depicts examples of manual annotations of tissue compartments and cells in the WSIROIS training set.

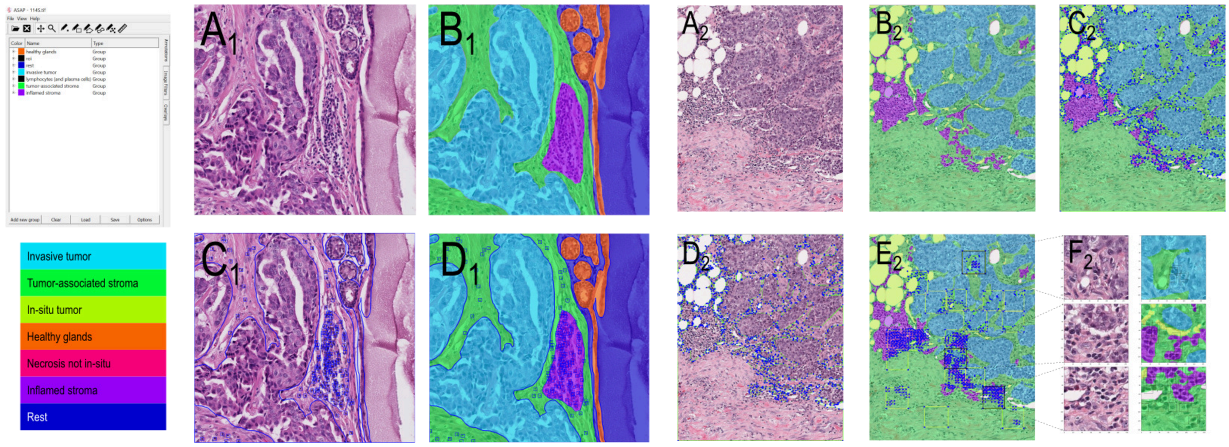


**Supplementary Figure 3:** Visual example of annotations in the WSIROIs dataset. **Left**: visual example of the first ROI in case 114S in the WSIROIS training set (A1), the corresponding mask of manually annotated tissue compartments (B1, from the TIF file), the tissue borders and bounding boxes from the XML file (C1) and the combination of B1 and C1 in D1. We show an overview of labels when XML files are loaded on the ASAP viewer, and a legend with class names and corresponding colors when the "label" LUT is used in ASAP with 255/127 window/level setting. **Right**: A2 depicts an ROI from the WSIROIS dataset of TIGER derived from the BCSS dataset. In TIGER, we also release corresponding annotations from BCSS where we have merged the original classes and relabeled them (see relabeling map in the text below) to match TIGER classes (B2), both as TIF files and as XML files (see C2 and D2). Within this ROI, the NuCLS dataset provides cell annotations in smaller ROIs (see E2). In each smaller ROI, bounding box annotations of lymphocytes + plasma cells were released (see F2 for details of 3 ROIs).

### WSIBULK

The WSIBULK dataset consisted of 93 WSIs, including biopsies and surgical resections of TNBC and HER2+ breast cancer tissues from RUMC-CV and JB. In this set, we provided coarse annotations of the "tumor bulk," referring to manual annotations of one or more regions in the slide containing invasive tumor cells. These annotations offer a rough estimate of the tumor extent in the slide (see Supplementary Figure 4 for examples), ensuring that all tissue outside of the annotations does not contain invasive tumor cells, although they may contain tumor cells related to in-situ lesions. The aim of this dataset was to give researchers the opportunity to implement strategies for identifying a tumor bulk, as well as considering regions that are guaranteed to be free of invasive tumor in the image. During the annotation process, the exact distance between the annotated border and the invasive front of the tumor was not considered and, therefore, this could vary among cases.

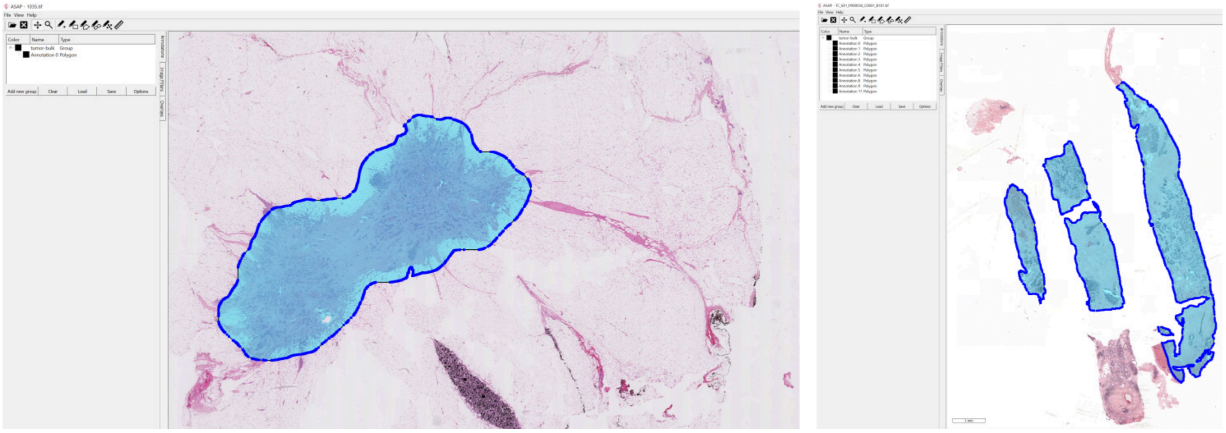

**Supplementary Figure 4**. Visual example of annotations in the WSIBULK dataset. In cyan the manual annotations of the tumor bulk in a surgical resection (left) and core-needle biopsy (right) of breast cancer tissue.

### WSITILS

The WSITILS dataset comprised 82 whole-slide images (WSIs) of biopsies and surgical tissue resections from TNBC and HER2-positive breast cancer patients. For each slide, a stromal TIL score was assessed via visual inspection, without the support of any manual annotation, by a board-certified pathologist (RS) with >10 years of experience in scoring sTILs. The assessment of sTILs was done adhering to the TILs working group's recommendations[7]. The aim of this dataset was to allow researchers to compare and scale their automated scores against the visual assessment by pathologists.

#### Release of training data

The n=370 training whole-slide images and their annotations were publicly released under a non-commercial Creative Commons License (CC BY-NC 4.0). Image data was shared at 20X magnification (0.5 micron/px spacing), tissue annotations were shared both as XML files with polygon coordinates and as multi-class masks in TIFF format, whereas cell annotations were shared both as part of the XML files and in the COCO format[8] for object detection, commonly used in the computer vision community. Data was released via two channels. First, as part of the AWS Open Data Registry[9], where the full set of whole-slide images was made available, for a total storage of 169.2 GB. Second, as part of the Zenodo platform[10], where only a lightweight version of the data was shared, namely only images and annotations limited to the content of the ROIs in the WSIROI set, without sharing full whole-slide images, for a total storage of 2.6 GB of compressed data. With this, we aimed at making a dataset to allow participants without particular expertise on WSI reading and processing a quick approach to TIGER.

### TIGER challenge evaluation

#### Computer Vision Track

##### Data

We selected 64 whole-slide images from RUMC-CV, JB and TCGA-NON-TNBC, and used them to build the sequestered test sets for the computer vision track of TIGER: n=26 slides (4 from RUMC-CV, 4 from JB, 18 from TCGA-NON-TNBC) for the test dataset of the experimental phase, and n=38 (7 from RUMC-CV, 4 from JB, 27 from TCGA-NON-TNBC) for the test set of the final phase.

The *experimental* test set contained the following data distribution: molecular subtypes (9 HER2+, 5 TNBC, 11 Luminal A, 1: Not Available); tissue type (23 resections, 3 biopsies); morphological subtype (11 IDC, 7 ILC, 1 apocrine carcinoma, 5 invasive NST, 1 malignant adenomyoepithelioma, 1 lobular).

The *final* test set contained the following data distribution: molecular subtypes (12 HER2+, 6 TNBC, 20 Luminal A); tissue type (32 resections, 6 biopsies); morphological subtype (26 IDC, 9 ILC, 1 neuroendocrine carcinoma, 1 apocrine carcinoma, 1 micropapillary).

In all slides we performed manual annotations in pre-selected regions of interest (see section *Manual annotations* for details). Since the purpose of this dataset was to measure computer vision performance and not their clinical implications, we decided to also include Luminal A cases in both sets, arguing that algorithms should be capable of interpreting morphological characteristics regardless of the molecular subtype. Furthermore, because we solely focused on regions of interest, the skewed distribution of tissue types (88% resections and 12% biopsies in experimental, 84% resections and 16% biopsies in the final set) does not affect the evaluation. In the rest of this manuscript, we also refer to the test set of the final phase as the TIGER-CV dataset, and we refer to the set of manually annotated regions and lymphocytes in TIGER-CV as $SEG_{TIGER}$ and $DET_{TIGER}$ respectively.

##### Evaluation

In the leaderboards of the computer vision track, we ranked algorithms based on their computer vision performance at tissue segmentation and cell detection. For tissue segmentation, we computed an overall Dice score as the average of two Dice scores, one for invasive tumor and one for tumor-associated stroma. The motivation for focusing on these two classes is that regions of invasive tumor and of tumor-associated stroma play a central role in the definition of the TILs score. The Dice-scores were computed over all slides by taking the target class (invasive tumor or tumor-associated stroma, respectively) as foreground and rest (including tumor-associated stroma or invasive tumor, respectively) as background.

For cell detection we performed a Free Response Operating Characteristic (FROC) analysis, computing sensitivity (true positive rate, TPR) versus average false positives (FP) per mm² over all slides. Given the i-th predicted cell locations $c_{p,i} = (x_{p,i}, y_{p,i})$ of lymphocytes and plasma cells, each with likelihood $l_i$, and the j-th manual annotation $c_{m,j} = \left(x_{m,j}, y_{m,j}\right)$, we considered a hit of manual annotation $c_{m,j}$ if $l_i > \tau$ and if $d\left(c_{p,i}, c_{m,j}\right) \leq D$, where $\tau$ is a threshold and $d(\bullet)$ is the Euclidean distance. In our case, τ assumed all likelihood values predicted by each algorithm on the entire test set, and D=4μm, based on previous work[6]. For each threshold τ, we obtained values for the true positives (TPs), false negatives (FN)s, and false positives (FPs) and used those to build an FROC curve. From the curve, we derived a "FROC score" by averaging sensitivity computed at five pre-selected values of FP/mm²: [10, 20, 50, 100, 200, 300]. Finally, methods were ranked based on the average (named “mean position” in the leaderboard) of the segmentation and detection rankings. The same evaluation procedure was used to build leaderboards in both the experimental and the final phase of TIGER. Supplementary Table 4 reports the leaderboard of the final phase of the computer vision track of TIGER.

| Team | Ts_dice (ranking) | Lymph_froc (ranking) | Mean position |
|---|---|---|---|
| Biototem | 0.8115 (1) | 0.5504 (1) | 1 |
| TIAger | 0.7872 (3) | 0.5441 (2) | 2.5 |
| AIVIS | 0.8048 (2) | 0.4820 (4) | 3 |
| GDPH_MIA | 0.7680 (4) | 0.4572 (6) | 5 |
| Spotlight Pathology | 0.7457 (5) | 0.4739 (5) | 5 |
| FengHu | 0.6616 (10) | 0.5437 (3) | 6.5 |
| FDA-CDRH-OSEL-DIDSR | 0.7387 (6) | 0.3205 (7) | 6.5 |
| PAIR | 0.7139 (7) | 0.2896 (8) | 7.5 |
| Mart.varijthoven (Radboud) | 0.6942 (8) | 0.2734 (9) | 8.5 |
| DeepDrivePL | 0.6668 (9) | 0.2232 (10) | 9.5 |
| 大胖胖墩 (CellsVision) | 0.5442 (11) | 0.0329 (11) | 11 |

**Supplementary Table 4:** Overview of the results of the final leaderboard of the computer vision track at the end of the TIGER challenge in June 2022.

### Survival Track

#### Data

We pooled together the RUMC-SURV$_{TNBC}$ and the FinHER cohorts and randomly split them into two sets for the analysis of the prognostic value of TILs scores in TIGER: the survival experimental test set (n=200) and the survival final test set (n=707). Differently from other challenges, where a fairly balanced split is made between the experimental and the final set, in TIGER the size of the experimental set was bounded by the computing budget available to the challenge organizers, which defined n=200 as the optimal sample size to allow participants to run multiple evaluations on a dataset with a sufficient number of events during the challenge within the computational limit set by the Grand Challenge platform.

#### Evaluation

To build a leaderboard in the survival track of TIGER, for each submission we built a multivariable Cox regression model (CRM) trained using as covariates a set of clinical variables (age group, morphology subtype, grade, molecular subtype, stage, surgery, adjuvant therapy) and the cTILs scores produced by the submitted algorithm. Successively, we used this model as a predictor of DFI and computed the Uno's concordance index (C-index)[11] of this model and rank algorithms based on its value. Given the N samples in the test set (N=200 for the experimental and N=707 for the final set), we split the set into K folds (with K=5), built K CRM using data from 4 folds and applied the CRM to the held-out set to compute the C-index. We repeated this process B=500 times shuffling the N samples at each iteration. The final C-index was computed as the average of all C-indexes and used to rank algorithms in the leaderboard (see Supplementary Table 5).

Additional information was also provided to participants for each submission via the Grand Challenge platform, although not used to rank methods in the survival leaderboard. First, we computed the 95% confidence interval (CI) over the distribution of the B*K values of C-indexes. Second, we computed Kaplan Meier curves by using the median TIL score as cut-off value from each submission (see Supplementary Figure 5).

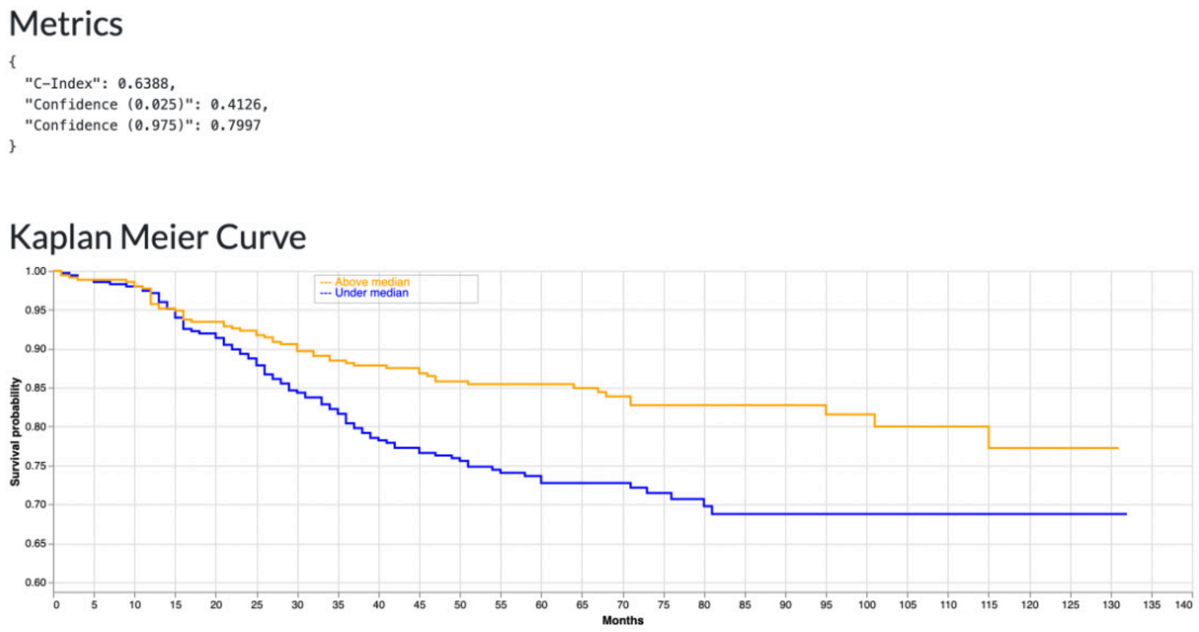


**Supplementary Figure 5:** Example of visualization of output during the experimental phase of the survival track of TIGER. Both the Kaplan Meier curve plotted based on the median value of computed cTILs and the Uno C-index with its confidence interval were visualized.

| Team | C-index (Uno) |
|---|---|
| VUNO | 0.6388 |
| mart.vanrijthoven (Radboud) | 0.6338 |
| Spotlight Pathology | 0.6224 |
| 大胖胖墩 (CellsVision) | 0.6120 |
| FDA-CDRH-OSEL-DIDSR | 0.6034 |
| SRI | 0.5996 |
| Clinical variables only | 0.5903 |
| TIAger | 0.5879 |
| Biototem | 0.5793 |

**Supplementary Table 5: tiger_final_survival.** Overview of the results of the final leaderboard L2 at the end of the TIGER challenge in June 2022.

### Re-stained data

We selected n=4 breast cancer cases (2 TNBC, 2 HER2+) that were part of the computer vision task test set used in the experimental phase of TIGER, and performed a re-staining using a combination of immunohistochemical markers. For this purpose, we collected tissue blocks from the archive of Radboudumc, cut 4um slides, stained with H&E, scanned with a 3DHISTECH Pannoramic 1000 scanner, restrained with IHC (mix of CD3 and CD79a), and finally scanned again, resulting in four pairs of whole-slide images. As part of the re-staining process, the HE-stained sections were subjected to acetone treatment for 5-10 minutes until the tape came off, followed by 5 minutes of fresh acetone, 5 minutes of 100% ethanol, and 5 minutes of tap water. These steps were performed to prepare the sections for restaining and de-staining, with most of the destaining occurring during antigen retrieval. Any remaining staining was removed during the peroxidase step, which involved antigen retrieval with TRIS-EDTA pH9 (resulting in section discoloration) and two 10-minute peroxidase block steps. Antibodies in a mix of CD3 (rabbit monoclonal antibody, clone SP7, Thermo Fisher Scientific, Fremont, CA) with a 1:100 concentration and CD79a (clone JCB117, Agilent, Santa Clara, CA) with a 1:800 concentration were applied, followed by Envision Flex HRP DAKO DM 842 (anti-mouse/rabbit) and color development with DAB, and hematoxylin staining.

After scanning, both the H&E and IHC stained slides were registered using the HistokatFusion WSI registration software developed by Fraunhofer MEVIS[12] (Supplementary Figure 6).

We manually selected n=26 regions of interest of average size 798 $mm^2$ (min: 178, max: 1530) to contain a variety of positivity to IHC markers, as well as regions with negative staining (i.e., lack of lymphocytes). Using the open-source ASAP software[13], a research assistant annotated lymphocytes in the IHC slide, transferred them to H&E via registration and adjusted them when necessary (e.g., lymphocyte not clearly visible in H&E; location shifted do to locally suboptimal registration), resulting in a set of 2402 IHC-confirmed lymphocytes. Annotations were finally checked by a resident pathologist. We refer to this dataset as $DET_{RESTAIN}$.

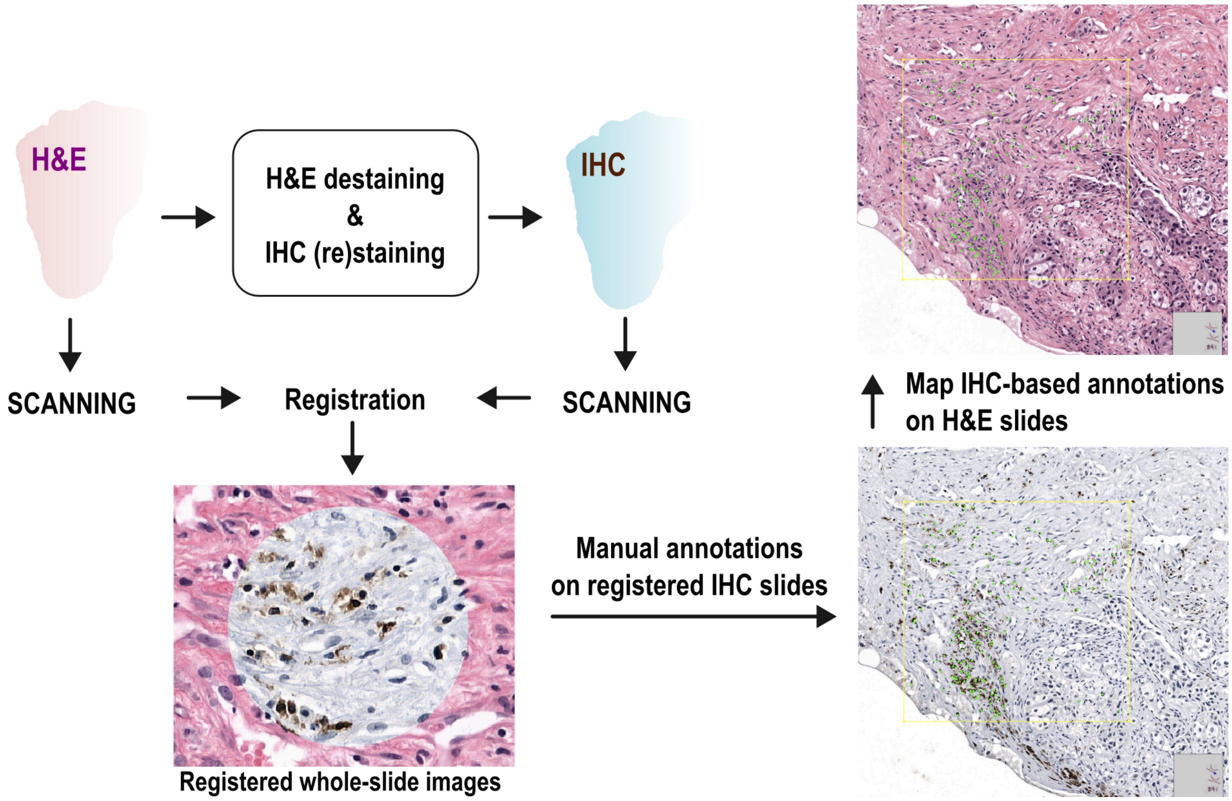


**Supplementary Figure 6:** Schematic representation of the pipeline to generate IHC-based manual lymphocyte annotation in H&E slides. An H&E slide is first scanned, de-stained and re-stained using a cocktail of IHC markers, and finally scanned. Digital slides are registered using the HistokatFusion algorithm[12] (the circle in the middle implements the "spy view" mode of HistokatFusion, indicating the IHC area of the image corresponding to HE after registration). The manual annotations are made in pre-selected regions of interest in IHC slides, and then mapped onto the corresponding registered H&E digital slides.

#### Scatter Correlations

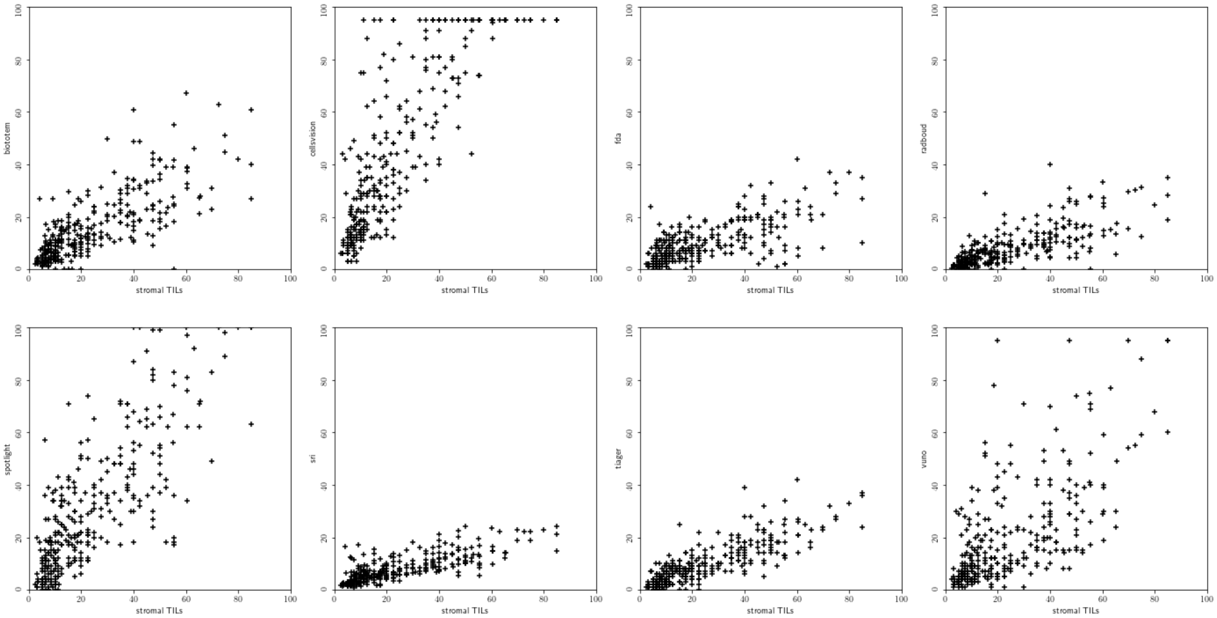


**Supplementary Figure 7:** Scatter Plots showing the correlation between vTILs (here referred to as stromal_TILs) and cTILs in the FinHER.

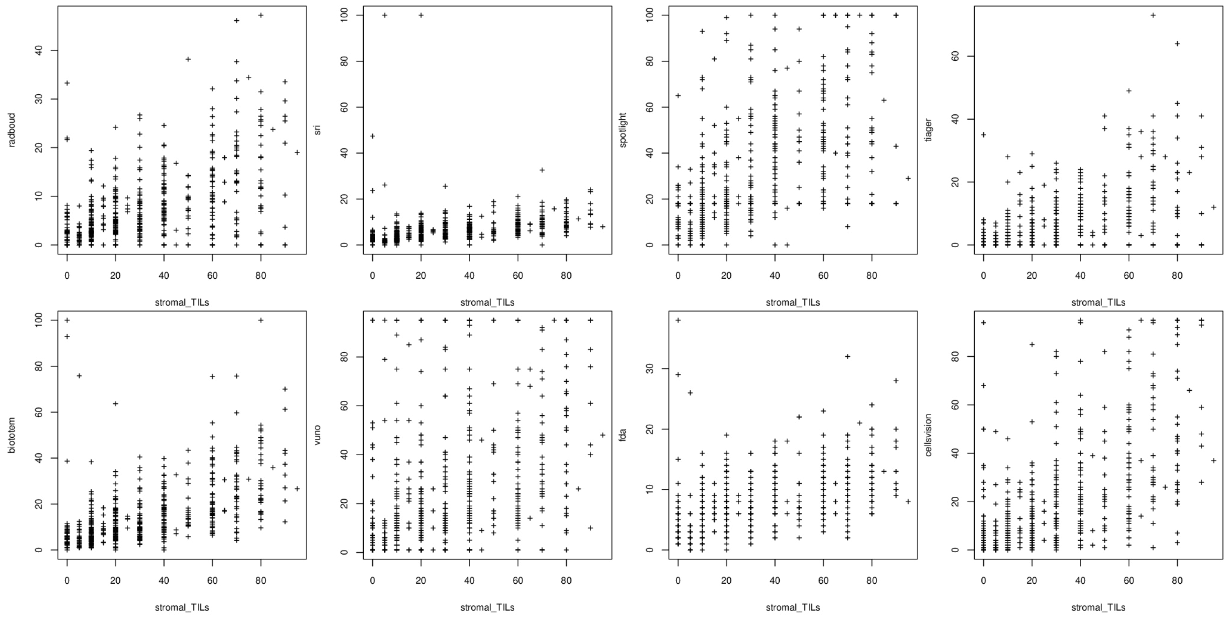


**Supplementary Figure 8:** Scatter Plots showing the correlation between vTILs (here referred to as stromal_TILs) and cTILs in the G6 cohort.

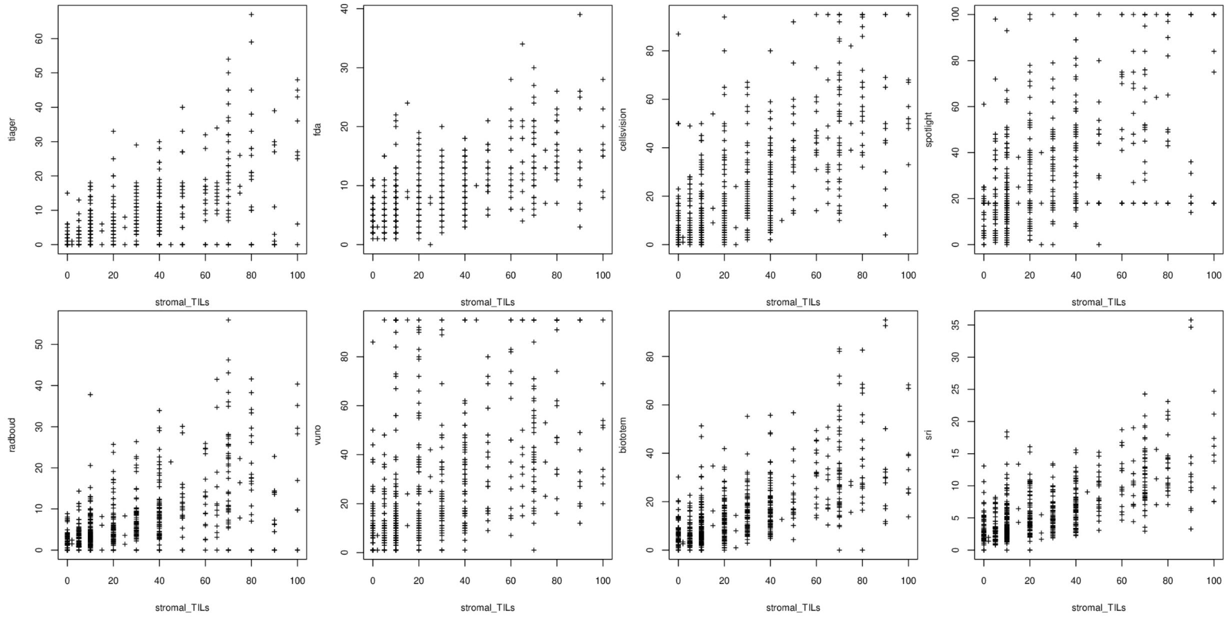


**Supplementary Figure 9:** Scatter Plots showing the correlation between vTILs (here referred to as stromal_TILs) and cTILs in the G7 cohort.

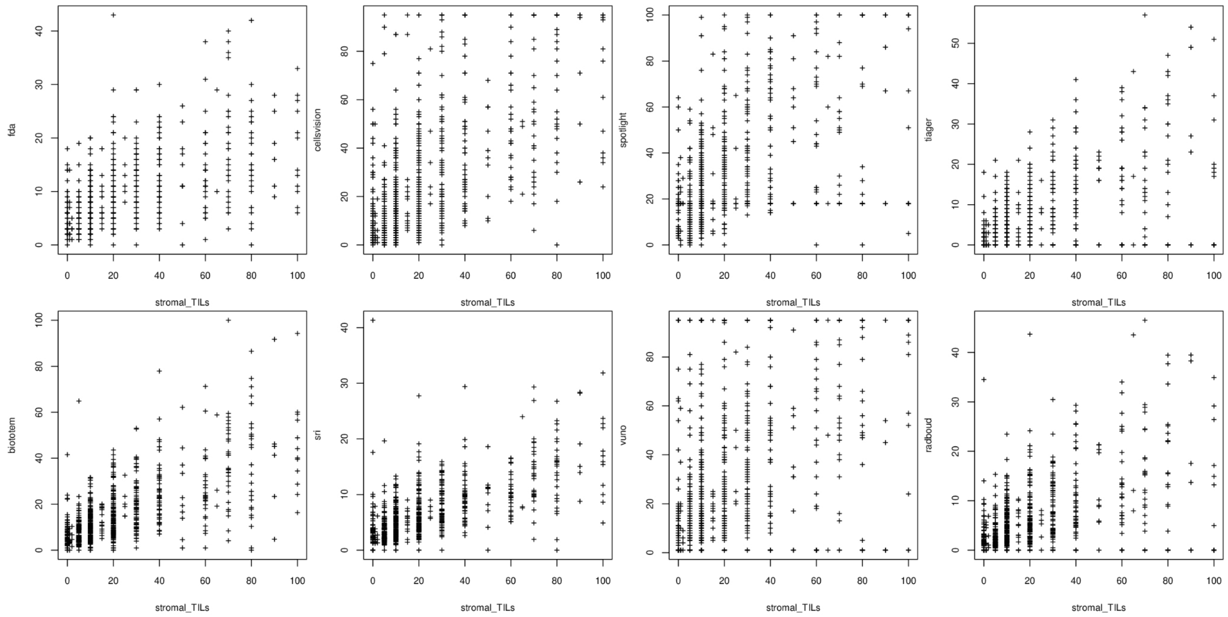


**Supplementary Figure 10:** Scatter Plots showing the correlation between vTILs (here referred to as stromal_TILs) and cTILs in the G8 cohort.

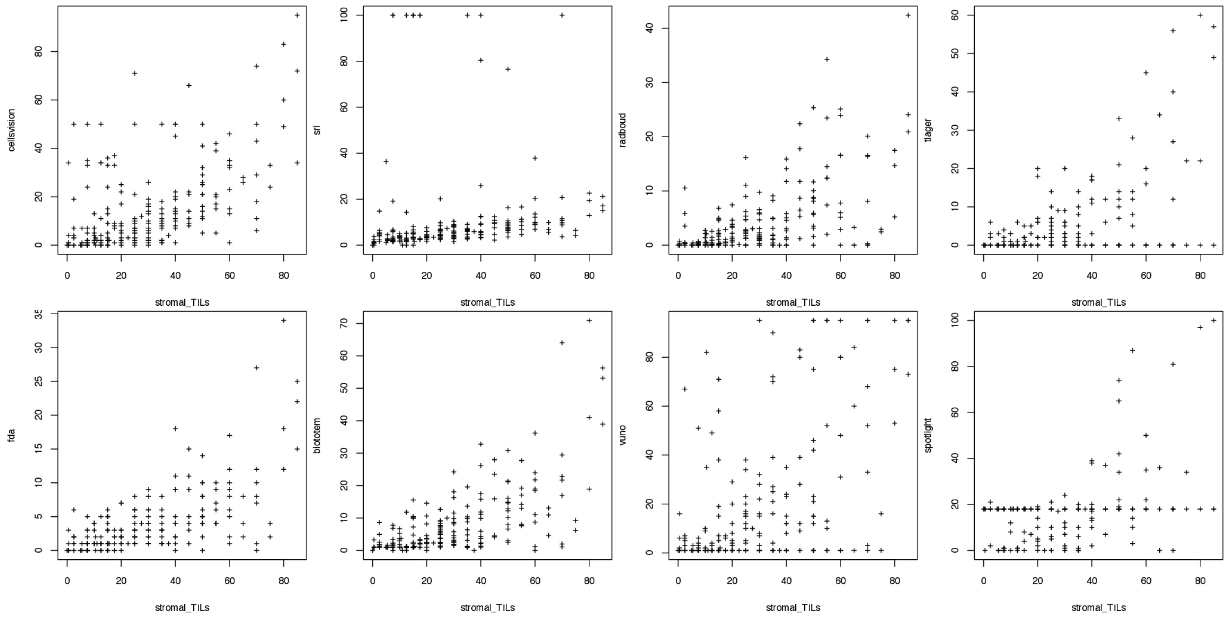


**Supplementary Figure 11:** Scatter Plots showing the correlation between vTILs (here referred to as stromal_TILs) and cTILs in the NKI cohort.

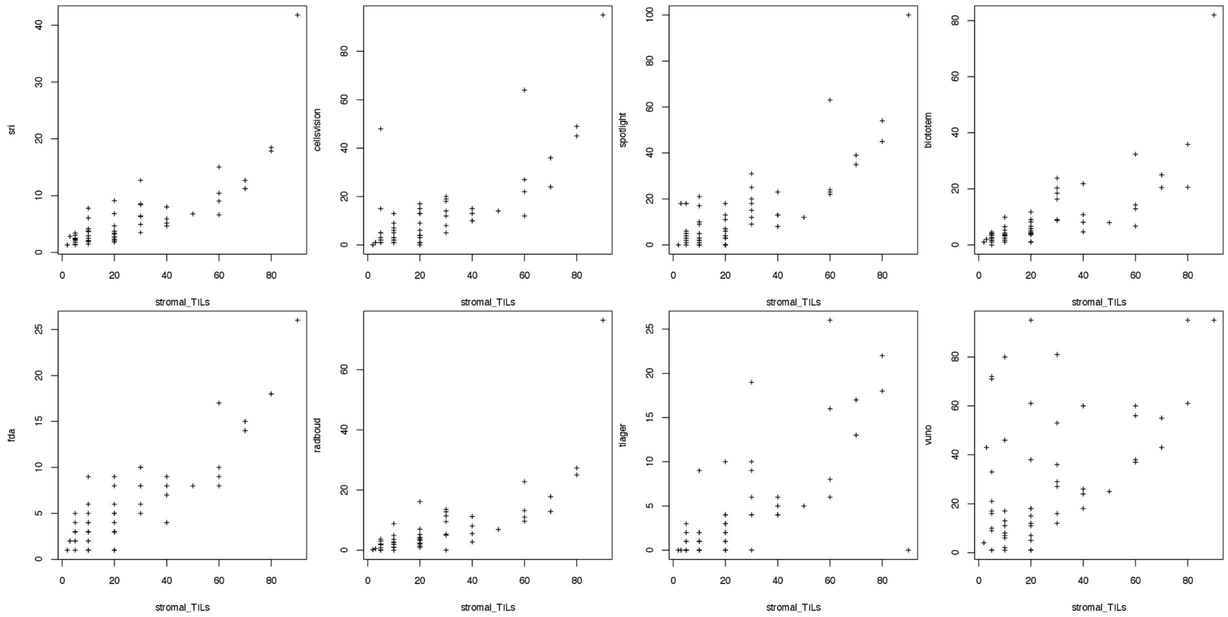


**Supplementary Figure 12:** Scatter Plots showing the correlation between vTILs (here referred to as stromal_TILs) and cTILs in the SCDC cohort.

### Resections - Kaplan-Meier curves

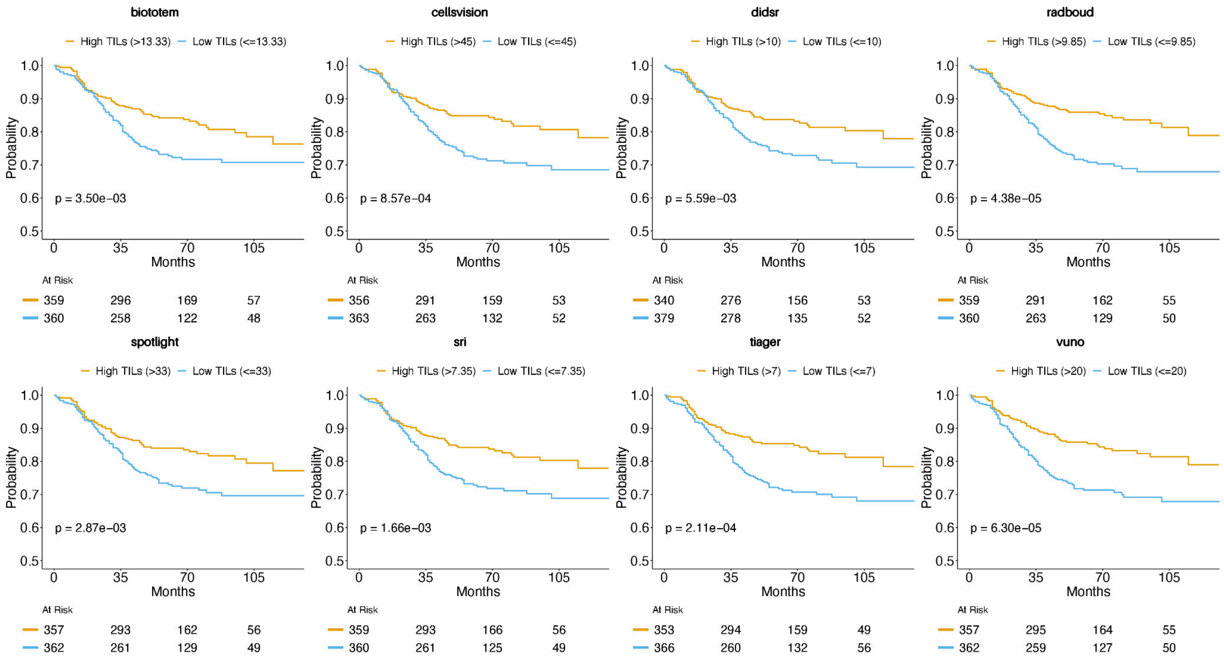


**Supplementary Figure 13:** Kaplan-Meier Curves for Disease-Free Interval (DFI) in Triple-Negative Breast Cancer (TNBC)

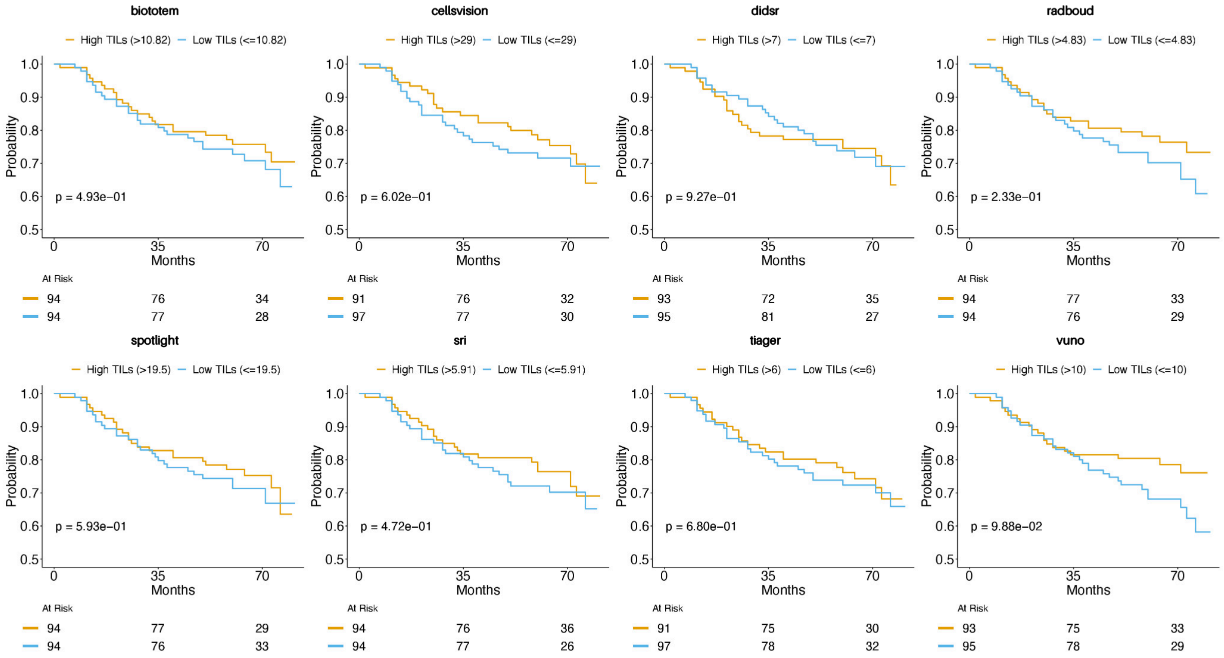

**Supplementary Figure 14:** Kaplan-Meier Curves for Disease-Free Interval (DFI) in HER2-Positive Breast Cancer Patients Receiving Resections.

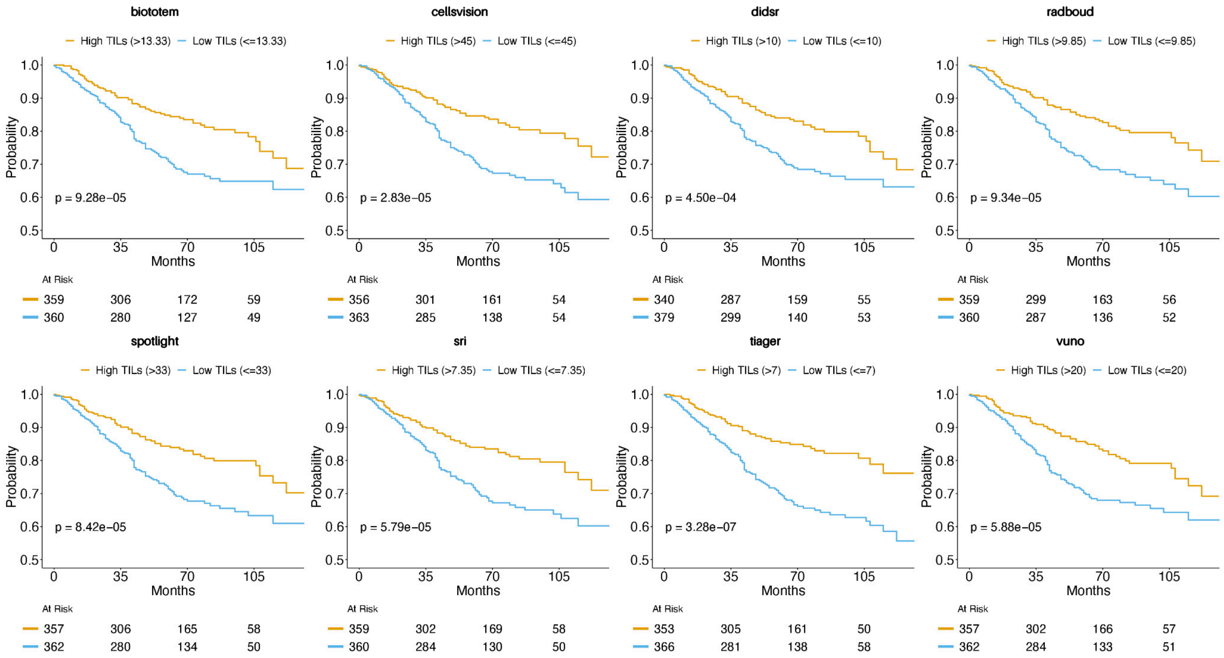


**Supplementary Figure 15:** Kaplan-Meier Curves for Overall Survival (OS) in TNBC

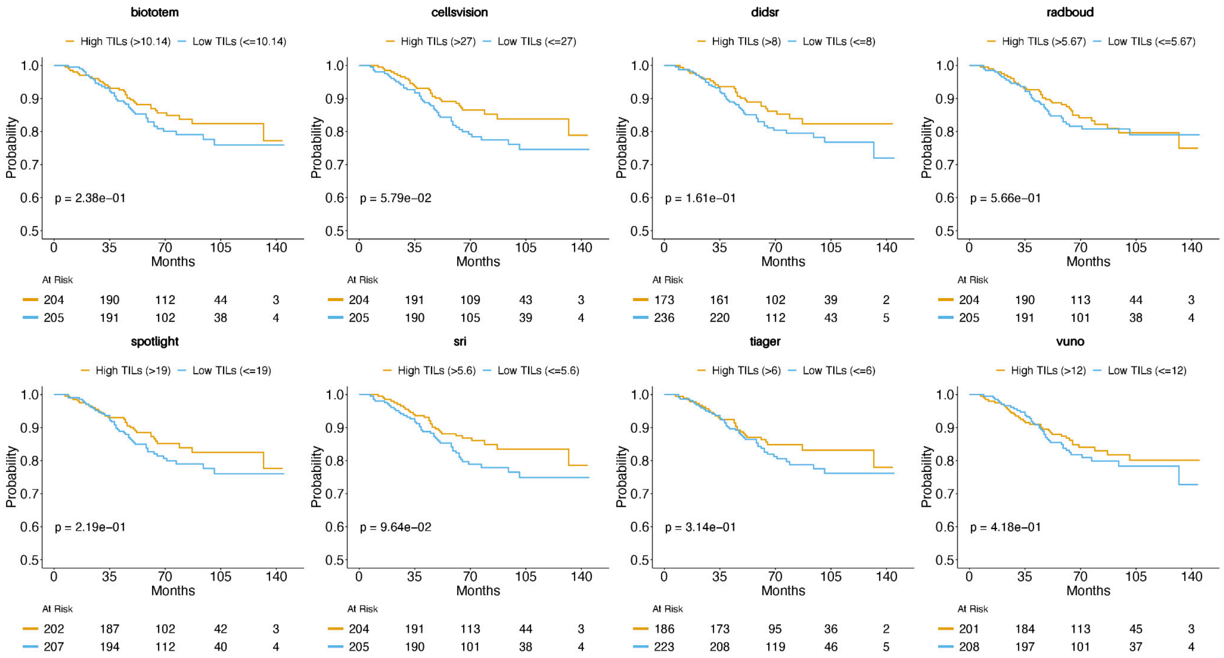


**Supplementary Figure 16:** Kaplan-Meier Curves for Overall Survival (OS) in HER2-Positive Breast Cancer

#### Biopsies - Kaplan Meier Curves

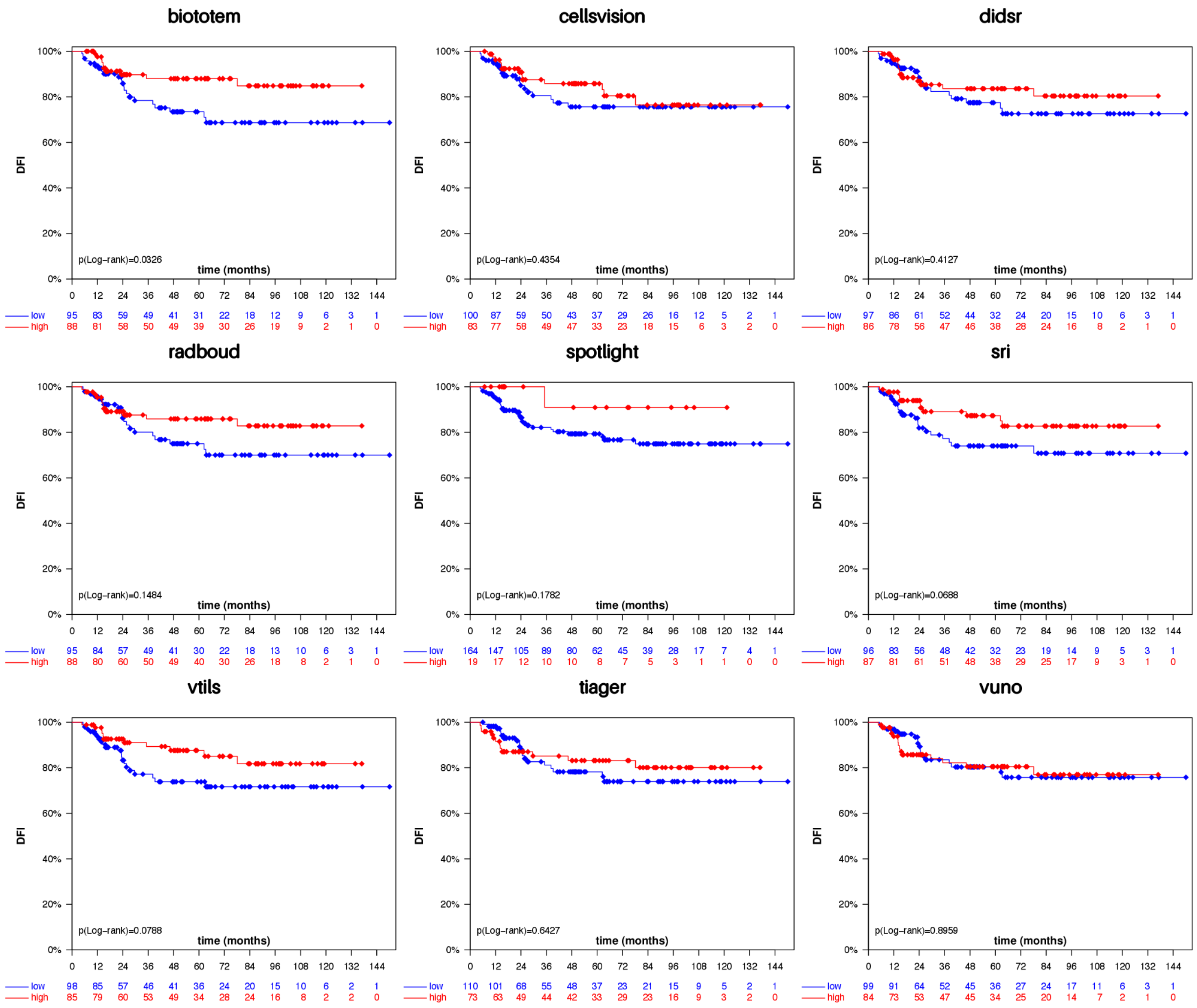


**Supplementary Figure 17:** Kaplan-Meier Curves for Disease-Free Interval (DFI) in NKI cohort.

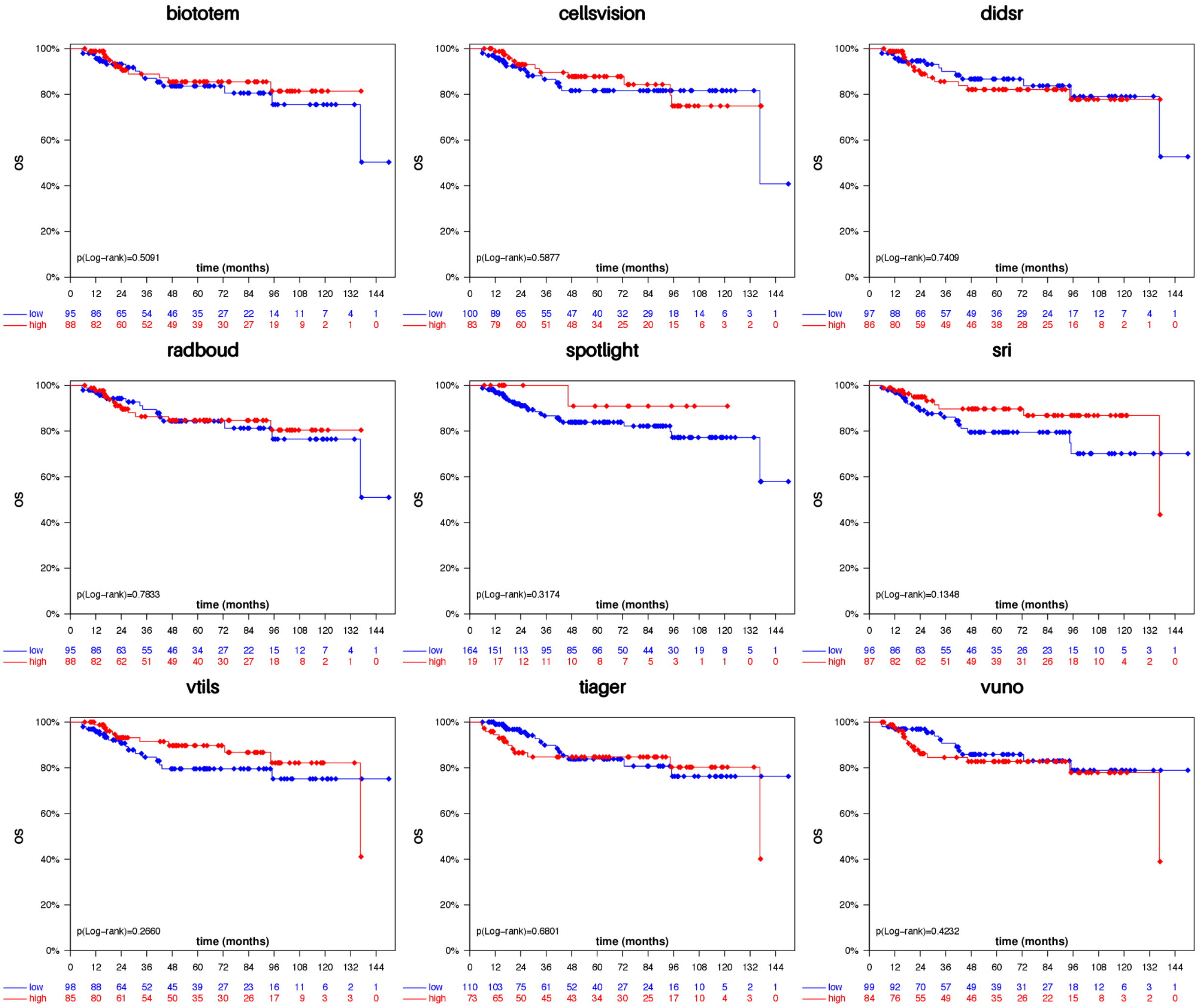


**Supplementary Figure 18:** Kaplan-Meier Curves for Overal Survival (OS) in NKI cohort.

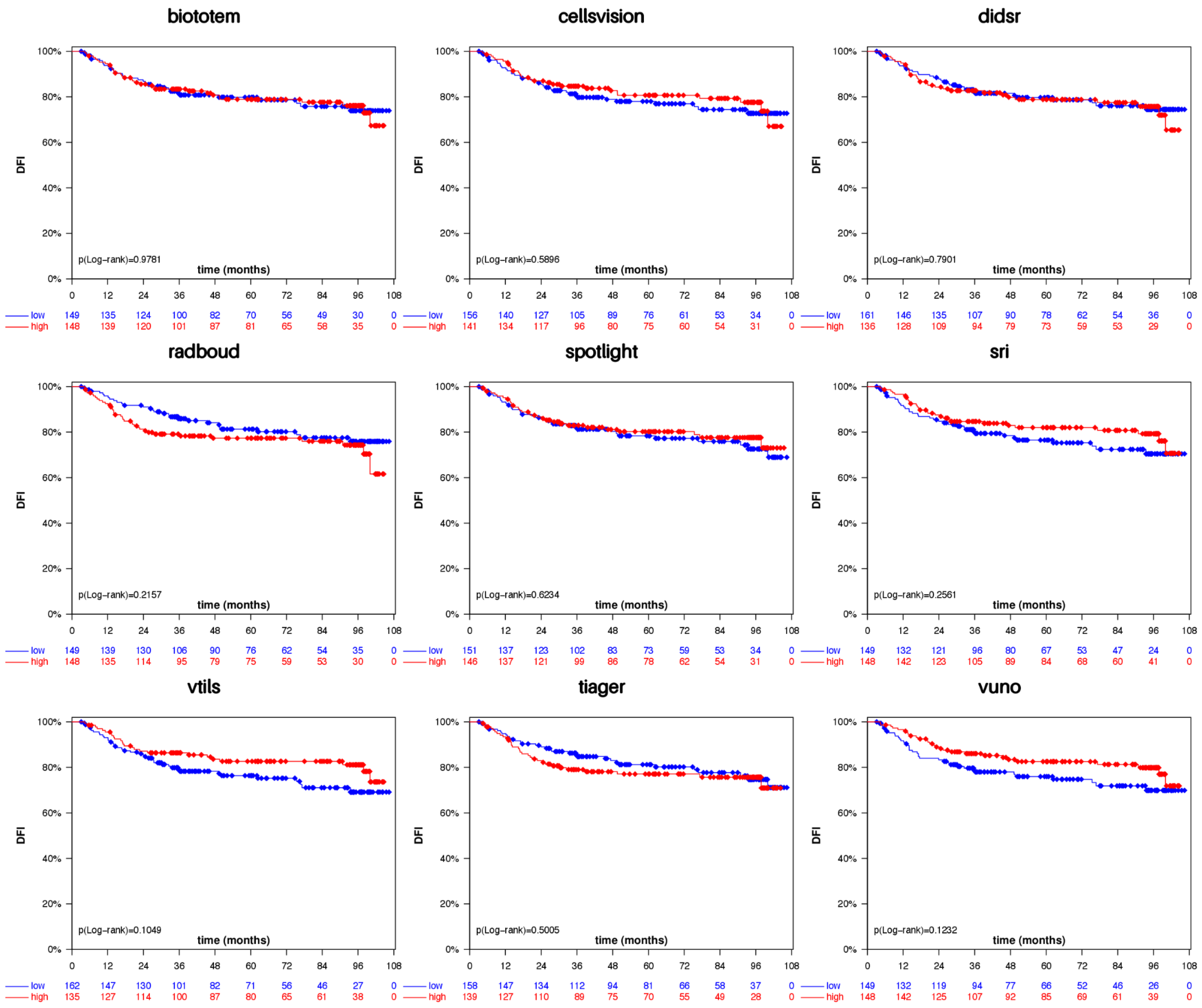


**Supplementary Figure 19:** Kaplan-Meier Curves for Disease-Free Interval (DFI) in G6 (TNBC) cohort.

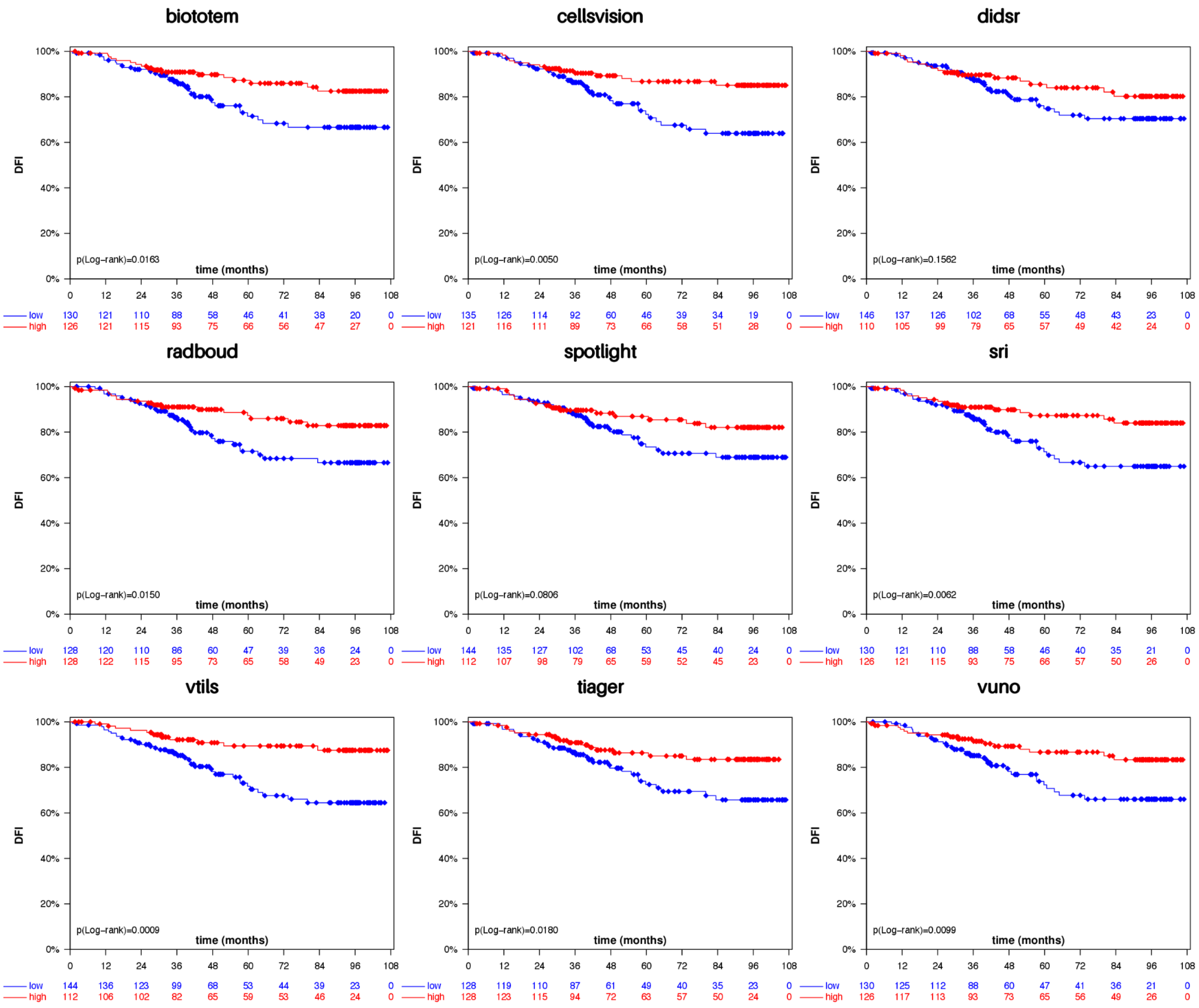


**Supplementary Figure 20:** Kaplan-Meier Curves for Disease-Free Interval (DFI) in G6 (Her2+) cohort.

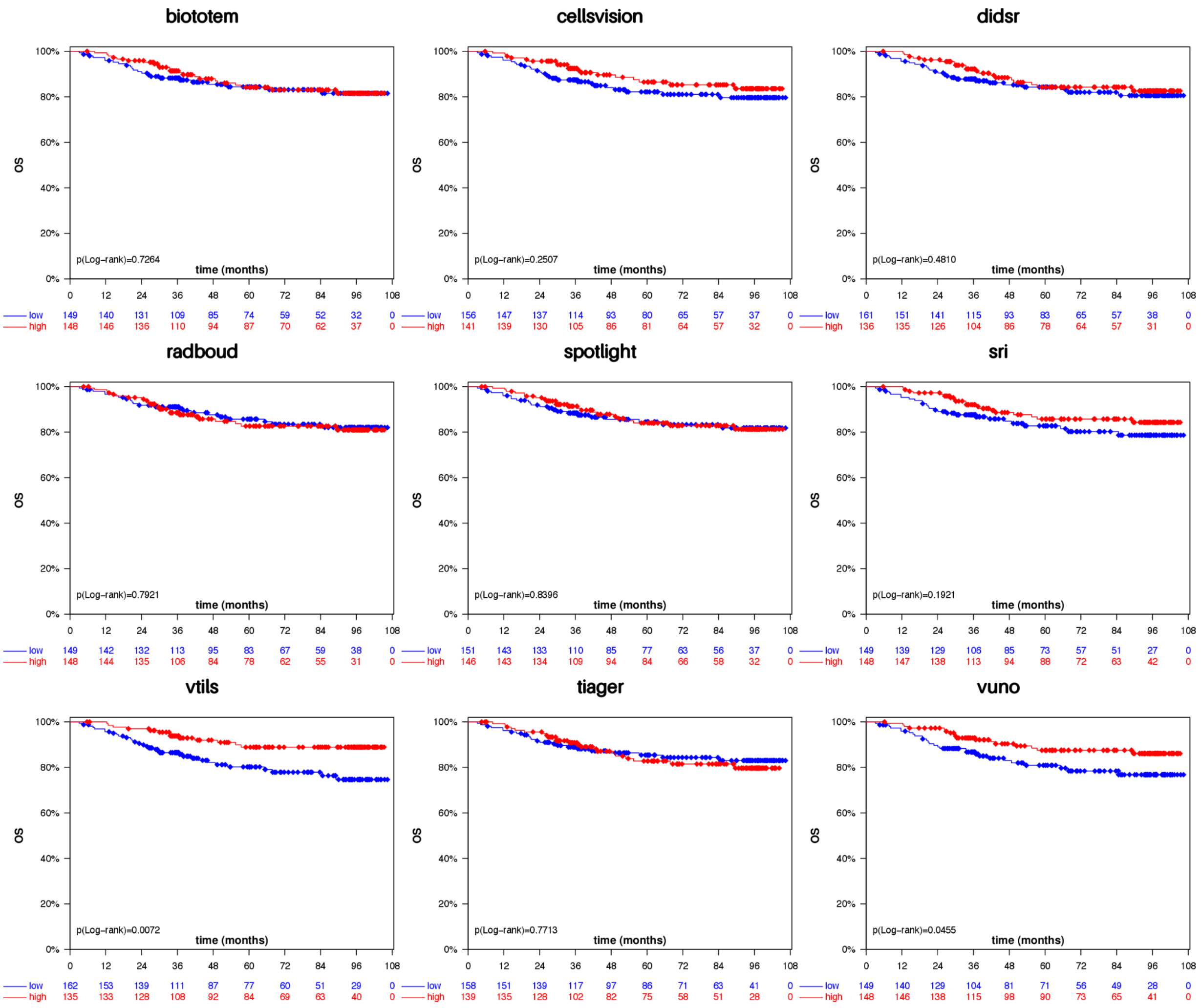


**Supplementary Figure 21:** Kaplan-Meier Curves for Overall Survival (OS) in G6 (TNBC) cohort.

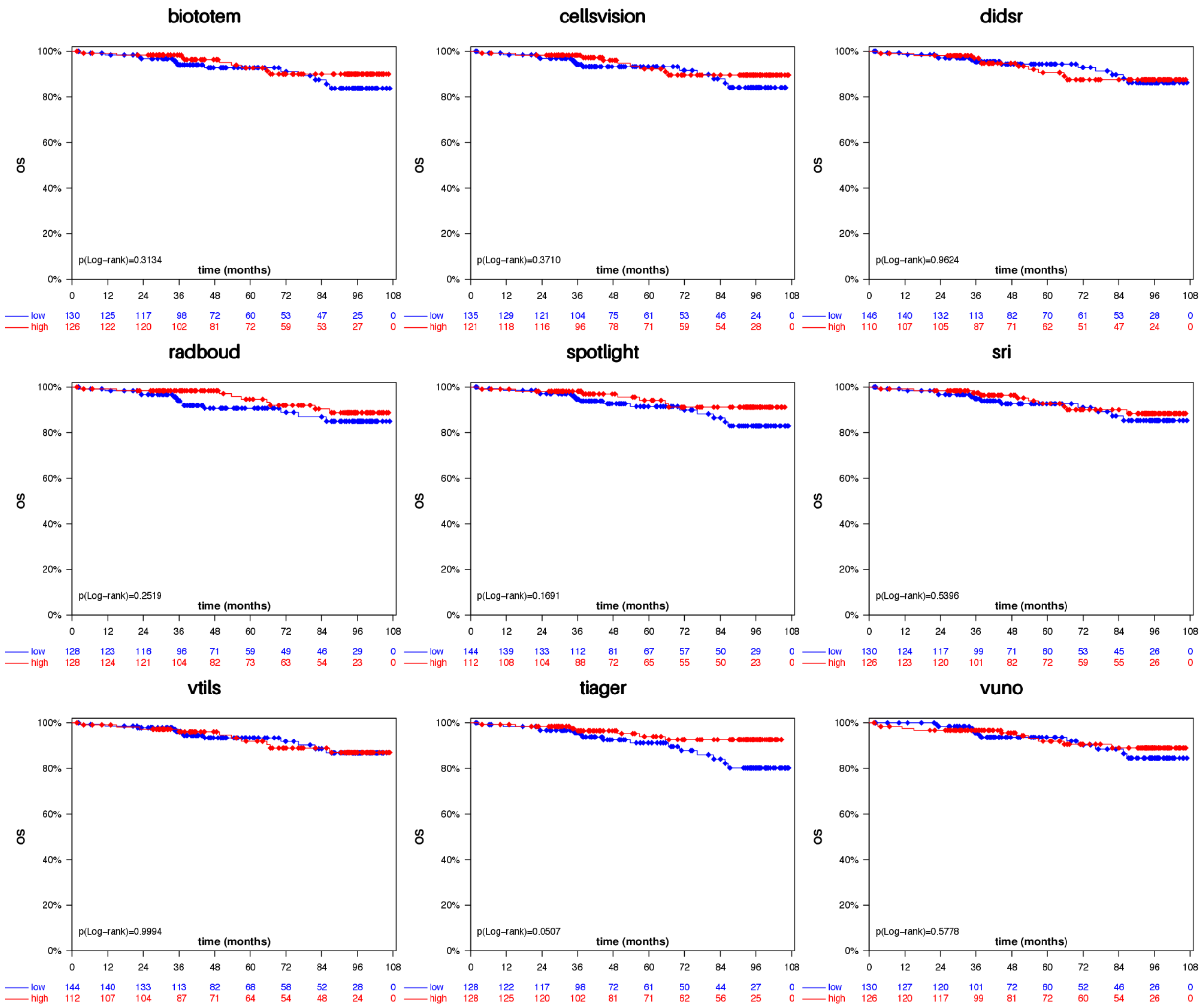


**Supplementary Figure 22:** Kaplan-Meier Curves for Overall Survival (OS) in G6 (Her2+) cohort.

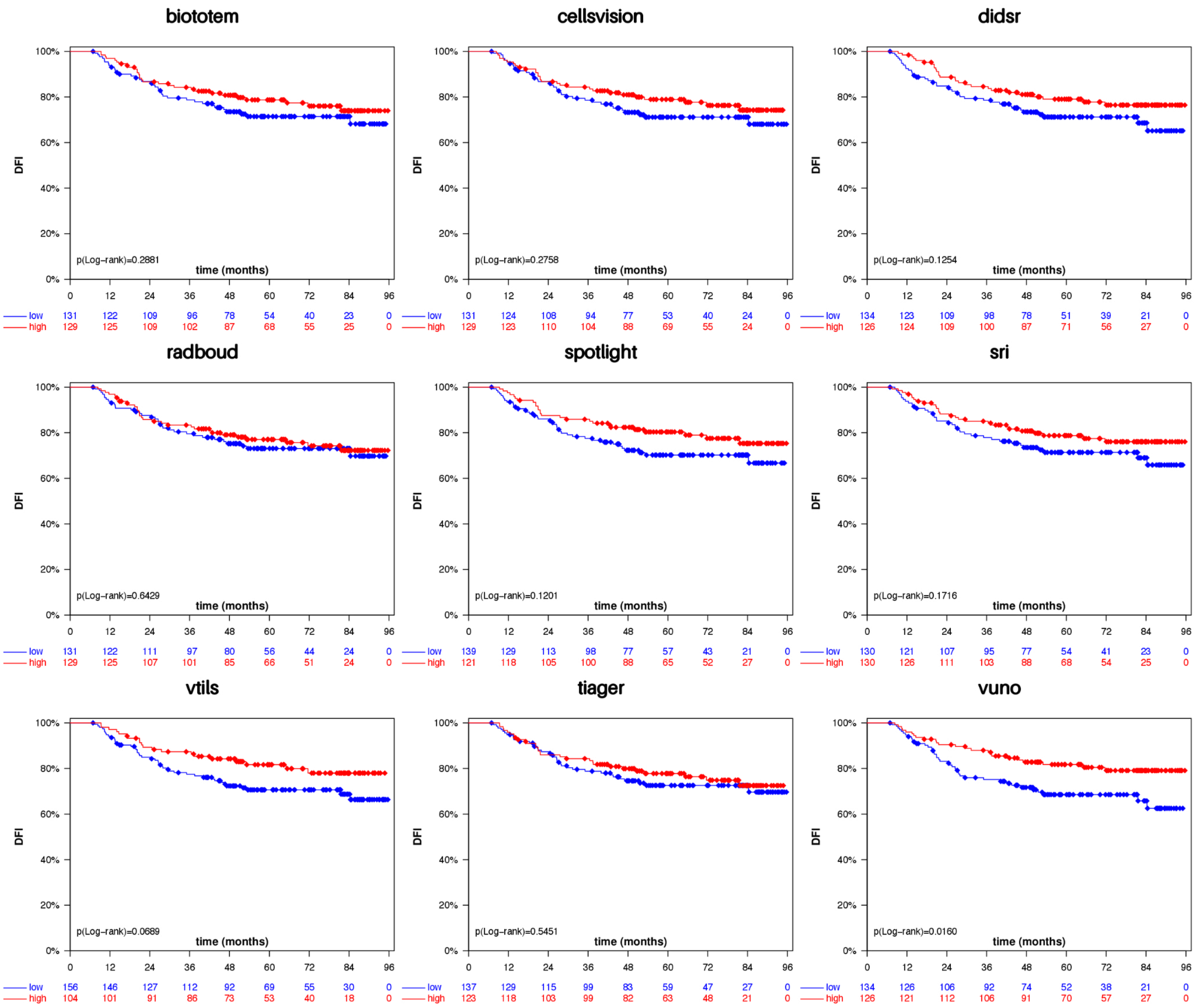


**Supplementary Figure 23:** Kaplan-Meier Curves for Disease-Free Interval (DFI) in G7 (TNBC) cohort.

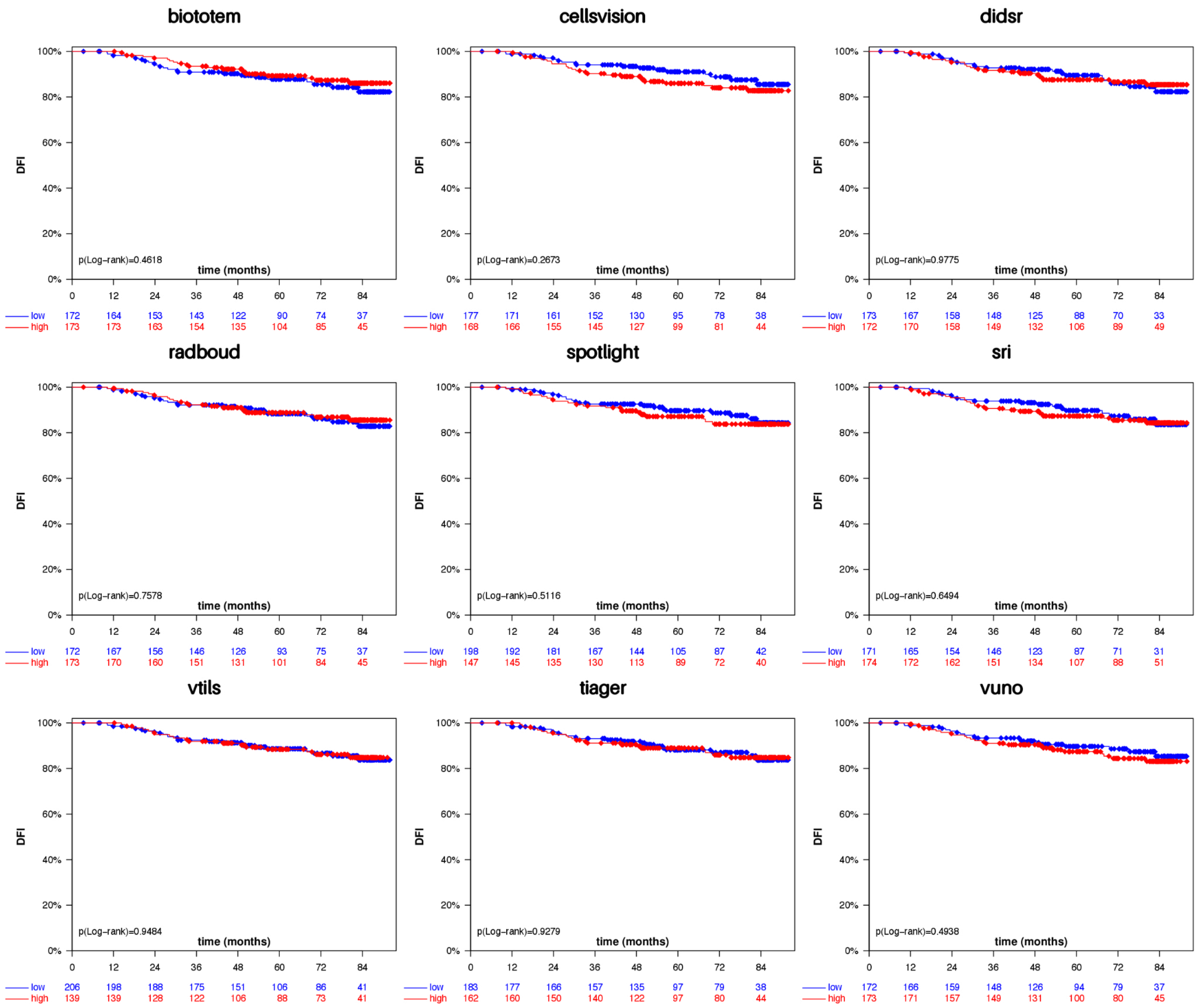


**Supplementary Figure 24:** Kaplan-Meier Curves for Disease-Free Interval (DFI) in G7 (Her2+) cohort.

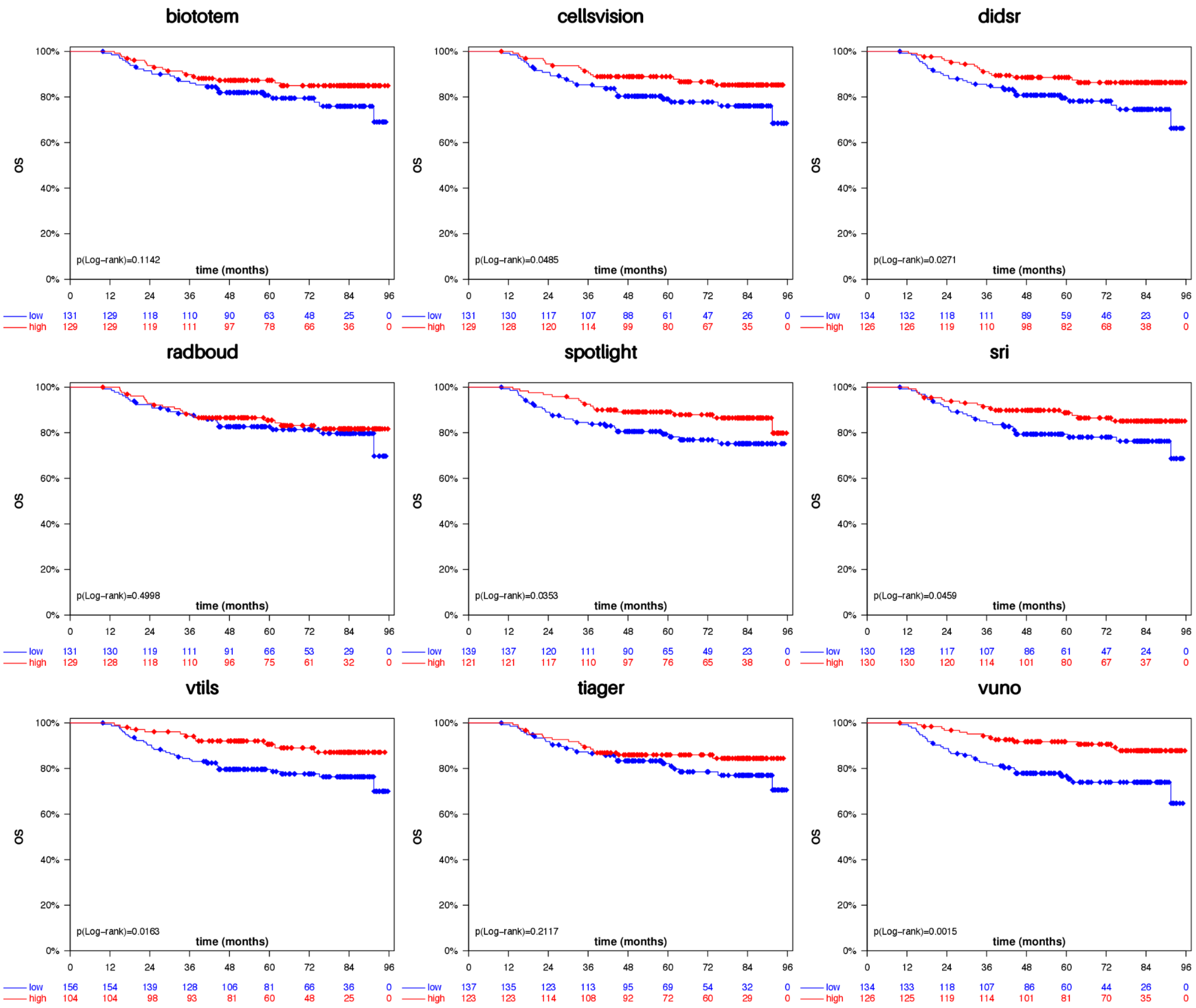


**Supplementary Figure 25:** Kaplan-Meier Curves for Overall Survival (OS) in G7 (TNBC) cohort.

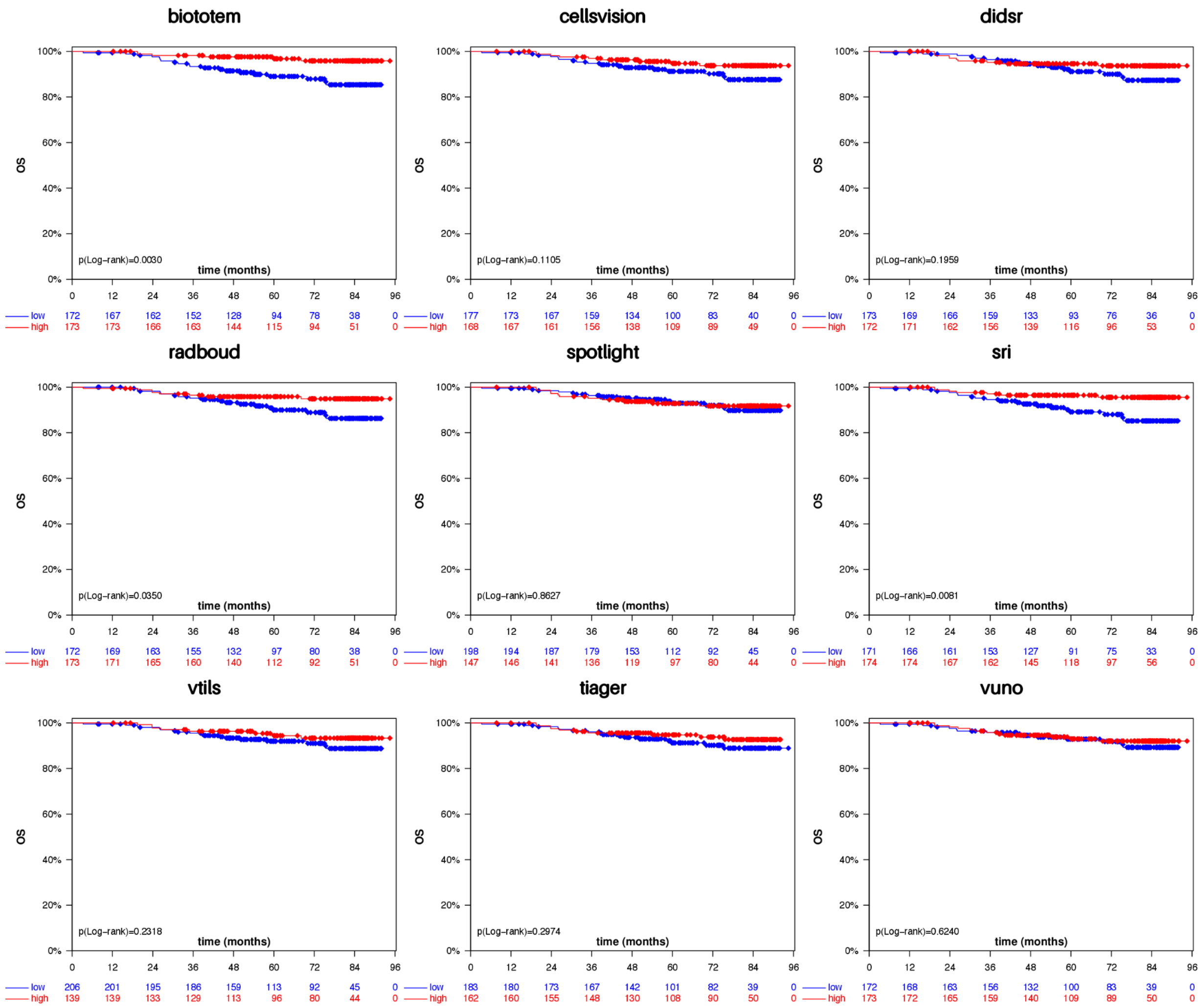


**Supplementary Figure 26:** Kaplan-Meier Curves for Overall Survival (OS) in G7 (Her2+) cohort.

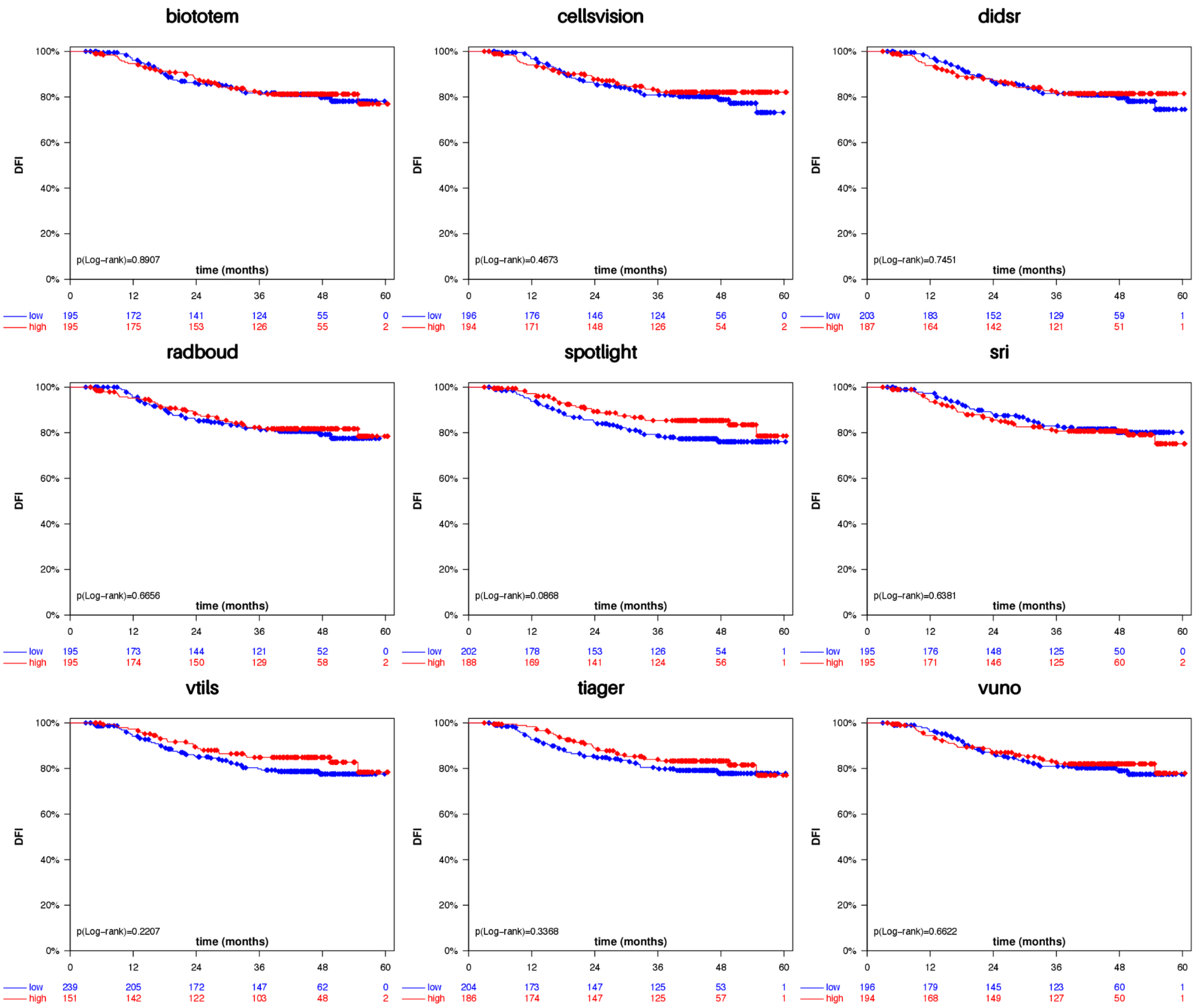


**Supplementary Figure 27:** Kaplan-Meier Curves for Disease-Free Interval (DFI) in G8 (TNBC) cohort.

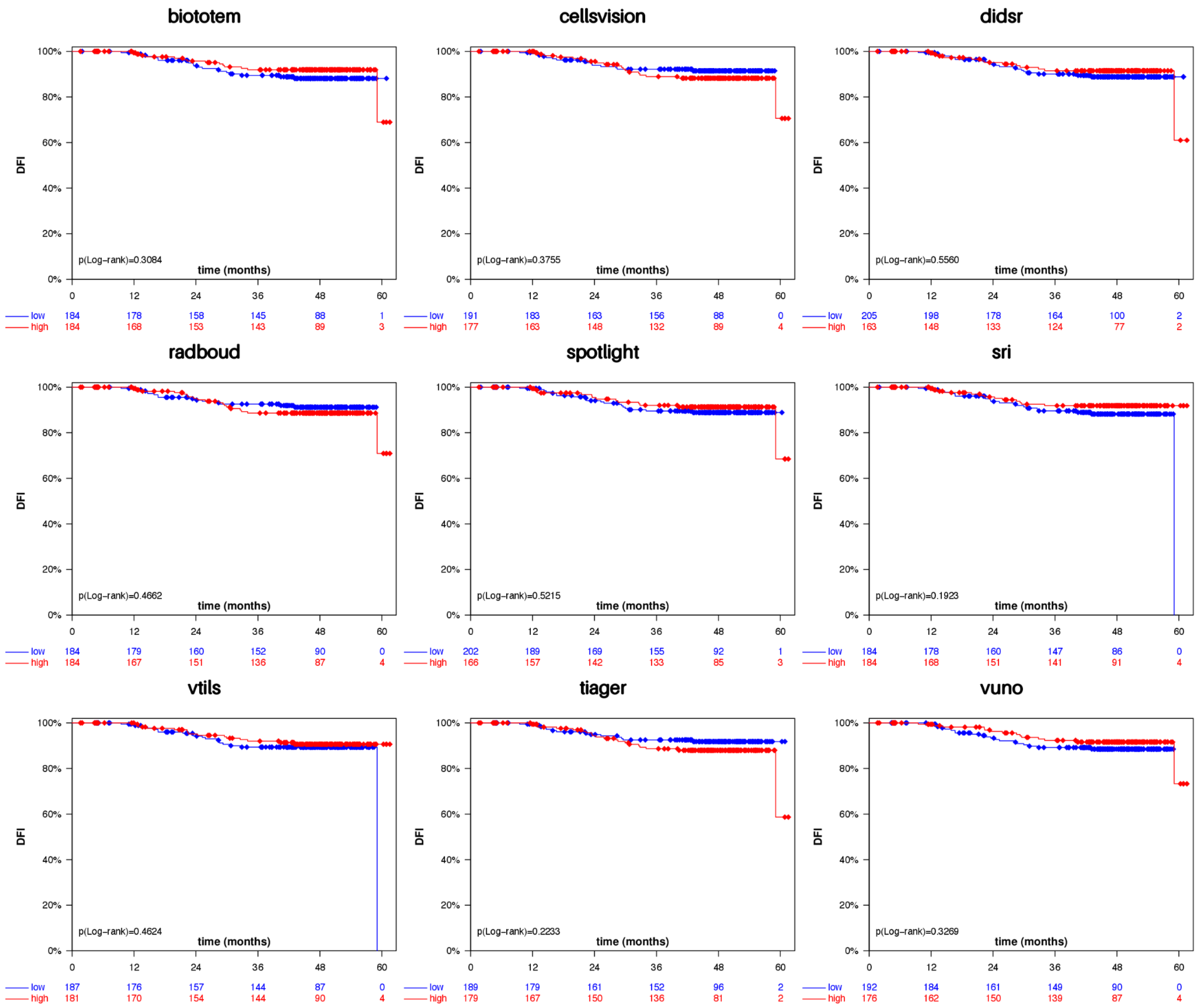


**Supplementary Figure 28:** Kaplan-Meier Curves for Disease-Free Interval (DFI) in G8 (Her2+) cohort.

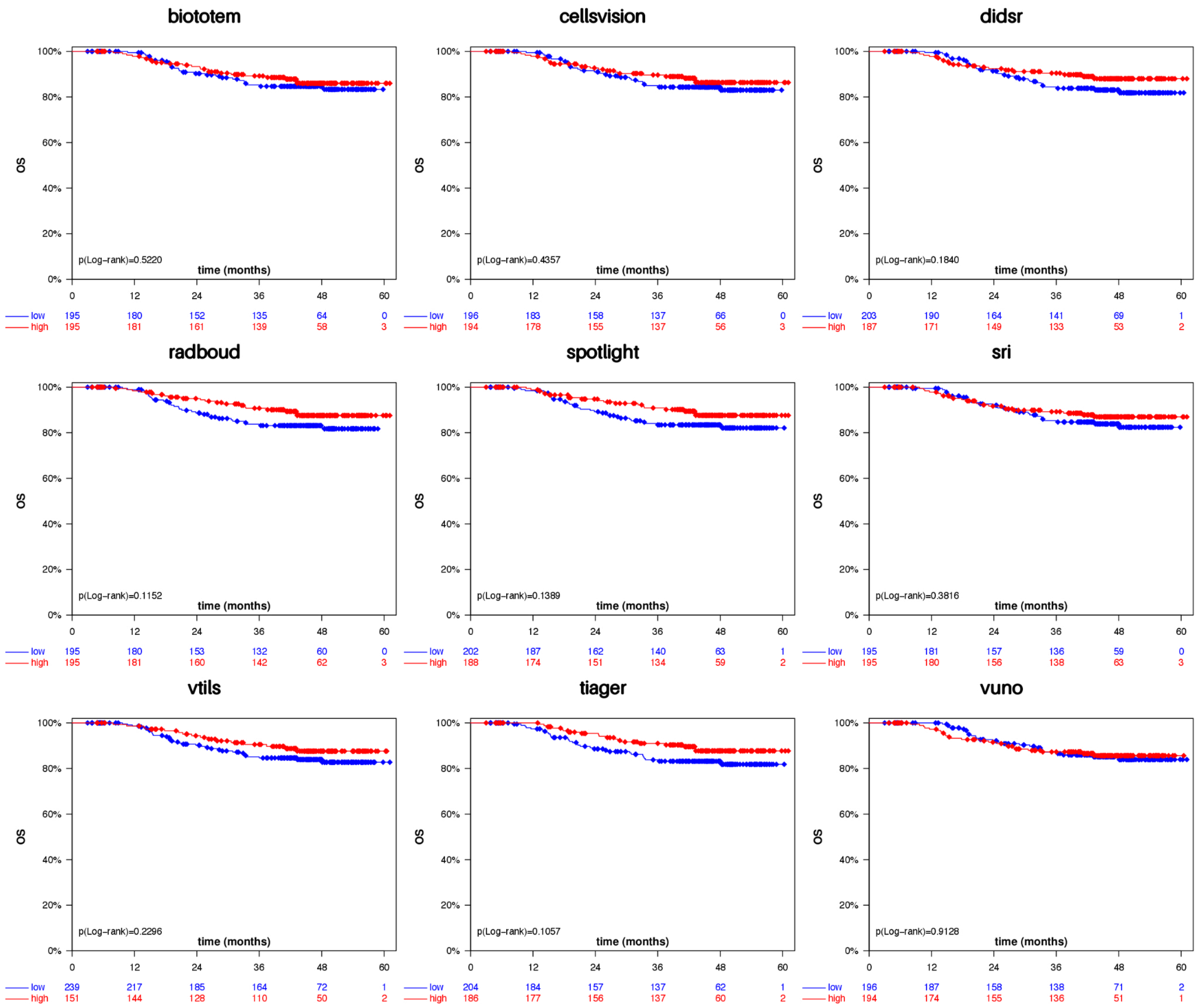


**Supplementary Figure 29:** Kaplan-Meier Curves for Overall Survival (OS) in G8 (TNBC) cohort.

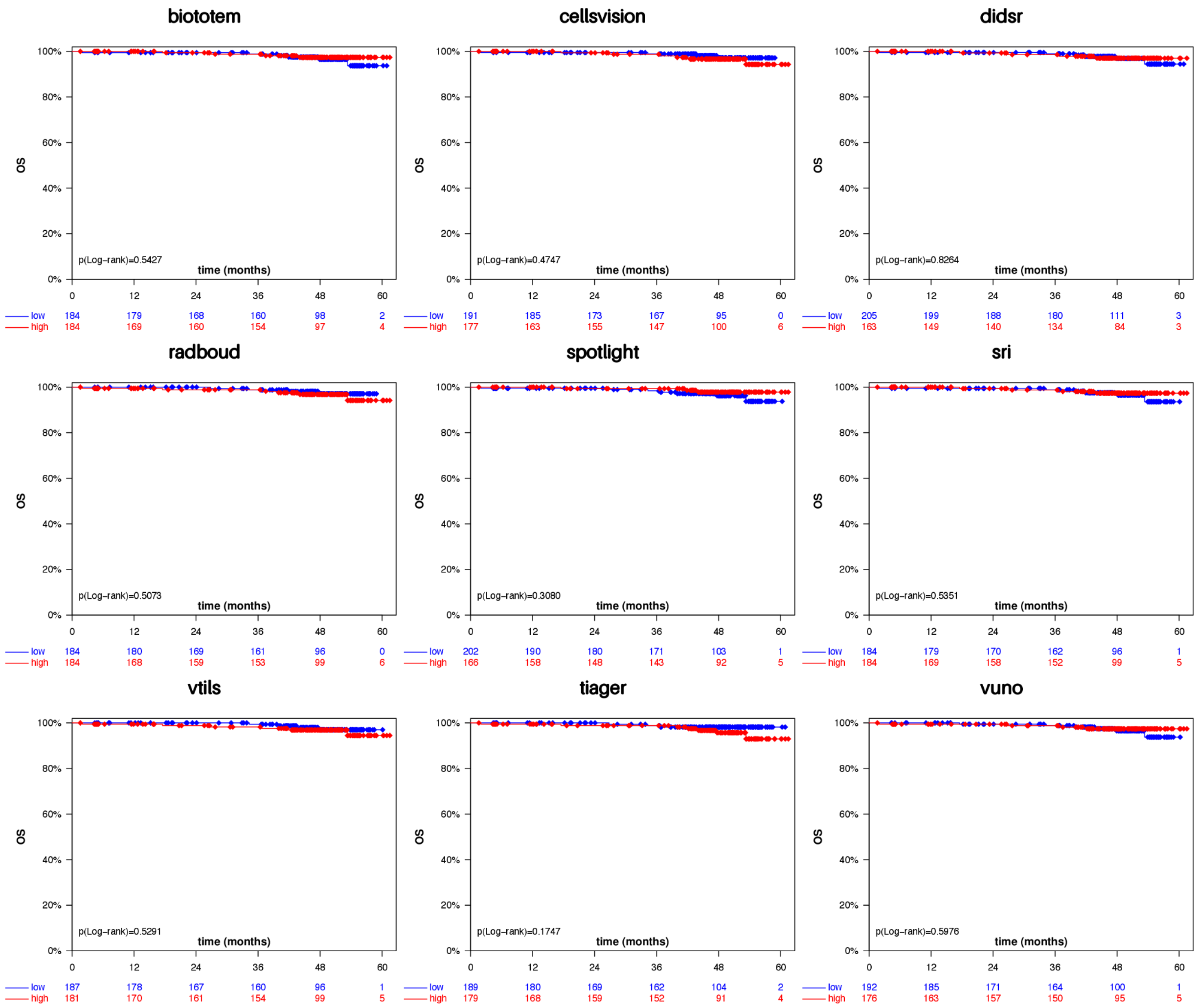


**Supplementary Figure 30:** Kaplan-Meier Curves for Overall Survival (OS) in G8 (Her2+) cohort.

#### Biopsies - Cox Proportional Hazards Analysis

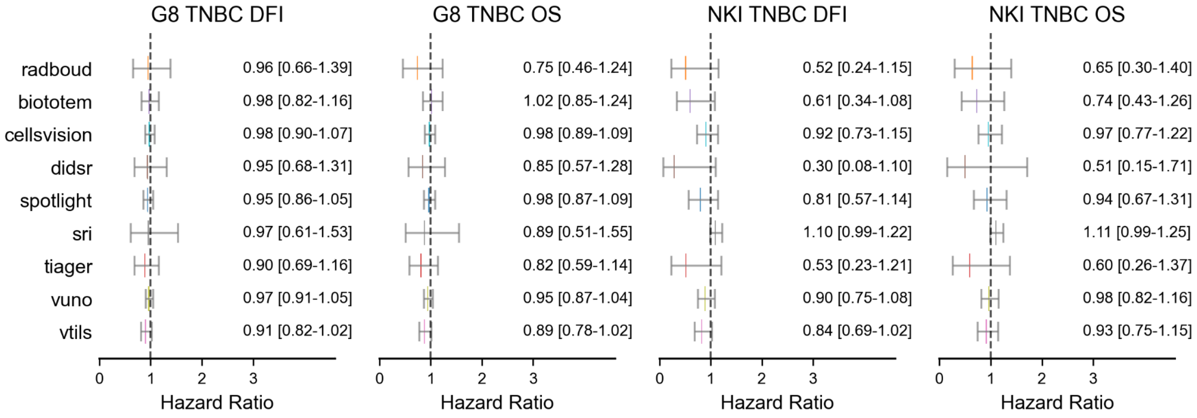


**Supplementary Figure 31:** Univariable Cox Regression analysis for DFI and OS in G8 (TNBC) and NKI (TNBC)

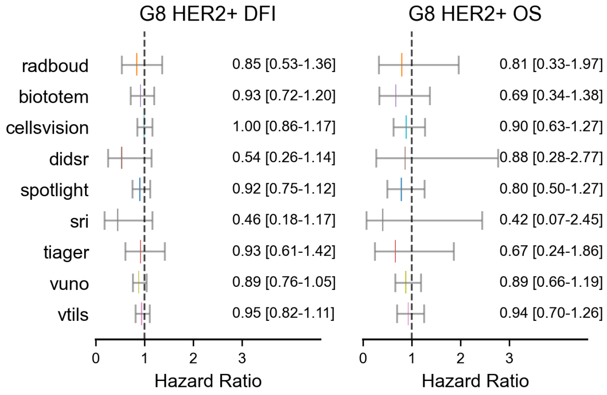


**Supplementary Figure 32:** Univariable Cox Regression analysis for DFI and OS in G8 (HER2+).

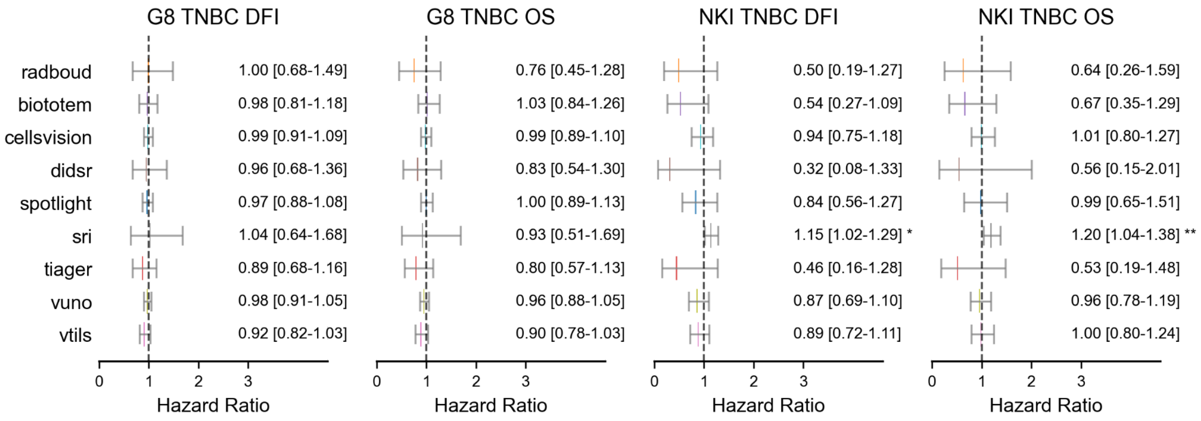


**Supplementary Figure 33:** MultiVariable Cox Regression analysis  for DFI and OS in G8 (TNBC) and NKI (TNBC).

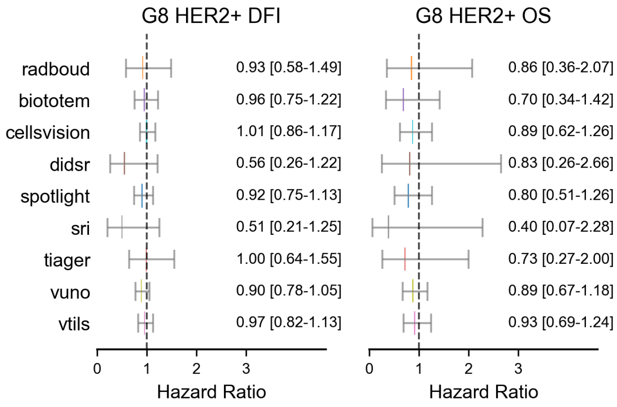


**Supplementary Figure 34:** Multivariable Cox Regression analysis for DFI and OS in G8 (HER2+).

### TIGER AI Models

### Team: Radboud

**Code and model availability**: https://github.com/DIAGNijmegen/pathology-tiger-baseline

#### Abstract

The Radboudumc team developed a baseline algorithm for the TIGER challenge, with the goal of providing a benchmark for comparison for other participants and set a lower bound for the performance of the developed algorithms. The baseline algorithm comprised three key components: a multiresolution segmentation model utilizing U-Net architecture, a Faster-RCNN object detector, and a Tumor-infiltrating-lymphocyte (TIL) scoring component.

Segmentation

##### Architecture

The baseline algorithm used the HookNet model [ref] for the segmentation task. HookNet is a multi-branch neural network that processes two concentric inputs of the same size, but with different resolutions. The branch that processes the high resolution patch is referred to as the "target branch," and is designed to capture high-resolutions features. The branch that processes the lower resolution patch is referred to as the "context branch" and enables the model to incorporate contextual information. Both branches are based on the U-net model and use a categorical cross-entropy loss function to make predictions during training. The Hooknet utilizes a hooking mechanism, inspired by the skip connections in U-net, to combine the contextual information from the context branch, with the high resolution features from the target branch.

##### Data Preprocessing and Augmentation

For the segmentation purposes the baseline used the WSI-ROIS dataset. patches and acomponied segmentation masks where extracted with the “wholeslidedata” python package which also included various augmentations such as rotation, flip, contrast, saturation, brightness augmentations.

##### Training

HookNet allows for training with a multiloss term based on the the two branches. During training a lambda of 0.75 and 0.25 percentage weres used to balance the loss for target and context branch, respectively. We followed the best settings for patch resolution depicted in theHookNet paper and trained the model with patches that have 0.5um resolution for the target branch and a 8.0um resolution for the context branch \cite{HookNet}. Other parameters for the model were also taken from the original paper. For example we used the pixel-based sampling procedure to train the model, which entailed the following strategy: the first batch was randomly sampled from the training annotations. Sampling for any subsequent batches is guided by how many pixels of each ground-truth label had been seen in previous batches. Class labels that are underrepresented in the total accumulation of pixels have a higher chance of being sampled to rebalance. A deviation from the original HookNet  the replacement of the ReLU activation function with the Leaky ReLU variant. Furthermore the HookNet was trained to predict outputs for all seven classes in the TIGER data and used 284x284x3 patches with a batchsize of 12. Training ran for 200 epochs with a 1000 batches being sampled each epoch. HookNet is engineered to use 'valid' convolutions, which result in a smaller output than input.

###### Inference

The use of valid padding eliminates any potential border artifacts, at the expense of increased computational requirements. To perform inference, we employed the "wholeslidedata" package to efficiently iterate over the test slides, ensuring that all tissue tiles were fully covered.

##### Detection

###### Architecture

For detection task in TIGER we used Faster R-CNN for lymphocyte and plasma cell detection. Faster-RCNN includes a Region Proposal Network (RPN) that predicts bounding boxes for objects and a probability that a box contains an object. The RPN is stacked on top of a Fully Convolutional Network (FCN), for which the features are shared with classifier used for the detection task.

###### Data Preprocessing and Augmentation

We trained Faster-RCNN using the WSIROIS COCO-formatted dataset, which contained images of varying sizes. To standardize the input images, we applied a fixed-size crop. If an image was larger than the crop size, we randomly sampled a patch of size from the image. For smaller images, we padded the bottom and right sides with zeros until they reached the required input shape. To increase the dataset in the training process, we applied additional augmentations to the training data, including rotations, flips, contrast adjustments, saturation changes and brightness augmentations.

###### Training

We trained the Faster R-CNN model using the Detectron2 library, \cite{wu2019detectron2}. The library provides a vast collection of pre-trained models as part of its model zoo, including various versions of the Faster R-CNN model that can be constructed using backbone models and RPNs. For our purposes, we chose the X101-FPN model, which features a combination of ResNet and a feature pyramid network (FPN) as the model backbone. This variant is the most powerful Faster R-CNN network available in the model zoo. We used the pre-trained weights that were trained on the ImageNet classification challenge. During the training, our model was trained for 200,000 steps using a learning rate of 0.000001, which was halved at pre-specified intervals of 1000, 10.000, 20.000, 50,000 and 100.000 steps.

In order to address the significant variability in image sizes within our dataset, we utilized a custom sampling method that accounts for this diversity. Typically, the standard approach in Detectron2 is to use a uniform sampler, which randomly selects images with equal probability, allowing each image to appear a uniform number of times. However, since our dataset includes images ranging from 65x65 to 1213x1249 pixels, we found that a uniform sampler would not adequately capture the full range of data available, as the larger images contain more annotations and information.

To overcome this limitation, we used the RepeatFactorTrainingSampler from the Detectron2 framework, which samples images based on their repeat factor. Specifically, the repeat factor for each image is defined as the square root of its total number of pixels. This method ensures that images with a higher repeat factor, and hence more pixels, are sampled more frequently during model training. By implementing this custom sampler, we can effectively utilize the full range of data available in our dataset, even for images with large dimensions that might otherwise be underrepresented during training

Unlike the standard training procedure in detectron, which filters out images without any annotations, we modified our training approach to include these images. This allowed us to maximize the use of our training data and enable the model to learn from non-infiltrated tissue as well. By incorporating a more diverse range of images into the training process, we aimed to improve the model's performance on WSI inference where non-infiltrated tissue is commonly encountered.

##### Inference

The Detectron module has an internal NMS component but it does not catch the same TIL that is predicted twice due to predictions in two neighbouring patches. At inference time the detection model iterates over a tissue mask with tiles of 128x128. If a part of a TIL is situated at the right edge of the tile, then the other part of that lymphocyte will be at the left edge of the next tile. The same principle applies to the top and bottom borders of each patch. If enough of the TIL is present for the model to recognise it as such then this results in the model predicting two boxes for one lymphocyte.

For this reason a second round of NMS is performed after the detection model has predicted all patches. The NMS is applied by iterating over the tissue mask used for detection. However, a patch-size of 1024x1024 is used to include any possible dual predictions made at the edges of the smaller tiles used during inference. The points produced by the detection model are converted to boxes of a fixed size with a radius of eight microns. Subsequently, the boxes are processed by a NMS function. The threshold is set to 0.7 based on empirical observations. This relatively tolerant threshold eliminates glaringly obvious dual predictions and preserves predictions in instances where lymphocytes are next to one another.

##### TIL scoring

To compute an automated TIL score based on the segmenation and detections, first a tumor tumor bulk is generated by finding the convex hull of the tumor. The tumor tissue extracted from the segmentation masks produced by Hooknet are used as the points around which to generate the shape. Our baseline method aims to emulate the practice of pathologists who manually count the proportion of tumor-associated stroma that is infiltrated by TILs. This proportion is then expressed as a percentage score. To reproduce this method using automated techniques, the total number of cells identified by Faster-RCNN is multiplied by a lymphocyte diameter of 8 µm/px. The product between the lymphocyte diameter and the total number of detected lymphocytes represents an estimate of the fraction of TILs in the tumor-associated stroma. The sum of the amount of lymphocytes detected multiplied by this constant is then taken as the area covered by TILs. To subsequently compute the TIL-score a constant of 100 is divided by the total amount of pixels covered by stromal tissue within the tumor bulk and subsequently this is multiplied by the total area covered by TILs.

### Team: VUNO

Sungduk Cho, Dong-Hee Kim, Hyungjoon Jang, Chanmin Park, Kyungdoc Kim

Affiliation: VUNO Inc., Seoul, South Korea

Contact: Kyungdoc Kim

Code and model availability: https://github.com/vuno/tiger_challenge

#### Abstract

In breast cancer, the tumor-infiltrating lymphocytes (TILs) act as an important prognostic and predictive biomarker. It requires the manual assessment of the pathologists on hematoxylin and eosin (H&E) slides, which is laborious and discordant among the readers. The Tumor InfiltratinG lymphocytes in breast cancER (TIGER) challenge aims to develop an algorithm for the fully automated assessment of TILs in H&E breast cancer slides. We, the VUNO team, have developed a novel algorithm to compute the TILs score. First, we run segmentation model on the whole slide image (WSI) at lower resolution. The most informative patches [1], which have high stroma density, are sampled from the peritumoral region. Then, we run detection model on the patches. The final TILs score is computed by using such information to consider the local and global statistics from the patches. We achieved the first place in the survival evaluation final leaderboard scoring a C-index of 0.6388 with the proposed algorithm.

#### Data

WSIROIS

- The dataset was used to train the segmentation model and the detection model.
- The dataset was divided into 5 folds based on the number of slides to gain meaningful validation results (i.e., all the patches generated from the same slide were assigned to same fold.)

WSITILS

- The dataset was used as the pseudo-label to train the segmentation model.
- Few WSIs were sampled to estimate the constant factor for the TILs score calculation.

#### Methods

##### Architecture

Fig 1. shows the overall pipeline at inference. The WSI is fed into the segmentation model to predict the tumor and stroma region. With the segmentation mask, we sampled top 20 patches with high peritumoral stroma density. Then, the detection model is run on the sampled patches to detect lymphocytes. We compute the final TILs score based on the detection result and the corresponding segmentation result.

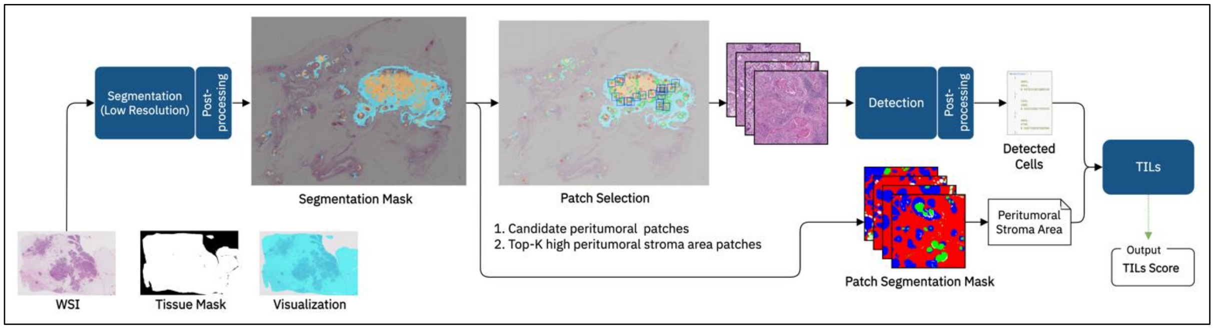


Figure 1. Overview of our TILs scoring pipeline

#### Segmentation

##### Preprocessing and augmentation

The WSI (whole slide image) was resized to resolution at 4 mpp (micron per pixel which is equivalent to 4 µm per pixel) when training the segmentation model. To alleviate the color variance among the individual slides, the means of saturation and value (luminance) on the tissue pixels were adjusted if they exceeded the preset value. The input patches were sampled from slightly wider regions than the given ROI bounds with null paddings on the label masks to guide edge pixels with sufficient neighboring pixel information. Following augmentations were applied in training: random scale, elastic transform, rotate, flip, color jitter, jpeg compression, and normalization [2].

##### Training Scheme

We used U-Net [3] with EfficientNet-B0 [4] encoder with ImageNet-noisy student as pretrained weights [5] and SCSE attention module [6] as decoder. The model semantically segmented given image into three classes: stroma, tumor, and rest. We selected each optimal epoch via 5-fold cross validation. Each fold of cross validation is optimized independently by observing tumor stroma dice, which is the official evaluation metric, over validation dataset. The cosine annealing warm restarts learning rate scheduling was used with different starting learning rates for encoder and decoder, 1e-3 and 2e-3 respectively. The stochastic weight averaging was applied with 10 epochs and learning rate of 2e-4 from 120 epochs. The model was trained for maximum 300 epochs with the Lamb optimizer [7]. The cross-entropy and the dice loss were used where the dice loss was calculated on the tumor and stroma classes only. They were summed and weighted 0.09 and 0.91 respectively.

##### Training Procedure

The segmentation model was trained in three steps. First, the WSIROIS dataset was fed into the base models. Then, the probability outputs of five different cross validated base models were mean ensembled.

Second, we trained the self-pseudo-curriculum models. The pseudo-labels were generated by predicting on the WSIROIS data with the previous base models. We used two types of pseudo-labels: 'consensus' where pseudo-labels were compared to the given GT and the mismatched areas were pruned and assigned ignore index, and 'sole' where no additional rules were applied. With the probability outputs of the pseudo-labels, we composed a self-pseudo-curriculum by applying 5 different reliability thresholds to confidence score maps as 90%, 80%, 70%, 60%, and 50%. The pixels whose confidence score were below the corresponding reliability threshold were assigned to the ignore index. The self-pseudo-curriculum dataset was fed with curriculum epochs setup as one of the following settings: [0, 10, 30, 60, 100], [0, 10, 20, 40, 80], [0, 10, 20, 30, 40]. Finally, the five different models were mean ensembled

Third, we trained the semi-supervised models. Several patches were sampled per slide at the peritumoral area. The pseudo-labels were generated by predicting on WSITILS dataset with the self-pseudo-curriculum models. In this time, only sole-type pseudo-labels were generated. Then, the same training procedure was repeated as the self-pseudo-curriculum models.

###### Post-processing

Several post-processing methods were applied to reduce the false positives on tumor predictions. The tumor class probability was suppressed over the region where the tumor class and the 'rest' class are briskly competing against each other (i.e., the tumor pixels whose confidence score is lower than 80% after applying temperature of 1.5). Suppressing tumor predictions in small size was also found effective.

##### Detection

###### Training

First, we trained a model using WSIROIS dataset with the detection annotations. Then, we trained the model by semi-supervised learning. The WSIs in WSIROIS dataset with the segmentation ground truth were used. The pseudo-labels were postprocessed with the segmentation result. We cropped image into 160x160 patches if the size of image was greater than 300x300 and used as given without cropping otherwise.

We used YOLOv5 [8] model with COCO pretrained. 5-fold cross validation was used. Linear learning rate scheduling was used with AdamW [9] optimizers. The model was trained for maximum 500 epochs with Bayesian hyperparameter optimization and early stopping. We monitored mAP (mean average precision), FROC (Free-response Receiver Operating Characteristic), and recall at precision 0.95 while training. The augmentations were applied as follows: HSV, shear, flip, mosaic, mix-up, blur, and CLAHE. Finally, seven models of different hyperparameters and different model sizes were ensembled.

###### Post-processing

We implemented NMS (non-maximum suppression) based on the center point distance instead of the existing NMS based on the IoU (intersection over union). The key point of our post-processing scheme is to increase the confidence of evident points and lower the confidence of ambiguous points. That is, the confidence is decreased if less than 2 detectors failed to detect certain point and vice versa. To focus on the stroma area, the points detected inside the tumor or rest class area of the segmentation result had their confidence lowered or were deleted.

#### TIL scoring

We estimated TILs score based on the local and global statistics from the sampled patches in the peritumoral region. First, we sampled 50 candidate patches along the tumor boundary in the segmentation results, and then defined the peritumoral stroma region around the tumor boundary by dilating the tumor region and overlapping it with the stroma mask. Note that the width of resulting region with respect to the tumor boundary is approximately 100 µm. Finally, we chose 20 patches with the high stroma density (i.e., the stroma pixels to the total pixels in a peritumoral region) as informative patches inspired from [1]. The detection model is run on the sampled patches.

Next, we computed the statistics related to TILs with the sampled patches 1) the patch-wise TILs score as a local statistic, and 2) the WSI-wise TILs score as a global statistic. To get the patch-wise TILs score, the ratio of the number of lymphocyte cells to the stroma pixel count was computed per patch and then we averaged the ratios of the sampled patches (Eq 1). To get the WSI-wise TIL score, the number of the lymphocytes cells was summed up and divided by the sum of stroma pixel count of the sampled patches (Eq 2). Finally, the TILs score is estimated by averaging of local and global TIL score, multiplying a constant factor for scaling, and clipping the score 1 to 95 (i.e., minimum to maximum) (Eq 3). The constant factor was estimated using WSITILS dataset.

$$(1)\ \text{patch} - \text{wise TIL score} = \frac{\Sigma \frac{\text{detected lymphocyte cells}}{\text{peritumoral stroma area}}}{\text{number of patches}}$$

$$(2)\ \text{WSI} - \text{wise TIL score} = \frac{\Sigma\, \text{detected lymphocyte cells}}{\Sigma\, \text{peritumoral stroma area}}$$

$$(3)\ \text{TILs score} = \text{MinMax}(1, 95, \text{constant factor} \times \frac{(\text{patch} - \text{wise ratios} + \text{wsi} - \text{wise ratio})}{2})$$

[9] Ilya Loshchilov, Frank Hutter. “Decoupled Weight Decay Regularization.” ICLR (Poster) 2019

### Team: Spotlight Pathology

Anna Maria Tsakiroglou, Martin Fergie, Richard Byers
Affiliation: Spotlight Pathology Ltd., Manchester, United Kingdom

Contact: Martin Fergie
Code and model availability:
https://github.com/Spotlight-Pathology/spotlight-tiger

#### Abstract

Higher density of tumour-infiltrating lymphocytes (TILs) is associated with improved survival in breast cancer patients; however, pathologist TIL scoring is based on approximate estimates and subject to inter-observer variability. The Tumor InfiltratinG lymphocytes in breast cancER (TiGER) challenge aims to develop algorithms for automated TIL scoring and assess their prognostic value. For this challenge, as the Spotlight Pathology team, we have developed an algorithm to a) segment tumour vs. stroma using an ensemble of U-net networks, b) identify the tumour bulk area, c) detect TILs with Faster-RCNN and d) produce a score per patient from the TIL density in the tumour associated stroma. On the final test set our approach achieved a tumour-stroma segmentation Dice score of 0.746 and a FROC score of 0.474 for lymphocyte detection. This approach ranked 2nd on the final leaderboard for survival prediction with a C-index of 0.622.

#### Data

##### Used Datasets

- WSIROIS: All ROIs and corresponding annotations were used to train and test the lymphocyte detection (tissue-cells) and segmentation models (tissue-cells and tissue-bcss).
- WSITILS: Used to build a model to map the predicted TIL area ratio to the known pathologist TIL score.
- WSIBULK: Used to enrich the “other” segmentation class by randomly sampling patches from the non-tumor-bulk areas.

##### Data Split

Models were trained with 3-fold cross validation to internally estimate the generalization error. We ensured that patches from different slides were used for training, validation and testing.

### Methods

#### Preprocessing

Patches were dynamically extracted from the ROI-level datasets during training using a sliding window approach. Patches of 256 x 256 were used for the detection task and 224x224 for the segmentation task, and normalized to ImageNet mean and standard deviation. ROIs smaller than the selected patch size were padded with a constant. Segmentation mask classes were combined into invasive tumor, stroma (including tumor-associated and inflamed), other (including in-situ tumor, healthy glands, necrosis and rest) and non-annotated.

#### Augmentation

We augmented the data using vertical, horizontal and 90-degree rotations, as well as random brightness, contrast, gamma permutations and color jitter. Random small permutations of resolution were added, to account for the fact that the first pyramid level in WSIs varied in resolution with an overall approximate resolution of 0.5 microns / pixel. Augmentations were applied on the fly on a different randomly sampled subset of images per epoch.

#### Segmentation-Model

For segmentation we used an ensemble of two multi-class U-net [1] type models trained at different resolutions, one at maximum resolution (~0.5 microns / pixel) and one downscaled 4x (~2 microns / pixel). The models had a ResNet-101 [2] and a Densenet-121 [3] backbone, respectively, pretrained on ImageNet, which were kept frozen as feature extraction. While performance was adequate at the highest resolution, adding the downscaled level increased the field of view of the model, allowing better discrimination for cases where context was important (e.g., in-situ tumor).

We trained the segmentation model using the Adam optimizer with a starting learning rate of $10^{-3}$, a momentum of 0.9, weight decay equal to $5 \cdot 10^{-4}$ and batch size of 20. Linear learning rate warmup was applied during the iterations of the first epoch to avoid early overfitting and stabilize training. Bayesian optimization was used to tune the training hyperparameters, such as the starting learning rate, proportion of training instances augmented, backbone architecture and patch resolution. Patches containing challenging tissue phenotypes, such as in-situ tumors and healthy glands were oversampled for better representation during training. Additionally, a weighted cross entropy loss was used for training to balance the contribution of each of three classes (invasive tumor, stroma and other). Pixels marked as non-annotated were ignored when calculating the loss. We trained for a maximum of 50 epochs and early stopping was applied by observing the validation loss.

#### Detection-Model

For detection we used a two stage Faster-RCNN object detector with a FPN-ResNet-50 backbone [4]. The model predicted an "objectness" score and a regressed bounding box for each detected TIL. The backbone of the detector was pre-trained on COCO and fully unfrozen. Non-maximum suppression (NMS) at post-processing with an intersection over union (IoU) threshold of 0.4 was applied twice, at patch and WSI level.

We trained the detection model using the Adam optimizer with a starting learning rate of 10-5, a momentum of 0.9, weight decay equal to $5 \cdot 10^{-4}$ and batch size of 5. The learning rate decayed exponentially when a training plateau was reached for at least 3 consecutive epochs by gamma equal to 0.9. We used the classic two-stage detector loss [5], a sum of the region proposal network (RPN), classifier and regression head losses. Bayesian optimization was used to tune the training and model architecture hyperparameters, such as the patch size, anchor size, backbone, starting learning rate, and NMS IoU threshold during post-processing. Models were trained for a maximum of 50 epochs and early stopping was applied to avoid overfitting. Patches without any lymphocytes were undersampled during training by a factor of 10%, as we wanted to include some representative "empty" tissue areas without creating a very imbalanced dataset where lymphocytes would be severely underrepresented compared to other cells.

#### Post-processing

To run inference on a WSI, large non-overlapping image regions (> 1024 pixels) are read consecutively and tiled into small patch sizes expected by the models. The predictions from the small patches are then merged to form the output for the large image region which is then written to file. This two-step tiling approach was adopted to minimize the number of expensive read/ write operations to/ from the WSI file format. The large image regions are tiled into smaller patches in a sliding window fashion with overlap and a stride of 200 pixels (same for L1 and L2). The edges of inferred patch masks are discarded to overcome artefacts from using "same" padding on convolutional layers of the segmentation model. Overlapping patch predictions are merged by averaging the logit scores.

There are two differences in post processing between L1 and L2:

- the size of the large non-overlapping regions (1024 x 1024 pixels for L1 and 2048 x 2048 for L2)
- in L2 lymphocyte detection is only carried out in the segmented tumour-associated stroma inside the tumour bulk region, while in L1 lymphocyte detection is carried out in all tissue regions defined by the tissue mask.

#### TIL Scoring

First the tumor and stroma segmentation for each slide is computed. Secondly, the tumour bulk area is identified by finding a concave hull around the tumour using the Radboud implementation [6]. Thirdly, the object detector locates all TILs. For speed, and because parallelization was not available on the Grand-Challenge platform, a maximum of 180 non-overlapping regions of size 2048 x 2048 were randomly sampled from the tumour associated stroma enclosed within the tumour bulk area to perform TIL detection. This mimics the process followed by pathologists when scoring large WSI.

The total TILs area is calculated assuming an average equivalent diameter of 8 microns per TIL, and finally, the ratio of TIL area / stroma area is fed as input to a linear regression model to predict the final pathologist score [0-100]. The linear model was built with the entire WSITILS dataset where the pathologist score was known by performing a type of regression robust to outliers (Huber regression [7] with l=1.9).

For cases where the tumour bulk was not found, either because no tumour was present on the WSI or due to model errors, the TIL score is undefined, and these values were imputed using the median score of all WSITIL cases.

### Team: FDA-CDRH-OSEL-DIDSR

Arian Arab, Victor Garcia, Brandon D. Gallas, Nicholas Petrick, Weijie Chen

Division of Imaging, Diagnosis and Software Reliability, U.S. Food and Drug Administration, MD, USA.

Contact: Arian Arab

Code and model availability: https://github.com/DIDSR/DIDSR-TiGER

#### Abstract

Tumor-infiltrating lymphocytes (TILs) are shown to be of predictive and prognostic importance in breast cancer patient survival. In clinic, pathologists' visual assessment of biopsies and surgical resections of human epidermal growth factor receptor-2 positive (HER2+) and triple-negative breast cancers (TNBC) results in a quantitative score of the TILs density (TILs-score). The TIGER challenge is one of the first public challenges on artificial intelligence based computerized TILs-scoring algorithms in hematoxylin and eosin-stained (H&E) breast cancer slides. We participated in the TIGER challenge by designing algorithms for tissue segmentation, cell detection, and TILs-scoring. Our algorithms estimate a TILs score between 0 and 100 by calculating the proportion of TILs area within the tumor-associated stromal area. Our method ranked 5th in the final leaderboard of the TIGER challenge with a concordance index (c-index) of 0.60 (95% CI [0.37, 0.78]).

#### Data

We used the data provided by the organizers of the TIGER challenge to develop our segmentation, detection, and TILs-scoring algorithms. For leaderboard-1 we used the WSIROIS data (which is a subset of the data provided by the organizers of the TIGER challenge) to develop the segmentation and detection models. For leaderboard-2, we used the predictions from our segmentation and detection models to design a whole slide image (WSI)-based TILs-scoring algorithm. The data annotated in the TIGER challenge is from three sources: Radboud University Medical Center in Netherlands (TC); Jules Bordet Institute from Belgium (JB), and the Genomic Data Commons Data Portal (TCGA). The tissue segmentation masks include 6 different compartments (invasive tumor, tumor-associated stroma, in-situ tumor, healthy glands, necrosis not in-situ and inflamed stroma) with label values from 1 to 6. Anything that does not fall into the above-mentioned categories is labeled as the "rest" class with the label value of 7. Some parts of the ROIs are left without any annotations and those correspond to the label value of 0. The cell detection annotations mark the centroid location of the TILs ($x_c$ and $y_c$). The cloud platform of the TIGER challenge contains hidden test data to rank the segmentation, detection, and TILs-scoring algorithms. We did not have access to this dataset, and we needed to deploy our models (stored in a Docker file) to obtain results for our segmentation, detection, and TILs-scoring algorithms. During the development phase of the TIGER challenge, we could run our algorithm on a portion of the hidden test data which is called the experimental holdout test set. After tuning and obtaining the best model, we ran this model on the final holdout test set to obtain the final results for our segmentation, detection, and TILs-scoring algorithms.

The TCGA dataset consisted of 151 slides. All 151 slides had a large region of interest (ROI) measuring approximately 2000x2000 pixels that was annotated with segmentation masks. 121 of these 151 slides also had several additional smaller ROIs measuring approximately 100x100 pixels that were annotated with both segmentation masks and cell detection marks. Figure 1 shows an example slide from the TCGA dataset with both large and small ROIs and the corresponding annotations.

Our model's input size for both the segmentation and cell detection models is 256×256 pixels. We trained our segmentation model using only patches from the larger ROIs. For our detection model, the TILs detection annotations are given on the smaller ROIs of size 100×100 pixels. As a result, we manually extended the smaller ROI to a size 500×500 pixels from the tiff image to obtain patches of size 256×256 pixels. An example of the manually extended ROI along with the original smaller ROI is shown in Figure 2. We should also note that some of the ROIs in the TCGA dataset were originally rotated. We used the metadata information in the xml files to correct for these rotations. An example of such rotations and the corrected mask is shown in Figure 3.

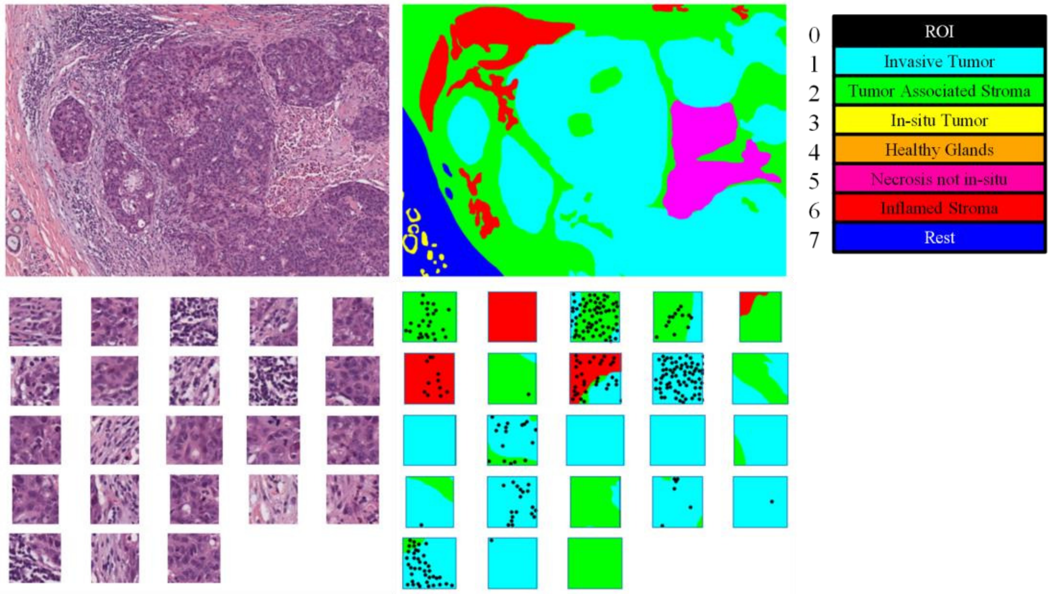


Figure 1. Top row shows the larger ROI of size 2000×2800 pixels (left column) along with the segmentation mask highlighting different tissue compartments (middle column). The remaining rows show the smaller sized ROIs of about 100×100 pixels from the same slide (left column) annotated with segmentation masks and cell detection marks (middle column). The black circles mark the centroid location of the annotated TILs. The color-coded legend (right column) identifies different tissue compartments with the mask values.

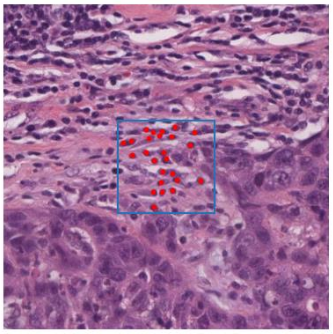

Figure 2. The manually extended ROI of size 500×500 pixels extracted from the tiff image is shown along with the original annotated ROI of size 100×100 pixels shown with the blue colored bounding box. 256×256 pixels patches were then extracted as inputs to our model for algorithm training with the TCGA dataset.

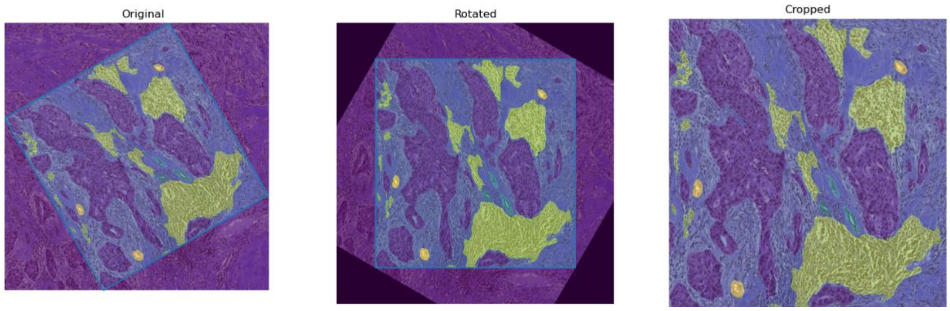


Figure 3. Left: The original ROI with the corresponding mask. Middle: The algorithm identifies the degree of rotation automatically and rotates both the image and the mask to the upright position. Right: The corrected image and corresponding mask after cropping the regions which were not annotated originally.

For the TC and JB datasets, the segmentation and detection annotations are given on the same sized ROI of about 1000×1000 pixels. The TC dataset has, 26 slides with 81 ROIs annotated for both segmentation and detection. Twenty-five slides have three annotated ROIs, and one slide has six ROIs. The JB dataset has 18 slides with 54 annotated ROIs. Each slide has three ROIs that are annotated for segmentation and detection. Figure 4 shows example ROIs from the TC and JB datasets.

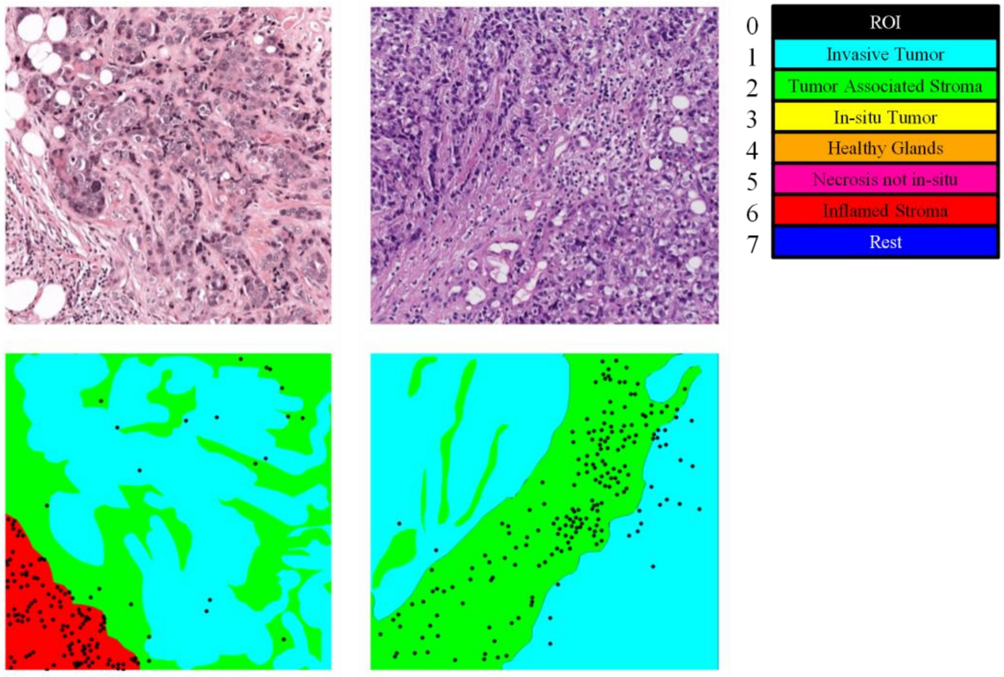


Figure 4. The left column shows an example ROI from the TC dataset (about 1000×1000 pixels) along with the segmentation mask and the centroid location of the annotated TILs shown with the black colored circles. The middle column shows an example ROI from the JB dataset (about 1000×1000 pixels) along with the segmentation mask and the annotated TILs. Finally, the color-coded legend (right column) identifies different tissue compartments for the mask values.

### Methods

#### Algorithm Deployment

In order to run our models on the experimental and final holdout test sets, we used the TIGER -algorithm-example provided by the challenge organizers. The tiger-algorithm-example contains codes to run the segmentation, detection, and TILs-scoring algorithms on the whole slide images (WSI). The first step is to extract patches from the whole slide images. This can be done in two ways:

1) Using the *tile-by-tile approach* with a stride equal to the tile's size which results in patches without an overlap between any pair of the extracted patches from the WSI.
2) Using the *center-crop approach*. In this approach, the stride between consecutive sliding windows is half the sliding window size.

The center-crop approach is intended to remove segmentation artifacts from model's predictions at the border of the patches. By extracting patches with strides equal to half of the patch size, we can potentially correct these segmentation artifacts. The center-crop approach was successful when applied to leaderboard-1; however, it failed when applied to leaderboard-2 due to the 2-hour limit constrained by the computational platform for leaderboard-2. The time to process the entire WSI for the center-crop approach is twice the tile-by-tile approach. As a result, for our final model, we used the tile-by-tile approach to predict segmentation, detection, and TILs-scores for the WSIs on the holdout test sets.

#### Segmentation Algorithm

##### Preprocessing and Augmentation

To design a TILs-scoring algorithm, we need to segment the tumor and the stroma regions. Hence, we relabeled the training masks into tumor, stroma, and other (rest class) regions as shown in Figure 5.

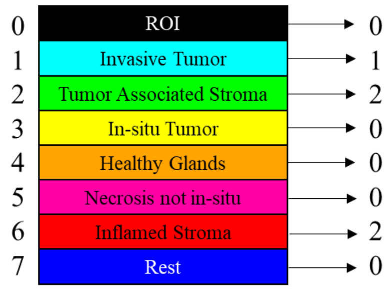


Figure 5. The legend identifying different tissue compartments with the mask values. The original annotation labels (0-7) on the left are re-labeled to only 0-2 (on the right) to train our segmentation algorithm, which is intended to segment the tumor (1) and stroma (2) regions.

We then extracted patches of size 256×256 pixels from each of the ROIs of the TCGA, TC, and JB datasets by using a tile-by-tile approach with a window of size 256×256 pixels and a stride of 256 pixels. We should note that there might be partial overlaps between the extracted patches at the borders of the ROIs due to the fixed stride of 256 pixels. To augment the number of training patches, we then rotated the original ROIs with angles of +45 degrees and -45 degrees and extracted additional patches using the sliding window technique as before. To augment the patches spatially, we used the D4 symmetry group of a square. Each patch is augmented eight times following the D4 symmetry group of a square: Original, Rot(90), Rot(180), Rot(270), fliplr, flipud, fliplr(Rot(90)), flipud(Rot(90)). We then randomly shuffled all the extracted and augmented patches.

##### Model architecture

The segmentation model we used is a U-Net model with an InceptionV3 backend. ImageNet pretrained weights are used to initialize weights and biases [1]. We also normalized the training patches RGB values ranging from 0 to 255 to -1 to 1. The inputs to the U-Net model are patches of size 256×256×3, and the outputs are 256×256×3 masks. For the output mask, the first channel is the rest-class, the second channel is the tumor class, and the third channel is the stroma class. We added a dropout layer before the output SoftMax layer to avoid overfitting.

##### Training

To train the U-Net model, a compound loss function of Dice Loss and Categorical Focal Loss is used. ADAM is chosen as the optimizer with a fixed learning rate of 0.0001. We trained the model for 30 epochs with a batch-size of 32. When tested on the experimental holdout test set, our model resulted in a Dice score of 0.7372 for the tumor class and 0.7513 for the stroma class. On the final holdout test set, our model achieved in a Dice score of 0.7056 for the tumor class and 0.7718 for the stroma class.

#### Cell Detection Algorithm

##### Preprocessing and Augmentation

For the detection model, we used the TCGA, TC, and JB datasets. For TCGA dataset, as explained before, we manually extracted a larger ROI of size 500×500 pixels from the original tiff images. In total, there are 1744 ROIs with 20,727 TILs annotations from the TCGA dataset, 81 ROIs and 54 ROIs with 4,728 and 5,523 TILs annotations for the TC and JB datasets respectively. We then combined all the ROIs from each of the data sources and split the ROIs from the TCGA, TC, and JB datasets into training/tuning datasets as shown in Table 1.

| Datasets | Number of training ROIs | Number of training TILs | Number of tuning ROIs | Number of tuning TILs |
|---|---|---|---|---|
| TCGA | 1578 | 18585 | 166 | 2142 |
| TC | 76 | 4456 | 5 | 272 |
| JB | 49 | 4891 | 5 | 632 |
| Total | 1703 | 27932 | 176 | 3046 |

Table 1. Number of training and tuning ROIs along with the number of annotated TILs for each of the TCAG, TC, and JB datasets.

To extract training and tuning patches from the TCAG, TC, and JB datasets, we did the following. For each ROI in the TCGA dataset, we first extracted a patch of 256×256 pixels at the center of our manually extended ROI of size 500×500 pixels. We then extracted eight additional patches from the same ROI with a stride of 32 pixels with respect to the patch at the center as shown in Figure 5. For the TC and JB datasets, we used the sliding window technique with a window size of 256×256 pixels and stride of 128 pixels.

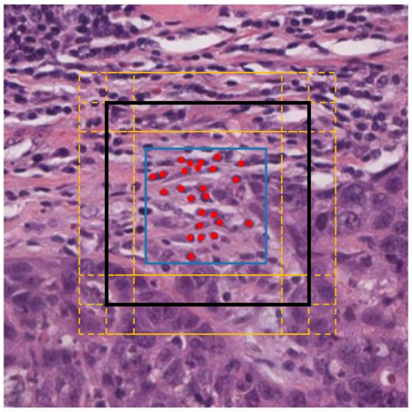

Figure 5. Outlines of extracted training and tuning patches from ROIs in the TCGA dataset for development of our cell detection algorithm. This figure shows a manually extended 500x500 pixel ROI with the blue box outlining the original 100x100 pixel provided in the dataset. The black box represents the 256x256 pixel patch extracted from the center of the ROI. The yellow boxes represent eight additional ROIs of size 256x256 pixels. Each of these eight patches were extracted using a stride of 32 pixels with respect to the center patch. The solid yellow lines represent shared patch edges, while the dotted yellow lines represent unshared patch edges.

The number of training and tuning patches used for algorithm development is shown in Table 2.

| | Training Set | Tuning Set |
|---|---|---|
| Total number of Patches | 23654 | 2296 |
| Total number of TILs | 199233 | 22411 |

Table 2. Total number of patches and number of TILs for the training and tuning sets after patch extraction.

For the training dataset, we augmented the patches by flipping the original patches left/right, up/down, and also taking the transpose of each of the patches. This way, the number of patches in the training dataset were augmented four times. For the tuning dataset, we didn't augment the patches; instead, we randomly flipped the original patches left/right, up/down or transposed each of the original patches. After these augmentations and random flips, we shuffled the patches randomly.

#### Model architecture

The detection model we used is a U-Net model with an InceptionV3 backend. ImageNet pretrained weights are used to initialize weights and biases [1]. The input to the U-Net model is a patch of size of 256×256×3 and the output of the model is a 256×256×1 mask.

#### Training

Binary cross-entropy is used to train the detection model. ADAM is chosen as the optimizer with a fixed learning rate of 0.001. We trained the model for only one epoch with a batch-size of 32. To obtain the training segmentation mask, we expanded the annotated centroid position of the TILs to a square of 12x12 pixels as shown in Figure 6. When tested on the experimental holdout test set, this model resulted in an FROC score of 0.488, which is defined as the average sensitivity values at 10, 20, 50, 100, 200, and 300 false positives per mm$^2$. When tested on the final leaderboard, this model resulted in an FROC score of 0.3205.

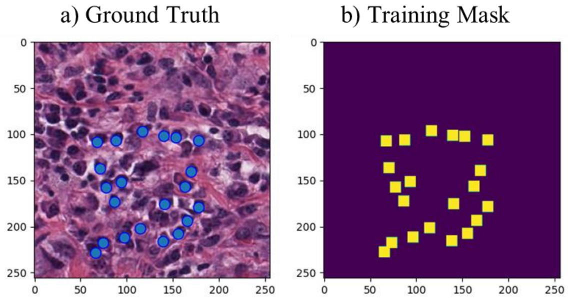


Figure 6. a) A sample training patch along with the annotated TILs. b) The training mask extended the centroid position of the annotated TILs to a square of size of 12 pixels with values of one; everything else in the background has a value of zero.

##### Inference and Postprocessing

After training, the detection model outputs a prediction mask as shown in Figure 7. In order to obtain the (x,y) location for each of the predicted TILs, we first filter out the predictions with probabilities less than 0.1. Next, we apply non-max suppression on a function of distance to obtain the individual location of the TILs from the prediction masks. The non-max suppression distance threshold is set to 12 pixels.

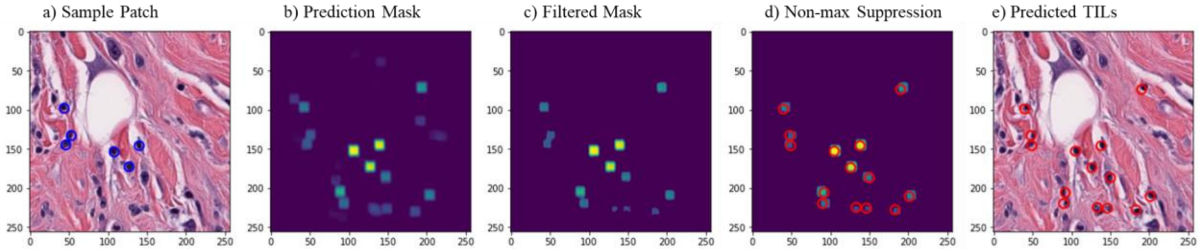


Figure 7. a) A sample patch along with the annotated TILs b) The prediction mask from the detection model on the sample patch c) The prediction mask after filtering the probability values less than 0.1 d) Applying non-max suppression on distance (12 pixels) to obtain the (x,y) locations for each of the predicted TILs. e) Predicted TILs as reflected on the input sample patch.

#### TILs-scoring Algorithm

The TILs-score is estimated as the proportion of TILs area with respect to stromal region area. For that, we first calculate the total area of the stromal regions and count the total number of TILs within the stromal regions for the entire WSI. Then, we estimate the total TILs area by multiplying the total number of TILs by the area of each TIL (8 microns). Finally, we divide the total TILs area by the total area of the stromal region. We tested our model on the experimental holdout test set, and the c-index was 0.68 (95% CI [0.48, 0.87]). For the final leaderboard, the c-index was 0.60 (95% CI [0.37, 0.78]).

#### References

[1] qubvel/segmentation_models: Segmentation models with pretrained backbones. Keras and TensorFlow Keras. (github.com)

### Team: SRI

Vishwesh Ramanathan, Dr. Anne Martel

Affiliation: University of Toronto

Contact: Vishwesh Ramanathan

Code and model availability: https://github.com/Vishwesh4/TigerSubmission

#### Abstract

Tumor Infiltrating Lymphocytes (TILS) are immune cells which have moved from the bloodstream to the tumor site to recognize and kill the cancer cells. Their count in a tumor microenvironment (TME) has been shown to be a very effective biomarker with high prognostic and predictive value, especially in breast cancer. However, manually counting TILs and giving a TIL score in a TME is both time and labor-intensive. TIGER (Tumor Infiltrating Lymphocytes in Breast Cancer) challenge was introduced with the goal of facilitating the development of algorithms that can automatically generate TIL scores with high prognostic value. As part of team SRI, we participate in this challenge and propose a lightweight and fast methodology to compute TIL score, given H&E slides. We propose two networks following the International TILWorking Group guidelines for TIL score evaluation. The first network identifies tumorbed in a given whole slide using patchwise classification. The second network outputs stromal TILs density and stromal tissue density using patch wise regression. We report the TIL score by using the results from these two networks. Our algorithm can process a given wholeslide within 5 minutes and has a final leaderboard score of 0.5996 C-index.

#### Data

Used datasets:

- **WSIROIS**: The patches sampled from this dataset were used for both the tissue area regression and TILs area regression.
- **WSITILS**: This dataset was not used separately to train a model, apart from being used for self-supervised learning
- **WSIBULK**: This dataset was used to perform tumorbed classification. Positive samples (tumor regions) were extracted from three classes in *WSIROI* (Invasive Tumor, Tumour Associated Stroma, Inflamed Stroma, and Lymphocytes) and the negative samples were extracted from outside of the annotated region given in *WSIBULK*.

All the slides were collected and used for self-supervised learning (SSL) to pretrain the encoders of the two models.

Data split:
In total, four datasets were formed

- **Tissue dataset**: 195 slides from *WSIROIS* were used with the help of wholeslidedata package [1]. The slides were split into training (181 slides) and validation (14 slides). Only the tissue segmentation masks were used

- **TILS dataset**: In this data set, 44 slides from *WSIROIS* containing both tissue and cell segmentation masks were used (These 44 slides do not contain any slides from TCGA). Patches were extracted from the 44 slides and saved using wholeslidedata package. These patches contain both tissue and cell segmentation masks. 1767 patches were separately added from the TCGA slides from *WSIROIS*. This was done due to the problems faced while extracting patches from TCGA slides containing cell masks using wholeslidedata package. The extracted patches were mixed and divided into training (80%) and validation (20%) set
- **Tumorbed dataset**: This dataset was formed by extracting patches outside the annotated regions (negative samples) of *WSIBULK* and patches inside regions of Invasive Tumor,Tumour Associated Stroma, Inflamed Stroma, and Lymphocytes (positive samples). For negative sampling [2], the patches from outside annotated regions, were sampled heavily and clustered using K-NN. The patches were then sampled from the top k clusters. All the sampled patches were mixed and divided into training (80%) and validation set (20%)
- **SSL dataset**: Patches were extracted with uniform stride from all the slides in the TIGER dataset. The sampled patches were mixed and divided into training (80%) and validation set (20%)

### Methods

#### Method overview

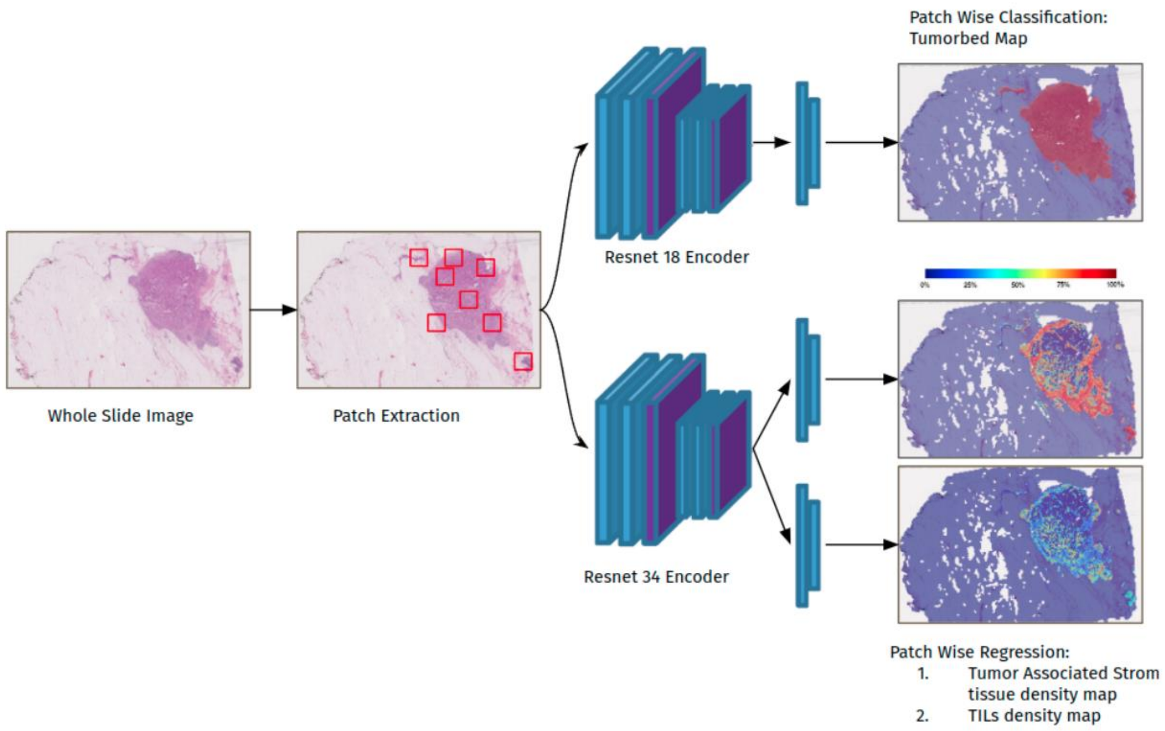


*Figure 1*: Overview of the methodology. The patches from input whole slide goes through two networks. The first network (Tumorbed Classifier), identifies regions of interest i.e tumorbed region using patch wise classification. The second network (TILS Regressor), is a multi-headed network which for a given patch outputs *Tumor Associated Stroma tissue density* and the *TILs density*

#### Self-Supervised Learning

As a first step, self-supervised learning with *Resolution Sequence Prediction* pretext task [3] was used to initialize weights for Resnet 18 and Resnet 34 encoders, which were used by our two models Tumorbed Classifier (top model) and TILS Regressor (bottom model).

#### Tumorbed Classifier

The tumorbed dataset as described in the previous section was used to train the model to classify a patch as relevant (tumorbed region) or non-relevant (outside of the tumorbed region). This model used an architecture consisting of Resnet [4] 18 as the backbone and 2-layer MLP for binary classification.

For training, RandAugment and ColorJitter routines were used for augmentation. Additionally, the ReduceLRonPlateau routine was used for the learning rate schedule. The multifocal loss was used with L2 regularization for the network and was optimized using the Adam optimizer.

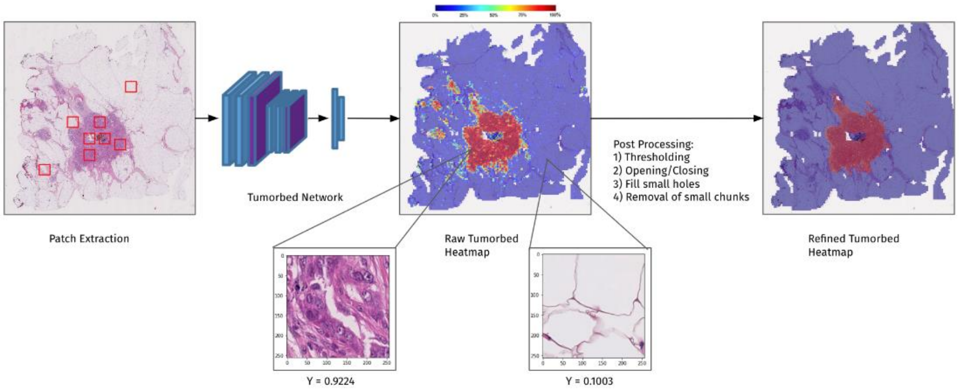


*Figure 2* : Tumorbed network patch wise classification and post processing

#### TILS Regressor

This model used an architecture consisting of Resnet34 as the backbone and has two heads both consisting of 3 layered MLP with the last layer having a sigmoid activation. The two heads are responsible for regression.

1. **TILS head** ($D_{tils}$ ): For each patch, the head calculates the TILS area present only in the tumor-associated stroma and inflamed stroma. The value is normalized to 1.

2. **Tissue head** ($D_{tissue}$ ): For each patch, this head calculates the tissue area consisting of only tumor-associated stroma and inflamed stroma. The value is normalized to 1. The model was first trained using the TILS dataset (described in the previous section). The model was then trained using online sampling on the Tissue dataset with the help of wholeslidedata package. Lastly, the encoder was frozen and the model

was again trained on the TILS dataset with a lower learning rate.

For training, Horizontal/Vertical flips, rotation, Gaussian blur, Brightness contrast, HSV color augmentation, and random affine transformations were used. Additionally, the ReduceLRonPlateau routine was used for the learning rate schedule. The Adam optimizer was used for optimization. L1 loss with L2 regularization was used for this network.

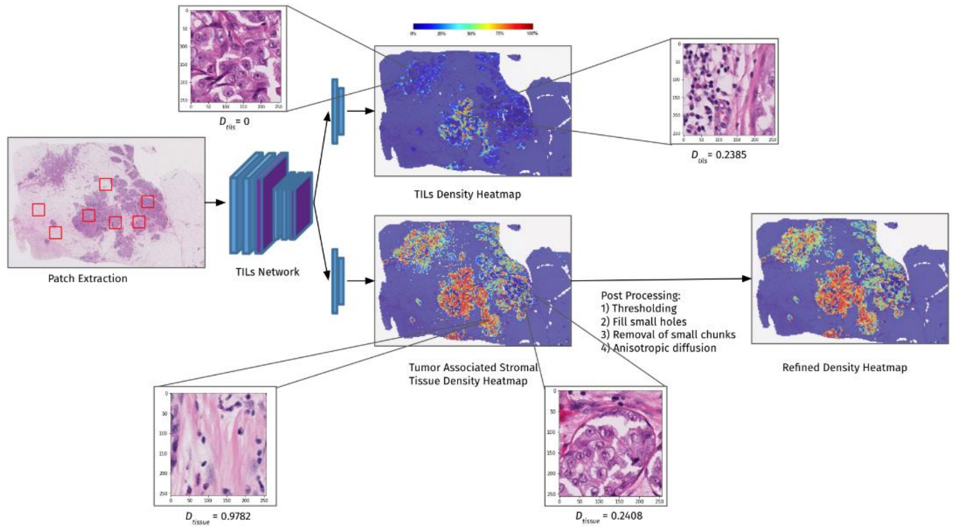


*Figure 3*: TILS network patch wise regression and post processing

#### Inference and Postprocessing

During inference, test-time augmentation was used with the augmentations:

- Vertical flip
- Horizontal flip
- Rotation
- Gaussian Blur

The tumorbed heatmap was postprocessed with

- Thresholding: cutoff = 0.5
- Opening/Closing
    - Resection: kernel size=(3,3), shape: ellipse
    - Biopsy: kernel size=(2,2), shape: ellipse
- Fill small holes: 0.05mm$^2$
- Removal of small chunks
    - Resection: 1.5mm$^2$
    - Biopsy: 0.5mm$^2$

The stromal tissue heatmap was postprocessed with

- Thresholding: cutoff = 0.15
- Fill small holes: 3 pixels
- Removal of small chunks: 3pixels
- Anisotropic Diffusion

#### TIL scoring

The TIL-score was computed as the given formula

**TILs Score**

$$til\,score = \frac{\sum_{i \in \mathbb{P}} (patch_i\ Stromal\ TILs\ density)}{\sum_{i \in \mathbb{P}} (patch_i\ Stromal\ Tissue\ density)} = \frac{\sum_{i \in \mathbb{P}} D^i_{tils}}{\sum_{i \in \mathbb{P}} D^i_{tissue}}$$

Where $\mathbb{P}$ is the set of patches belonging inside the tumorbed. $D_{tils}$ and $D_{tissue}$ are the patch wise Stromal TILs and Stromal Tissue density.

### Team: TIAger

Adam Shephard*, Mostafa Jahanifar*, Ruoyu Wang, Muhammad Dawood, Simon Graham, Kastytis Sidlauskas[2], Ali Khurram[3], Nasir Rajpoot, Shan Raza

Affiliation
* Joint first authors contributed equally.
[1] Tissue Image Analytics Centre, Department of Computer Science, University of Warwick, Coventry, UK
[2] Barts Cancer Institute, Queen Mary University of London, London, UK
[3] School of Clinical Dentistry, University of Sheffield, Sheffield, UK

Contact: Adam Shephard
Code and model availability: https://github.com/adamshephard/TIAger

Preprint: https://arxiv.org/pdf/2206.11943.pdf

#### Abstract

The quantification of tumor-infiltrating lymphocytes (TILs) has been shown to be an independent predictor for prognosis of breast cancer patients. Typically, pathologists give an estimate of the proportion of the stromal region that contains TILs to obtain a TILs score. The Tumor InfiltratinG lymphocytes in breast cancER (TiGER) challenge, aims to assess the prognostic significance of computer-generated TILs scores for predicting survival as part of a Cox proportional hazards model. For this challenge, as the TIAger team, we have developed an algorithm first segment tumor vs. stroma, before localizing the tumor bulk region for TILs detection. Finally, we use these outputs to generate a TILs score for each case. On preliminary testing, our approach achieved a tumor-stroma weighted Dice score of 0.791 and a FROC score of 0.572 for lymphocytic detection. For predicting survival, our model achieved a C-index of 0.719. These results achieved first place across the preliminary testing leaderboards of the TiGER challenge.

#### Data

Used datasets

- WSIROIS: This dataset was used for the training and testing of the Efficient U-Net models for both segmentation and detection.
- WSITILS: This dataset was used for the tuning of the L2 pipeline parameters

Stratified 5-fold cross-validation was used for the training/testing of the segmentation/detection models.

### Methods

The data was augmented using image flipping, rotation, scaling, blurring, distortion (adding noise and JPEG compression), and brightness and colour (hue and saturation) adjustment techniques. Additionally, stain augmentation techniques from TIAToolbox [1] were utilized to randomly change the Haematoxylin and Eosin components of the image. The extent and combination of these augmentation techniques were randomly selected on-the-fly and differ from epoch to epoch.

#### Model architecture

We have implemented Efficient U-Net in TensorFlow for both the segmentation and detection tasks. Efficient U-Net is a fully convolutional network based on an encoder-decoder design paradigm where the encoder branch is the B0 variant of Efficient-Net [1]. These models were pre-trained on ImageNet data.

#### Segmentation

##### Training

We trained the Efficient-UNet to semantically segment the input image into three prediction maps: invasive tumor (label index 1), stroma (label indices 2 and 6), and others (which comprises the background and all other tissue components rather than the first two). The model was trained via a stratified 5-fold cross-validation framework, where each fold contained approximately the same number of images from each of the three subsets of the 'WSIROIS' dataset. For training, patches of size 512 × 512 pixels were extracted at 10× magnification, with a stride of 256 pixels. Images were normalized prior to being fed into the model for training by subtracting the ImageNet mean intensity and dividing by the ImageNet standard deviation. The segmentation model was trained using standard data augmentation techniques, with the addition of stain augmentation. The extent and combination of these augmentation techniques were randomly selected on-the-fly and differ from epoch to epoch. Model training was performed across two phases. First, weights of the encoder were fixed to train the randomly initialized decoder for 10 epochs (Adam optimizer with learning rate of 0.003). We then trained the entire network for an additional 50 epochs using the same optimizer but a reduced learning rate of $4 \times 10{-}4$.

##### Inference

Model Inference. For model inference, we used the top three performing segmentation models from 5-fold cross-validation to obtain an ensembled result. Patches of size 512 × 512 were extracted from the tissue region at 10× magnification with a stride of 256 and a zero-padding of 128 pixels. Inference was then performed on each patch using each of the three segmentation models. The predicted segmentation maps were then averaged and the resulting segmentation mask was then thresholded for each predicted tissue type ($\tau$ stroma = 0.35, $\tau$ tumor = 0.20, optimized over experiments) and morphological opening was performed on the predicted tumor region (disc of radius 10). Following this, each segmentation mask was combined with the corresponding tissue mask. Only the centrally cropped 256 × 256 region was saved to the overall segmentation mask.

#### Detection

##### Training

The model was trained via a stratified 5-fold cross-validation framework, where each fold contained approximately the same number of images from each of the three subsets of the 'WSIROIS' dataset. A binary segmentation was produced for each TIL by dilating each ground truth detection by a radius of 5 pixels using morphological operations. In producing pseudo-segmentation masks for each detection we transformed the detection task into a segmentation task. For training, patches of size 128 × 128 were extracted at 20× magnification, with a stride of 100 pixels. The same image normalization, data augmentation, loss function and two-phase training procedure were employed as for segmentation.

The training dataset for the detection task had a severe class-imbalance, owing to much fewer extracted patches containing TILs than background. To alleviate this challenge, we devised an on-the-fly under-sampling approach which guarantees that the number of patches containing TILs and the ones that do not are almost equal in all batches.

##### Inference

For model inference we used the top three performing models from the 5-fold cross-validation and ensembled their output. Tiles of size 1024 × 1024 were extracted from the tissue region at 20× with a stride of 1024 on inference. The patch was then further divided into sub-patches of size 128 × 128 with an overlap of 28 pixels. These patches were then fed into each of the three detection models. The output segmentation maps were then aggregated across each 1024 × 1024 tile. The output tile segmentations were then averaged to generate an ensembled lymphocyte segmentation map. We then thresholded the output segmentation mask (threshold = 0.3) and performed connected components analysis to find each individual detection. The centroids of the predicted nuclei and the mean intensities were then stored as the output detection coordinates/probabilities

#### TIL scoring

First, we found the 'tumor bulk' region. This was defined using the same pipeline as the baseline algorithm (e.g., morphological operations followed Delaunay triangulation). Following this, the tumor associated stroma was found by taking the overlap between the bulk tissue mask and the stroma from the inferred segmentation mask. We then performed detection in this stroma region alone. Next, we performed slide-level non-maxima suppression on the detected lymphocytes (patch size 2048 × 2048 pixels, at 20× magnification). We then counted the number of lymphocytes within the bulk-stroma area. We multiplied this by an estimated lymphocyte area of 16 microns (optimized on 'WSITILs') and calculated the area of the bulk stroma. The TILs score was calculated by dividing the total predicted TILs area by the stromal area and multiplying by 100. The score was further constrained to be an integer between 0 and 100.

### Team: Biototem

Xu Shuoyu, Ji Zheng, Xie Feng, Kuang Jinbo, Feng Wentai

Affiliation: Bio-totem Pte Ltd, China

Contact: Xu Shuoyu

Algorithm: https://grand-challenge.org/algorithms/bio-totem-tiger-breast-leaderboard-2-algorithm

Code and model availability: https://github.com/biototem/TIGER_challenge_2022

#### Abstract

We describe here the tissue segmentation and nuclei detection methods developed for our participation in the Tiger (Tumor infiltrating lymphocytes in breast cancer) Challenge. The tissue segmentation model was modified from UperNet by adopting a visual attention network as the backbone and adding an auxiliary network to improve the performance. The lymphocyte detection model was modified from SFCN-OPI which is a sibling fully convolutional network with prior objectness interaction to perform lymphoctye detection and false positive suppressing. The results showed that our approaches achieved good performance for both tissue segmentation and lymphocyte detection tasks, and may facilitate the automated scoring of TILs in breast cancer in the future.

#### Data

WSIROIS dataset was used for training tissue segmentation and TILs detection algorithms.

#### Methods

##### Preprocessing

Stain normalization [1] was performed to increase the data size. Except TCGA cohort, we assume there are at least two more cohorts in the dataset which could be recognized from the WSI filename (Cohort 1: filename starting with "TC"; Cohort 2: filename staring with a number). We selected one template image from each of Cohort 1 and 2. All the images from TCGA cohort in the dataset were normalized according to two template images. Moreover, all the images from Cohort 1 were normalized according to the template image from Cohort 2, and vice versa. the original images without stain normalization from three cohorts were put together with the normalized images to form the new dataset for the following processing.

### Segmentation

#### Preprocessing and Augmentation

For the segmentation task, original images were sampled into 512*512 patches at the highest resolution for training. The original label was used without modifications.

The input image was first normalized using the mean and std from the ImageNet. Image augmentation include: random rotation between [-45, +45] degrees, 1, 2, or 3 times random counterclockwise 90° rotation, symmetric transformations, color augmentation in HSV channel, and random scaling between [1, 1.5].

#### Model architecture

The segmentation model we used was UperNet [2] with visual attention

network (VAN) [3] as the backbone. VAN is a neural network based on large kernel attention (LKA) module to enable self-adaptive and long-range correlations in self-attention. Pre-training weights on ImageNet was used to initiate training. Apart from the Uper head, we also added an FCN head as the auxiliary network, and the outputs of both heads were computed separately according to different loss functions and added together as the total loss.

#### Training

<u>Data split</u>

For the segmentation task, the dataset was separated into training, validation and test set at the ratio of 7:2:1. However, since the pixel numbers of each class varied significantly, we exhaustively tested various allocations of training, validation and test sets and kept the combination with the most similar proportions of each class between the three.

The learning rates of the encoder and decoder are slightly different, with the former being 1e-4 and the latter 1e-3. As for the loss function, we used some mixture of Dice and Cross Entropy, where "label smoothing" and "maximal restriction" strategies were included in the CE loss calculation. The "background" class is excluded in the calculation of the loss function. We used the cosine warm-up strategy and the Adam optimizer to train 20 epochs with a batch size of 12.

36000 patches were sampled using the sampling strategy described previously in each epoch. The trained model from each epoch was tested on the validation set, and the best model was selected based on the highest dice score generated from tumor and stroma classes.

#### Postprocessing and Inference

During the image prediction, the original image was divided into 512*512 patches with 50% overlapping. The heatmap of the prediction results of each patch was first generated and multiplied with a Gaussian kernel. The overlapping parts were merged by keeping the larger value to form the final heatmap. The argmax was then applied to the heatmap to generate the segmentation results.

#### Detection

##### Preprocessing and Augmentation

For TILs detection task, original images were sampled into 64*64 patches at the highest resolution for training. The original label was converted to the image mask of point annotations. The center position of each detection box was identified and a circular mask was generated with radius of 3 pixels.

##### Model architecture

Our model was modified from SFCN-OPI [4] which demonstrates a sibling fully convolutional network with prior objectness interaction to perform nuclei detection and classification using weak labels (dot annotation of nuclei center). The original SFCN-OPI first contains a detection branch which is an 80-layer deep FCN. The thresholding would be performed from the output of the detection branch to recognize regions with high confidence of nuclei existence (so called objectness prior) and fed into the fine-grained nuclei classification branch while the classification branch would only focus on these regions. Two branches share features in earlier layers and hold respective higher layers targeting for each specific task. The training was conducted in three-phases: 1) Training the detection branch only; 2) Freeze the detection branch (shared earlier layers) and train the classification branch; 3) Unfreeze the detection branch and train both branches at the same time. The output of both branches will be multiplicated to form the final probability heatmap.

We simply changed the fine-grained nuclei classification branch into a false positive suppressing branch for the TILs detection task. The original multi-class classification task of this branch was changed to a binary classification task (false positive TILs vs true positive TILs from the detection branch). The objectness prior generated from the detection branch would contain as many candidate TILs as possible by lowering the thresholding and the false positive suppressing branch would further eliminate false positive detections.

##### Training

For TILs detection task, the dataset was randomly separated into training, validation and test set at the ratio of 7:2:1.

During training, image augmentation includes random image flips (including vertical and horizontal flips), random Gaussian noise, random brightness enhancement and random sharpening of the images. Each training image is randomly cropped to a size of 64x64 and then fed into the network for training, with same operation performed on the corresponding mask.

For the detection branch we choose the weighted binary cross-entropy loss function. The weights were calculated according to the proportion of positive and negative pixels. For the false positive suppression branch, a modified binary cross-entropy loss function was used. The loss was calculated only from the pixels in therecognize regions (objectness prior) from the thresholded output of the detection branch. The threshold the generated the objectness prior was chosen as 0.6 by several ablation experiments.

The training process was similar to the original implementation in [4]. In the first phase, the detection branch was trained to detect as many TILs as possible. In the second phase, the detection branch is frozen, and the false positive suppression branch was trained to eliminate the false positive TILs detections. In the third phase, both the branches were trained at the same time, to further refine the detection and false positive suppression by the joint training. Each phase was trained for 200 epochs with batch size of 500. The number of patches sampled for each epoch was 10000. The optimizer used is the adam optimizer. Cosine warm-up strategy of the learning rate and the Adam optimizer were used. After each epoch is trained, we calculated the f1-score for TILs detection from the validation set and the best model with highest F1-score was selected as the final model.

##### Postprocessing and Inference

During image prediction, the original image was separated into 1024*1024 big patches with an overlapping of 16 pixels in between. Each 1024*1024 big patch was fed into data loader which would generate 64*64 small patches with an overlapping of 32 pixels in between as input data, and put them to the TILs detection model for prediction. Then we merge these small patches prediction outputs with average method to generate the final big patch prediction heatmap.

The border of each 1024*1024 big patch with the width of 8 pixels were cropped as the prediction at the border of the image was found not reliable. The cropped heatmap of each patch were grouped together to form the entire heatmap. Non- maximum suppressing (NMS) was performed on the heatmap to recognize the local maximum locations as the detected TILs. A detected TIL would be removed if the probability of the local maximum point was too low (less than 0.15). We found that the majority of the false positive detection from our model appeared in non-stroma regions and there are very few true positive TILs exist in these non-stroma regions, hence we finally removed all the TILs detected in regions other than stroma to decrease the false positive ratio while not sacrificing detection sensitivity too much.

#### TIL scoring

The segmentation was first performed on each WSI. Compared to the segmentation performed in L1 with small size of ROI, the entire WSI needs to be segmented in L2 which is too large to put in the memory for processing. We designed a "line-by-line Gaussian kernel weighted fusion" strategy to segment the entire WSI. An image cache was created in the memory which has the width of the WSI and the height of the image patch (512 pixels). This line image was first segmented with 50% overlapping 512*512 patches to generate the merged result sas in L1. Next the top half of the line image was written to the hard disk and the bottom half of the line image was kept in the cache to merge with the top half of the next line image. In this way, all the patches could be merged similar to the implementation in L1 from the entire WSI. During the prediction, only the patches with the percentage of tissue area more than 5% (calculated from the provided tissue mask) would be segmented.

Tumor bulk area was then generated from the segmented mask. We first grouped three classes (invasive tumor, tumor associated stroma and inflamed stroma) into one class (assigned pixel value of 1) and set all the other pixels into zeros. All the connected components were then identified and the connected components with the size smaller than the 10% of the largest connected component size in the WSI were removed. Moreover, a connected component

would also be removed if the percentage of invasive tumor areas inside was less than 5%. The remaining connected components were kept as the tumor bulk. The TILs detection was only performed in the stroma areas (include tumor associated stroma and inflamed stroma) inside tumor bulk. The detection process was similar to the procedure in L1. The cut-off threshold of the prediction heatmap was set to 0.15 to identify each TIL which was same as in L1.

##### TIL score

We followed the same way to calculate TIL score as presented in Tiger Challenge baseline. The number of TILs detected in the stroma areas inside tumor bulk was counted and multiplied by an estimated TIL nuclei size to represent the TIL area. This TIL area was finally divided by the area of stroma areas inside tumor bulk and multiplied by 100 to generate the TIL score.

### Team: AIVIS

Ming Fan, Daehong Lee, Jaehyung Ye, Kangwon Byun, Jeongyeol Kim

Affiliation: Aivis Inc

Contact: Daehong Lee

Grandchallenge algorithm: https://grand-challenge.org/algorithms/try-docker

Code and model availability: https://github.com/AIVIS-MING/TIGER_SEG-DET

#### Abstract

In this challenge, we proposed a novel network for segmenting the tumor, stroma and others. In our method, we use the upper-net as our baseline method. To improve the performance, we added the attention module to the bottlenecks of the backbone ResNet-101, To produce a segmentation result with fine details, similar to the expanding path of the U-net, the feature map is fed into the decode part which is composed of a series of convolution and up-sampling layers. For detection, we used a hybrid approach to detect the lymphocytes and plasma cells. We first detect the cells by using five state-of-the-art algorithms and weighted fuse the estimated bounding boxes according to their confidence scores.

#### Data

In the phase of segmentation and detection, we use the data refer to WSIROIS.

##### Segmentation

###### Preprocessing and Augmentation

For segmentation, we tiled whole slide images into image patches with resolution 2048x2048 according to the available segmentation annotation. To train the network, we split all the captured images in the ratio of 9:1. In our experiments, two-fold cross validation was utilized. In the training pipeline, for each image patch, we randomly change the brightness, contrast, hue and saturation. In addition, we swap the channels of each image patch.

#### Model architecture

For segmentation, we proposed a novel encode-decode network as shown in the Figure below.

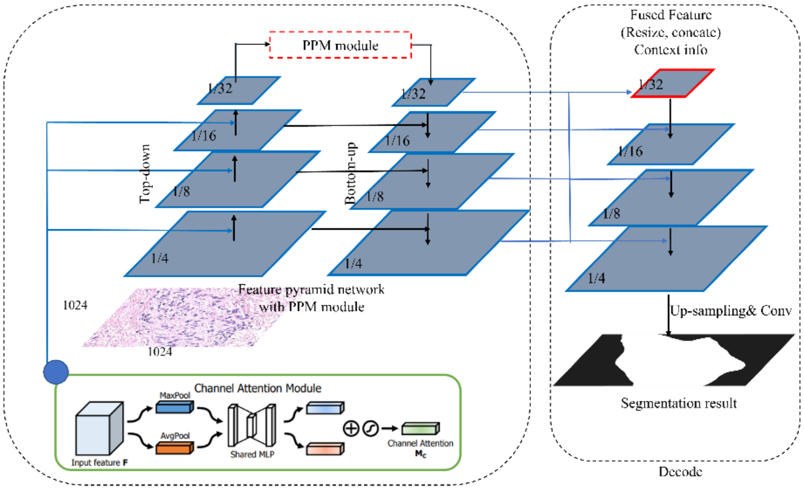


In the encode part, we incorporate the channel attention module [1] after each stage of Resnet-101. To learn the robust features, the PPM module [2] is further used to capture the global contextual cues. The obtained feature map is then fed into the decode part to produce the segmentation map with the skip-connection module. The channel attention module is added at bottlenecks of the ResNet where the down-sampling of feature maps occurs; the feature map is first aggregated by both global and max polling. Next, the attention map is obtained by a 1x1 convolution, followed by a multi-layer perceptron with one hidden layer [1]. The feature map is obtained from the element-wise product of the attention map with the input feature map. Before feeding the feature map into the decode part, PPM module used in uperNet [2] is utilized for understanding the contextual interactions of the features in the scene globally. To this end, the feature map from the last layer of the backbone is first pooled with different ratios. Then, the pooled feature maps are concatenated and convoluted to produce a feature map with 512 channels. To produce a segmentation result with fine details, similar to the expanding path of the U-Net [3], the feature map is fed into the decode part which is composed of a series of convolution and up-sampling layers. Every step in the expansive path consists of an up-sampling of the feature map followed by a 2x2 convolution ("up-convolution") that halves the number of feature channels, a concatenation with the correspondingly cropped feature map from each stage of encode, and two 3x3 convolutions, each followed by a ReLU. For more detail, please refer to the Unet [3]. Since only four stages are used in the encode part, the number of the upsampling layer is set to 4.

#### Training

The networks used in this study were developed using the open-source mmSegmentation [5].

The parameters of encode part used for training were the same as those of the config file provided by mmSegmentation if not mentioned otherwise. We used the AdamW optimizer with an initial learning rate of 0.0002, weight decay of 0.05. The batch size is set to 16 and the cross-entropy is used as a loss function for training. The segmentation model was trained on two NVIDIA 3090 GPUs for eight hours. The weight with the best validation accuracy was saved. Due to runtime limitation, two-fold cross validation is used in the experiment.

##### Inference and Postprocessing

As mentioned above, two-fold cross validation is utilized to train our model. For each image in the test dataset, we computed the probability of each pixel by using the two trained models; the probability belongs to tumor, stroma or others. The final segmentation result was obtained by labeling each pixel according to the computed the mean probability.

#### Detection

##### Preprocessing and Augmentation

For detection, we extracted the image patch with two different resolutions (128x128 and 256x256) in the valid box region which was obtained from the available annotation. If the valid box region is smaller than 256x256, the outside region of image patch is padded with 114.

The dataset set are split in the ratio of 9:1. In the training pipeline, for each sample, we resized the image patch and randomly flip the image with probability of 0.5. The more detail parameter setting of our algorithm be found in https://github.com/AIVIS-MING/TIGER_SEG-DET.

##### Model architecture

For detection, we ensemble 4 detection models to produce the final result. The model used in our method are: yolov5 [4], cascade RCNN with resnext101 backbone, deformable detr with resnext101 backbone and ATSS with swin transformer backbone. For each input image, four networks are performed and obtained the final result via non-maximum non-maximum suppression.

##### Training

Similar to the segmentation, we used the open-source mmDetection to produce the detection result. In training phase, the weight with best mAP for validation set is stored and used for prediction. The parameter for training were the same as those of the config file proved by mmDetection if not mentioned otherwise. For cascade-based method, the scales and base_sizes of the anchor generator are set to 4. For yolov5, we trained the model based on the default parameter setting provided by the publicly available open source [6].

##### Postprocessing and Inference

After obtaining the detection boxes in each model, we used the Weighted Box Fusion method [7] for merging the boxes. This method gives better performance comparing with simple non-maximum suppression. The figure shown below gives an overview of the postprocessing.

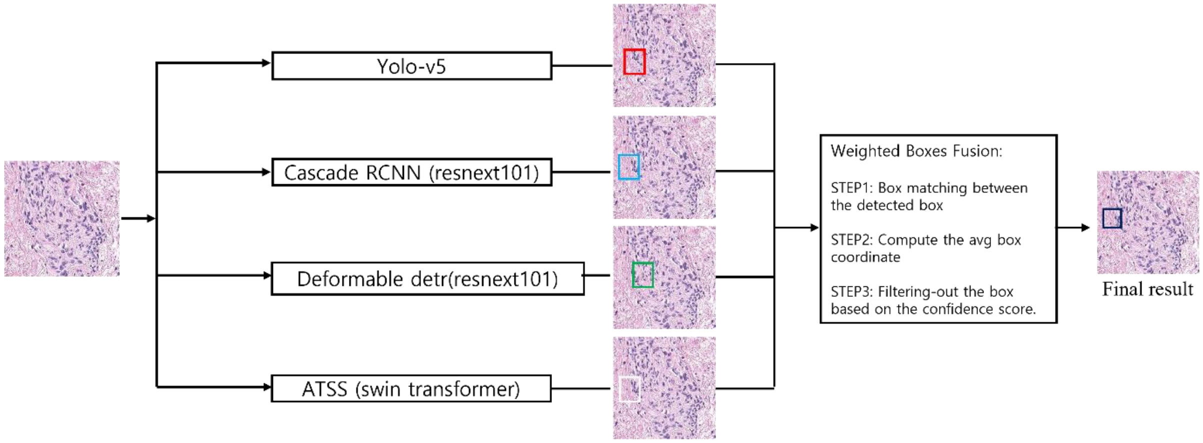

[4] https://github.com/ultralytics/yolov5

[5] https://github.com/open-mmlab/mmsegmentation

[6] https://github.com/open-mmlab/mmdetection

[7] Weighted Boxes Fusion: Solovyev R, Wang W, Gabruseva T. Weighted boxes fusion: Ensembling boxes from different object detection models[J]. Image and Vision Computing, 2021, 107: 104117.

### Team: CellsVision

Xulin Chen, Liliang Chen
Cells Vision (Guangzhou) Medical Technology Inc.
Contact: Xulin Chen

Work done of each team member:
Xulin Chen designed and developed the algorithm, did the data analysis, trained and developed the AI models, updated the inference codes, built the docker image, and wrote the document of the developed algorithm.
Liliang Chen provided guidance for building the docker image.

Code and model availability: https://github.com/XulinChen/Algorithm-for-Tiger-Challenge

#### Abstract

The TIGER challenge is to evaluate computer algorithms for the automated assessment of tumor-infiltrating lymphocytes (TILs) on H&E histopathological images in breast cancer. Our approach begins with the utilization of a semantic segmentation model to segment patches within a whole slide image (WSI). Subsequently, patches containing a threshold number of tumor tissues are selected based on the segmentation results and passed through a detection model to pinpoint lymphocytes cells. Then, the TILs score could be calculated for each patch based on our well-designed formula by introducing specific parameters to characterize the degree of infiltration between invasive tumor and lymphocytes cells. Finally, we employ a tumor-guided aggregation technique to obtain the overall TILs score for the entire WSI.

#### Data

For semantic segmentation, besides all the images from the directory “wsirois/roi-level-annotations/tissue-bcss”, the images from “wsirois/roi-level-annotations/tissue-cells” with sizes exceeding 1000 pixels are also used for segmentation training. For detection, all the images in the directory “wsirois/roi-level-annotations/tissue-cells” are utilized for detection training. For tils-scoring, all the images in “wsitils” are used. The segmentation and detection tasks follow a training/validation split ratio of 5:1. For TILs scoring, where only post-processing steps are introduced, no validation set is established.

### Methods

#### Segmentation

##### Preprocessing and Augmentation

Given the ROIs of the segmentation datasets, multiple patches with resolutions between [1024, 2048] are cropped randomly from the ROIs. These patches serve as the training images for segmentation. Subsequently, the patches are resized to 1024×1024 pixels and are input to the segmentation network.

##### Model architecture

The backbone network of semantic segmentation is HRNet-W18[1]. The training procedure of semantic segmentation is illustrated in Figure 1.

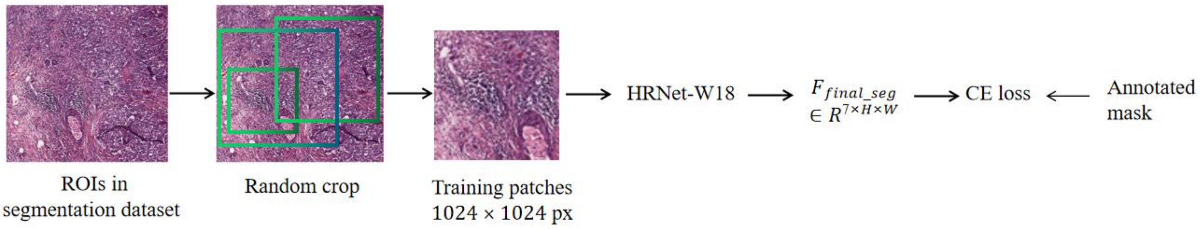


Figure 1. The training procedure of semantic segmentation.

##### Training

The feature maps produced by the HRNet-W18[1] are denoted as $F_{seg} \in R^{270\times\frac{H}{4}\times\frac{W}{4}}$, where $H$ and $W$ represent the height and width of the input images. Utilizing the feature maps $F_{seg} \in R^{270\times\frac{H}{4}\times\frac{W}{4}}$, a 1×1 convolutional layer with the channel size of 270, the stride of 1 and ReLU activation is employed, yielding the feature maps $F'_{seg} \in R^{270\times\frac{H}{4}\times\frac{W}{4}}$. Subsequently, a 1×1 convolutional layer with the channel size of 7, the stride of 1 is applied, resulting in the final feature maps $F_{final\_seg} \in R^{7\times\frac{H}{4}\times\frac{W}{4}}$, where 7 represents the number of classes for semantic segmentation. Following this, the $F_{final\_seg} \in R^{7\times\frac{H}{4}\times\frac{W}{4}}$ is sampled to $F_{final\_seg} \in R^{7\times H\times W}$ through bilinear interpolation. The softmax function is used to the $F_{final\_seg} \in R^{7\times H\times W}$. Afterward, the prediction of the network is optimized utilizing the annotated mask and cross entropy loss.

The backbone HRNet-W18 is pretrained on the Imagenet. The Stochastic Gradient Descent (SGD) optimizer is employed with an initial learning rate of 0.01, which decays by 0.1 after 70 epochs. The batch size is configured as 12 on four NVIDIA GeForce RTX 1080Ti GPUs.

##### Inference

During the inference stage of segmentation, non-overlapped patches of size 2048×2048 pixels are extracted from the WSI at a 20× objective magnification, with the stride of 2048 pixels. Subsequently, the patches are resized to 1536×1536 pixels and fed into the segmentation model, producing the predicted segmentation map $F_{final\_seg} \in R^{7\times\frac{H}{4}\times\frac{W}{4}}$. Following this, the argmax function is applied to $F_{final\_seg} \in R^{7\times\frac{H}{4}\times\frac{W}{4}}$, yielding the predicted segmentation label $F_{seg\_label} \in R^{1\times\frac{H}{4}\times\frac{W}{4}}$. Further, the $F_{seg\_label} \in R^{1\times\frac{H}{4}\times\frac{W}{4}}$ is sampled to $F'_{seg\_label} \in R^{1\times 2048\times 2048}$ using nearest neighbor interpolation. Afterward, the elements of $F'_{seg\_label}$ add one, and the results are multiplied by the corresponding tissue mask, resulting in the final result of segmentation.

##### Postprocessing

The segmentation mask of each patch is combined to generate the final segmentation mask for the WSI.

#### Detection

##### Preprocessing and Augmentation

Due to the substantial variation in the sizes of ROIs within the detection dataset, the training of the detection model involves two steps. In the initial phase, multiple patches with sizes ranging from 150 to 170 are randomly extracted from all ROIs. Subsequently, these patches are resized to 160×160 pixels and input to the detection network. In the second phase, patches ranging in size from 700 to 800 are randomly cropped from ROIs larger than 1000 pixels. These patches are then resized to 768×768 pixels and input into the detection network. In the above two steps, the ROIs with a higher number of lymphocytes cells were more likely to be cropped. The model trained in the first step serves as the initialization for the parameters of the model in the second step.

##### Model architecture

The backbone network for cell detection is also HRNet-W18, but it does not share the same parameters with the model of semantic segmentation. The feature map output by the backbone is denoted as $F_{det} \in R^{270\times\frac{H}{4}\times\frac{W}{4}}$, with $H$ and $W$ representing the height and width of the input images. The overall training procedure of cell detection is shown in Figure 2.

##### Training

Utilizing the feature map $F_{det} \in R^{270\times\frac{H}{4}\times\frac{W}{4}}$, a 1×1 convolutional layer with the channel size of 270, the stride of 1 and ReLU activation is employed, producing the feature maps $F'_{det} \in R^{270\times\frac{H}{4}\times\frac{W}{4}}$. Following this, a 1×1 convolutional layer with the channel size of 1, the stride of 1 is applied, resulting in the final feature maps $F_{final\_det} \in R^{1\times\frac{H}{4}\times\frac{W}{4}}$.

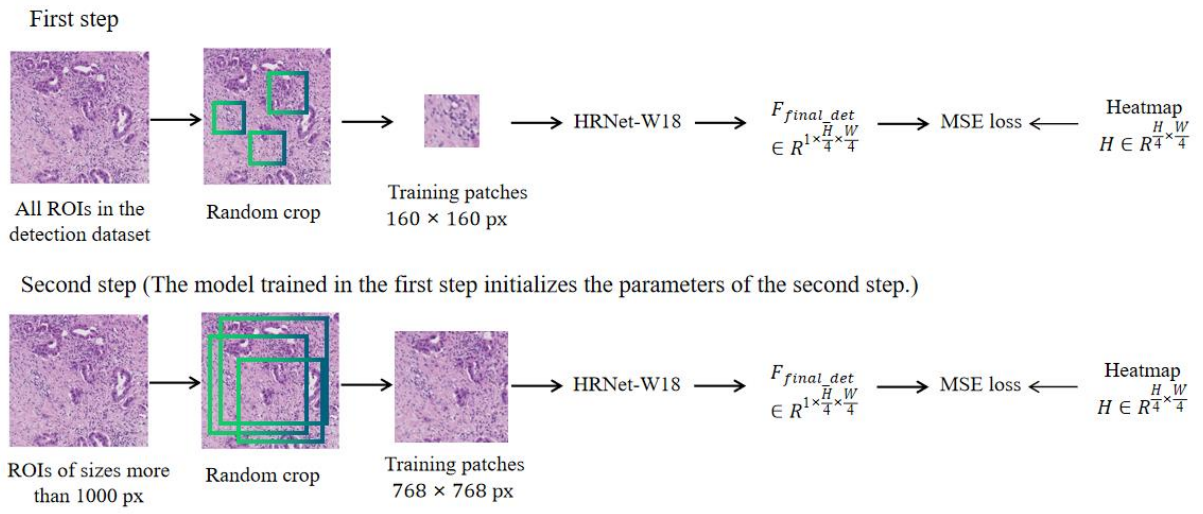


Figure 2. The training procedure of lymphocytes-cells detection.

Obtaining the ground-truth bounding boxes of the cells (lymphocytes cells), the center points of the bounding boxes are denoted as $T = \{(x_1^c, y_1^c), (x_2^c, y_2^c), \dots, (x_N^c, y_N^c)\}$, where $c$ signifies the center point of the bounding box, and $N$ denotes the number of cells in a given image. Subsequently, the Gaussian kernel is utilized to compute the heatmap $H \in R^{\frac{H}{4}\times\frac{W}{4}}$ using the formula as

$$H_{x,y} = e^{-\frac{(x-x_i^c)^2+(y-y_i^c)^2}{2\sigma^2}} \quad (1)$$

where $\delta = 1$, $i$ represents the $i$-th center point. The mean squared error loss function is used for cell detection, expressed as

$$L_{mse} = \frac{1}{\frac{H}{4}\times\frac{W}{4}} \sum_{x=1,y=1}^{x=\frac{W}{4},y=\frac{H}{4}} (H(x,y) - F_{final\_det}(x,y))^2 \quad (2)$$

The Adam optimizer is employed with an initial learning rate set to 0.0001, which decreases by a factor of 0.1 after 30 epochs. For the first step, the batch size is configured as 24, and for the second step, it is set to 12.

##### Inference

In the inference phase of the detection model, patches of size 2048×2048 pixels with the number of tumor tissues surpassing the threshold $T_{tm}$ are collected. Then these patches are resized to 1536×1536 pixels and fed into the detection model. The predicted detection maps $F_{final\_det} \in R^{1\times\frac{H}{4}\times\frac{W}{4}}$ are produced.

##### Postprocessing

To compute the area of lymphocytes cells for the subsequent TILs scoring and optimize processing time, the pixels of the predicted map $F_{final\_det} \in R^{1\times\frac{H}{4}\times\frac{W}{4}}$ with values exceeding the threshold of 0.01 are collected. The count of the collected pixels is regarded as the area of lymphocytes cells and denoted as $N_{lym\_cells}$.

#### TIL scoring

The procedure for inference of the WSI and TIL scoring is demonstrated in Figure 3.

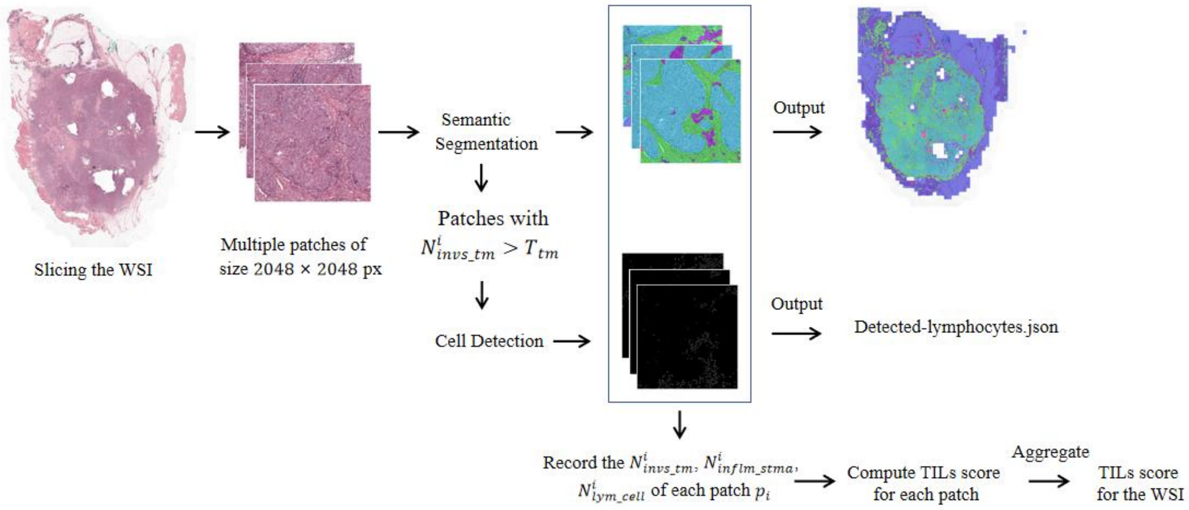


Figure 3 The procedure for inference of the WSI and TIL scoring.

Firstly, the TILs score is computed for each patch. Utilizing the predicted segmentation label $F_{seg\_label} \in R^{1\times\frac{H}{4}\times\frac{W}{4}}$ of each patch, the number of pixels corresponding to invasive tumor and inflamed stroma are recorded and denoted as $N^i_{invs\_tm}$ and $N^i_{inflm\_stma}$, respectively, where *i* signifies the *i*-th patch of the WSI. Additionally, the area of lymphocytes cells $N^i_{lym\_cells}$ for each patch is recorded.

Following that, three base numbers are defined, which are represented as $N_{base1}$, $N_{base2}$ and $N_{base3}$. The TILs scoring based on $N^i_{inflm\_stma}$ and denoted as $TIL^i_A$, is calculated as follows:

$$TIL^i_A = \frac{N^i_{inflm\_stma}}{N_{base1}} \cdot \max(1, \frac{N^i_{invs\_tm}}{N_{base2}}) \quad (3)$$

where *i* represents the *i*-th patch of the WSI. Then, the TILs scoring based on $N^i_{lym\_cells}$, denoted as $TIL^i_B$, is calculated in a similar way as

$$TIL_B^i = \frac{N_{lym\_cells}^i}{N_{base3}} \cdot \max(1, \frac{N_{invs\_tm}^i}{N_{base2}}) \quad (4)$$

In the equation (3) and (4), some specific base numbers are introduced to describe the degree of infiltration between invasive tumor and lymphocytes cells in the patch level.

Subsequently, the TILs score for the entire WSI, based on $TIL_A^i$, is computed as

$$TIL_A = \frac{1}{NUM(N_{invs\_tm}^i > T_{tm})} \sum_{N_{invs\_tm}^i > T_{tm}} TIL_A^i \quad (5)$$

where $T_{tm}$ represents the threshold for the number of invasive tumor, and $NUM(N_{invs\_tm}^i > T_{tm})$ denotes the count of patches with $N_{invs\_tm}^i$ greater than $T_{tm}$. The equation (5) implies that only the patches with a specific number of invasive tumor contribute to the calculation of TILs score for the entire WSI.

Similarly, the TILs score for the entire WSI, derived from $TIL_B^i$, is computed as

$$TIL_B = \frac{1}{NUM(N_{invs\_tm}^i > T_{tm})} \sum_{N_{invs\_tm}^i > T_{tm}} TIL_B^i \quad (6)$$

The ultimate TILs score for the entire WSI is determined by both $TIL_A$ and $TIL_B$, expressed as

$$TIL = \min(\frac{1 \cdot TIL_A + 2 \cdot TIL_B}{3} \times 100, 95) \quad (7)$$

The output TILs score is in the range of [0, 95]. In the setting of this algorithm, all the three base numbers $N_{base1}$, $N_{base2}$ and $N_{base3}$ are set to around 1/6 of the total number of pixels of a patch, and $T_{tm}$ is set to around 1/20 of the total number of pixels of a patch. These settings are based on the statistical analysis on the data of the wsitils dataset. In the future work, it is worth exploring to learn the $N_{basei}$ ($i$ = 1, 2, 3) and $T_{tm}$ in an end-to-end manner.
